# Whole genome sequencing identifies multiple loci for critical illness caused by COVID-19

**DOI:** 10.1101/2021.09.02.21262965

**Authors:** Athanasios Kousathanas, Erola Pairo-Castineira, Konrad Rawlik, Alex Stuckey, Christopher A Odhams, Susan Walker, Clark D Russell, Tomas Malinauskas, Jonathan Millar, Katherine S Elliott, Fiona Griffiths, Wilna Oosthuyzen, Kirstie Morrice, Sean Keating, Bo Wang, Daniel Rhodes, Lucija Klaric, Marie Zechner, Nick Parkinson, Andrew D. Bretherick, Afshan Siddiq, Peter Goddard, Sally Donovan, David Maslove, Alistair Nichol, Malcolm G Semple, Tala Zainy, Fiona Maleady-Crowe, Linda Todd, Shahla Salehi, Julian Knight, Greg Elgar, Georgia Chan, Prabhu Arumugam, Tom A Fowler, Augusto Rendon, Manu Shankar-Hari, Charlotte Summers, Charles Hinds, Peter Horby, Danny McAuley, Hugh Montgomery, Peter J.M. Openshaw, Yang Wu, Jian Yang, Paul Elliott, Timothy Walsh, GenOMICC Investigators, 23andMe, Covid-19 Human Genetics Initiative, Angie Fawkes, Lee Murphy, Kathy Rowan, Chris P Ponting, Veronique Vitart, James F Wilson, Richard H Scott, Sara Clohisey, Loukas Moutsianas, Andy Law, Mark J Caulfield, J. Kenneth Baillie

## Abstract

Critical illness in COVID-19 is caused by inflammatory lung injury, mediated by the host immune system. We and others have shown that host genetic variation influences the development of illness requiring critical care^1^ or hospitalisation^2;3;4^ following SARS-Co-V2 infection. The GenOMICC (Genetics of Mortality in Critical Care) study recruits critically-ill cases and compares their genomes with population controls in order to find underlying disease mechanisms.

Here, we use whole genome sequencing and statistical fine mapping in 7,491 critically-ill cases compared with 48,400 population controls to discover and replicate 22 independent variants that significantly predispose to life-threatening COVID-19. We identify 15 new independent associations with critical COVID-19, including variants within genes involved in interferon signalling (*IL10RB, PLSCR1*), leucocyte differentiation (*BCL11A*), and blood type antigen secretor status (*FUT2*). Using transcriptome-wide association and colocalisation to infer the effect of gene expression on disease severity, we find evidence implicating expression of multiple genes, including reduced expression of a membrane flippase (*ATP11A*), and increased mucin expression (*MUC1*), in critical disease.

We show that comparison between critically-ill cases and population controls is highly efficient for genetic association analysis and enables detection of therapeutically-relevant mechanisms of disease. Therapeutic predictions arising from these findings require testing in clinical trials.

## Introduction

Critical illness in COVID-19 is both an extreme disease phenotype, and a relatively homogeneous clinical definition including patients with hypoxaemic respiratory failure^5^ with acute lung injury,^6^ and excluding many patients with non-pulmonary clinical presentations^7^ who are known to have divergent responses to therapy.^8^ In the UK, the critically-ill patient group is younger, less likely to have significant comorbidity, and more severely affected than a general hospitalised cohort,^5^ characteristics which may amplify observed genetic effects. In addition, since development of critical illness is in itself a key clinical endpoint for therapeutic trials,^8^ using critical illness as a phenotype in genetic studies enables detection of directly therapeutically-relevant genetic effects.^1^

Using microarray genotyping in 2,244 cases, we previously reported that critical COVID-19 is associated with genetic variation in the host immune response to viral infection (*OAS1, IFNAR2, TYK2*) and the inflammasome regulator *DPP9*.^1^ In collaboration with international groups, we recently extended these findings to include a variant near *TAC4* (rs77534576).^2^ Several variants have been associated with milder phenotypes, such as the need for hospitalisation or management in the community, including the ABO blood type locus,^4^ a pleiotropic inversion in chr17q21.31,^9^ and associations in 5 additional loci including the T lymphocyte-associated transcription factor, *FOXP4*.^2^ An enrichment of rare loss-of-function variants in candidate interferon signalling genes has been reported,^3^ but this has yet to be replicated at genome-wide significance thresholds.^10;11^

We established a partnership between the GenOMICC Study and Genomics England to perform whole genome sequencing (WGS) to improve resolution and deepen fine-mapping of significant signals to enhance the biological insights into critical COVID-19. Here, we present results from a cohort of 7,491 critically-ill patients from 224 intensive care units, compared with 48,400 population controls, describing discovery and validation of 22 gene loci for susceptibility to life-threatening COVID-19.

## Results

### Study design

Cases were defined by the presence of COVID-19 critical illness in the view of the treating clinician - specifically, the need for continuous cardio-respiratory monitoring. Patients were recruited from 224 intensive care units across the UK in the GenOMICC (Genetics Of Mortality In Critical Care) study. As a control population, unrelated participants recruited to the 100,000 Genomes Project were selected, excluding those with a known positive COVID-19 test, as severity information was not available. The 100,000 Genomes Project cohort (100k cohort) is comprised of UK individuals with a broad range of rare diseases or cancer and their family members. We included an additional prospectively-recruited cohort of volunteers (mild cohort) who self-reported testing positive for SARS-CoV-2 infection, and experienced mild or asymptomatic disease.

### GWAS analysis

Whole genome sequencing and subsequent alignment and variant calling was performed for all subjects as described below (Methods). Following quality control procedures, we used a logistic mixed model regression, implemented in SAIGE,^12^ to perform association analyses with unrelated individuals (critically-ill cases *n* = 7, 491, controls (100k) *n* = 46, 770, controls (mild COVID-19) *n* = 1, 630) (Methods, Supplementary Table 2). 1,339 of these cases were included in the primary analysis for our previous report.^1^ Genome wide association studies (GWAS) were performed separately for genetically predicted ancestry groups (European - EUR, South Asian - SAS, African - AFR, East Asian - EAS, see Methods). Subsequently, we conducted inverse-variance weighted fixed effects meta-analysis across the four predicted ancestry cohorts using METAL^13^ (Methods). In order to reduce the risk of spurious associations arising from genotyping or pipeline errors, we required supporting evidence from variants in linkage disequilibrium for all genome-wide significant variants: observed z-scores for each variant were compared to imputed z-scores for the same variant, with discrepant values being excluded (see Methods, Supplementary Figure 12).

In population-specific analyses, we discovered 22 independent genome-wide significant associations in the EUR ancestry group (Figure 1, Supplementary Figure 11 and Table 1) at a *P*-value threshold adjusted for multiple testing for 2,264,479 independent linkage disequilibrium-pruned genetic variants: 2.2 × 10^−08^ (Supplementary Table 3). The strong association at 3p21.31 also reached genome-wide significance in the SAS ancestry group (Supplementary Figure 11).

**Figure 1:**
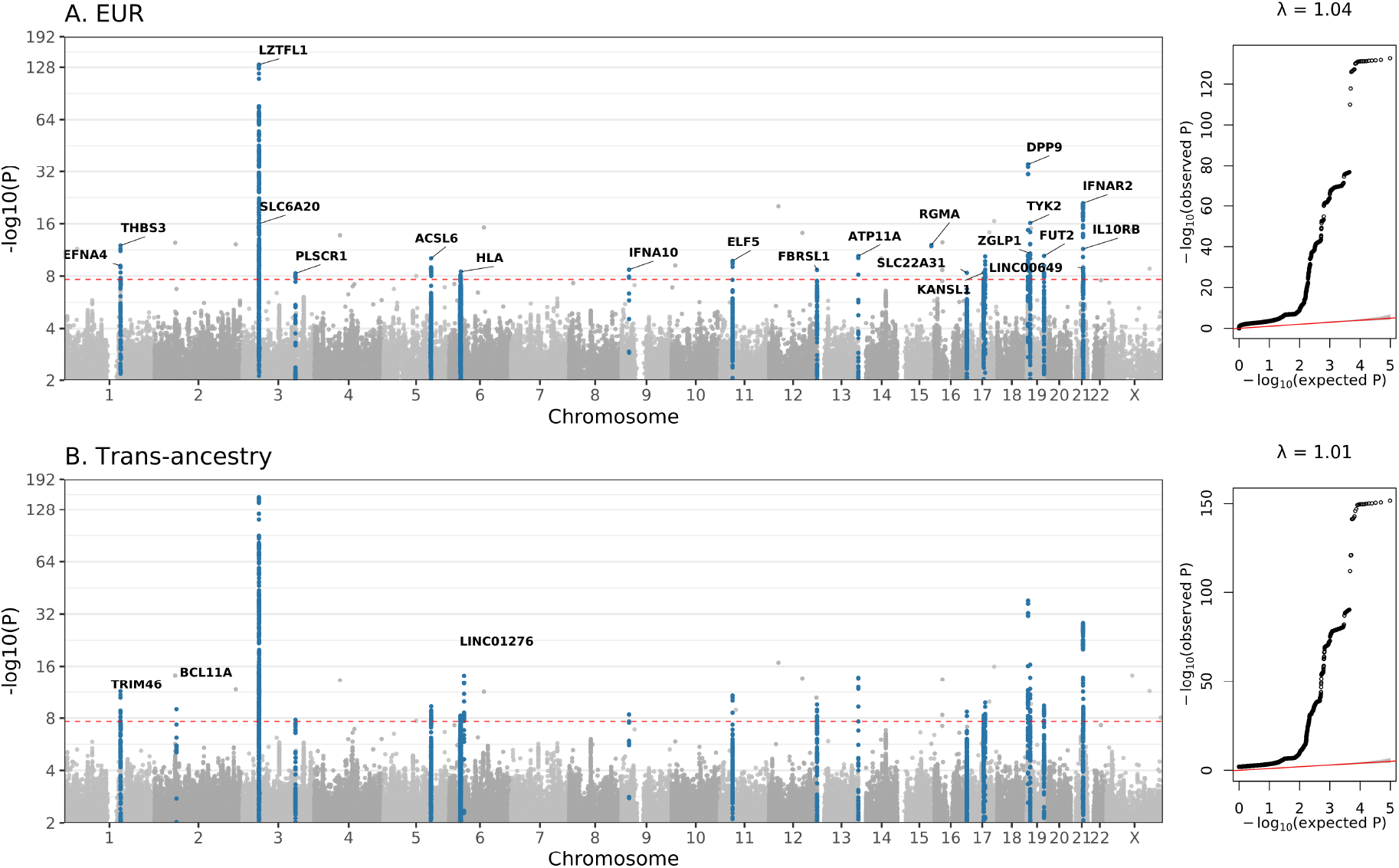
GWAS results for EUR ancestry group, and trans-ancestry meta-analysis. Manhattan plots are shown on the left and quantile–quantile (QQ) plots of observed versus expected *P* values are shown on the right, with genomic inflation (*λ*) displayed for each analysis. Highlighted results in blue in the Manhattan plots indicate variants that are LD-clumped (*r*^2^=0.1, *P*_2_=0.01, EUR LD) with the lead variants at each locus. Gene name annotation by Variant Effect Predictor (VEP) indicates genes impacted by the predicted consequence type of each lead variant. The red dashed line shows the Bonferroni-corrected *P*-value=2.2 × 10^−8^.

**Table 1:**
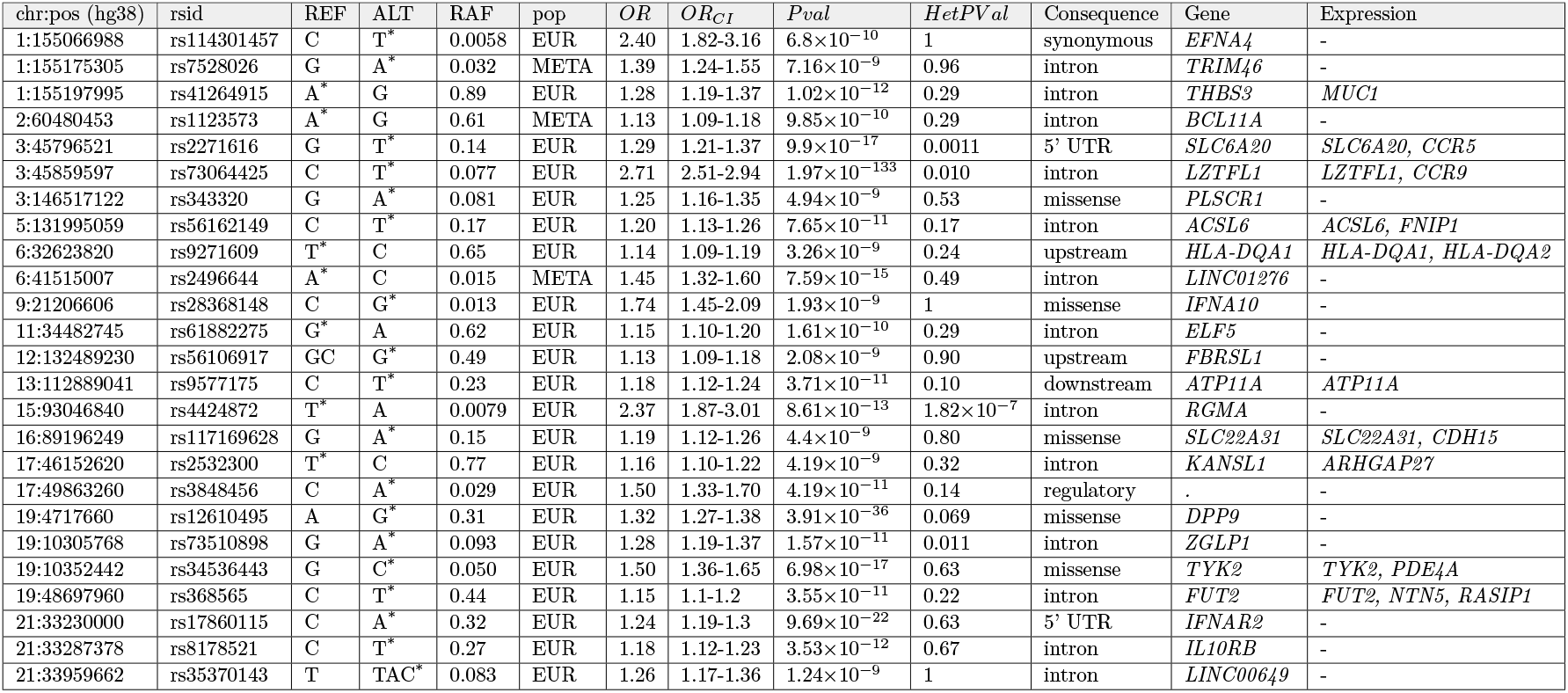
Lead variants from independent regions in the per-population GWAS and trans-ancestry meta-analysis. Variants and the reference and alternate allele are reported with hg38 build coordinates. Asterisk (*) indicates the risk allele. For each variant, we report the risk allele frequency in Europeans (RAF), the odds ratio and 95% confidence interval, and the association *P*-value. Consequence indicates the worst consequence predicted by VEP99, and Gene indicates the VEP99-predicted gene, but not necessarily the causal mediator. Expression indicates genes where is evidence of gene expression affecting COVID-19 severity, found by TWAS and colocalisation analysis.

In trans-ancestry meta-analysis, we identified an additional three loci with genome-wide significant associations (Figure 1, Table 1). We tested the meta-analysed set of 25 loci for heterogeneity of effect size between predicted ancestries and detected significant (at *P* < 1.83 × 10^−3^) evidence for heterogeneity for two variants (Table 1, Supplementary Figure 13).

Fine mapping of the association signals revealed putative causal variants for several genes (See Supplementary Information). For example, we detected variants at 3q24 and 9p21.3 predicted to be missense mutations by Variant Effect Predictor (VEP). These impact *PLSCR1* and *IFNA10* respectively, and both are predicted to be deleterious by the Combined Annotation Dependent Depletion (CADD) tool^14^ (*PLSCR1* (chr3:146517122:G:A, rs343320,p.His262Tyr, OR:1.24, 95%CIs [1.15-1.33], CADD:22.6; *IFNA10* (chr9:21206606:C:G, rs28368148, p.Trp164Cys, OR:1.74, 95% CIs [1.45-2.09], CADD:23.9). Structural predictions for these loci suggest functional effects (Figure 3 and Supplementary Figure 15).

### Replication

Replication was performed using summary statistics generously shared by collaborators: data from the COVID-19 Host Genetics Initiative (HGI) data freeze 6 were combined using meta-analysis with data shared by 23andMe (Methods). Although the HGI programme included an analysis intended to mirror the GenOMICC study (analysis “A2”), there are currently insufficient cases from other sources available to attempt replication, so we used the broader hospitalised phenotype (analysis “B2”) for replication. We removed signals in the HGI data derived from GenOMICC cases using mathematical subtraction (see Methods) to ensure independence. Using LD clumping to find variants genotyped in both the discovery and replication studies, we required *P* < 0.002 (0.05/25) and concordant direction of effect (Table 2) for replication.

**Table 2:** Replication in a combined data from external studies - combined meta-analysis of HGI freeze 6 B2 and 23andMe. Odds ratios and *P*-values are shown for variants in LD with the lead variant that were genotyped/imputed in both sources. Chromosome, reference and alternate allele correspond to the build hg38. An asterisk (*) next to the HGI and 23andme meta-analysis *P*-value indicates that the lead signal is replicated with *P*-value<0.002 with a concordant direction of effect. Citation lists the first publication of confirmed genome-wide associations with critical illness or (in brackets) any COVID-19 phenotype; in this column, (*) indicates a variant which met genome-wide significance without GenOMICC data in the public latest version of the HGI analysis (B2 v6) but has not been reported yet.

| chr:pos (hg38) | rsid | REF | ALT | OR | $OR_{CI}$ | $P_{val}$ | $OR_{hgi, 23m}$ | $OR_{CI_{hgi, 23m}}$ | $P_{val_{hgi, 23m}}$ | Gene | Citation |
| --- | --- | --- | --- | --- | --- | --- | --- | --- | --- | --- | --- |
| 1:155066988 | rs114301457 | C | T | 2.40 | 1.81-3.18 | $1.51 \times 10^{-9}$ | 1.46 | 1.21-1.77 | 0.00011 * | *EFNA4* | - |
| 1:155175305 | rs7528026 | G | A | 1.39 | 1.24-1.55 | $7.16 \times 10^{-9}$ | 1.14 | 1.07-1.22 | 0.00012 * | *TRIM46* | - |
| 1:155197995 | rs41264915 | A | G | 0.80 | 0.76-0.86 | $3.79 \times 10^{-12}$ | 0.9 | 0.87-0.933 | $1.51 \times 10^{-9}$ * | *THBS3* | - |
| 2:60480453 | rs1123573 | A | G | 0.88 | 0.85-0.92 | $9.85 \times 10^{-10}$ | 0.95 | 0.93-0.97 | 0.000018 * | *BCL11A* | - |
| 3:45796521 | rs2271616 | G | T | 1.26 | 1.19-1.34 | $2.45 \times 10^{-15}$ | 1.11 | 1.07-1.15 | $4.95 \times 10^{-9}$ * | *SLC6A20* | (<sup>2</sup>) |
| 3:45859597 | rs73064425 | C | T | 2.52 | 2.35-2.70 | $2.18 \times 10^{-152}$ | 1.46 | 1.4-1.51 | $1.02 \times 10^{-77}$ * | *LZTFL1* | <sup>4</sup> |
| 3:146517122 | rs343320 | G | A | 1.24 | 1.15-1.33 | $1.52 \times 10^{-8}$ | 1.08 | 1.04-1.13 | 0.00028 * | *PLSCR1* | - |
| 5:132441275 | rs10066378 | T | C | 1.20 | 1.13-1.27 | $4.48 \times 10^{-10}$ | 1.05 | 1.02-1.08 | 0.00074 * | *IRF1-AS1* | - |
| 6:32623820 | rs9271609 | T | C | 0.88 | 0.84-0.92 | $1.27 \times 10^{-8}$ | 1 | 0.98-1.03 | 0.89 | *HLA-DQA1* | - |
| 6:41515007 | rs2496644 | A | C | 0.69 | 0.63-0.76 | $7.59 \times 10^{-15}$ | 0.87 | 0.83-0.92 | $3.17 \times 10^{-7}$ * | *LINC01276* | - |
| 9:21206606 | rs28368148 | C | G | 1.74 | 1.45-2.1 | $4.09 \times 10^{-9}$ | 1.21 | 1.07-1.37 | 0.0024 | *IFNA10* | - |
| 11:34482745 | rs61882275 | G | A | 0.87 | 0.84-0.91 | $1.62 \times 10^{-11}$ | 0.93 | 0.91-0.95 | $1.9 \times 10^{-10}$ * | *ELF5* | * |
| 12:132479205 | rs4883585 | G | A | 1.13 | 1.09-1.18 | $1.12 \times 10^{-9}$ | 1.04 | 1.02-1.06 | 0.00047 * | *FBRSL1* | - |
| 13:112889041 | rs9577175 | C | T | 1.18 | 1.13-1.23 | $1.61 \times 10^{-12}$ | 1.07 | 1.04-1.09 | $1.29 \times 10^{-6}$ * | *ATP11A* | - |
| 15:93046840 | rs4424872 | T | A | 0.64 | 0.53-0.76 | $1.99 \times 10^{-6}$ | - | - | - | *RGMA* | - |
| 16:89196249 | rs117169628 | G | A | 1.18 | 1.12-1.25 | $6.04 \times 10^{-9}$ | 1.1 | 1.07-1.14 | $6.57 \times 10^{-9}$ * | *SLC22A31* | - |
| 17:46152620 | rs2532300 | T | C | 0.87 | 0.82-0.91 | $1.4 \times 10^{-8}$ | 0.92 | 0.89-0.94 | $2.49 \times 10^{-9}$ * | *KANSL1* | <sup>9</sup> |
| 17:49863260 | rs3848456 | C | A | 1.42 | 1.27-1.58 | $1.47 \times 10^{-10}$ | 1.15 | 1.09-1.21 | $1.34 \times 10^{-7}$ * | . | <sup>2</sup> |
| 19:4717660 | rs12610495 | A | G | 1.32 | 1.27-1.38 | $6.44 \times 10^{-39}$ | 1.11 | 1.09-1.14 | $5.74 \times 10^{-19}$ * | *DPP9* | <sup>1</sup> |
| 19:10305768 | rs73510898 | G | A | 1.24 | 1.16-1.33 | $1.47 \times 10^{-9}$ | 1.08 | 1.04-1.12 | 0.00016 * | *ZGLP1* | - |
| 19:10352442 | rs34536443 | G | C | 1.50 | 1.37-1.66 | $4.22 \times 10^{-17}$ | 1.22 | 1.15-1.29 | $4.06 \times 10^{-11}$ * | *TYK2* | <sup>1</sup> |
| 19:48697960 | rs368565 | C | T | 1.13 | 1.09-1.18 | $3.74 \times 10^{-10}$ | 1.04 | 1.02-1.06 | 0.00087 * | *FUT2* | - |
| 21:33230000 | rs17860115 | C | A | 1.26 | 1.21-1.31 | $6.28 \times 10^{-28}$ | 1.11 | 1.08-1.13 | $1.77 \times 10^{-18}$ * | *IFNAR2* | <sup>1</sup> |
| 21:33287378 | rs8178521 | C | T | 1.17 | 1.12-1.22 | $4.23 \times 10^{-12}$ | 1.06 | 1.03-1.09 | $8.02 \times 10^{-6}$ * | *IL10RB* | - |
| 21:33914436 | rs12626438 | A | G | 1.22 | 1.14-1.31 | $1.78 \times 10^{-8}$ | 1.1 | 1.06-1.14 | $2.33 \times 10^{-7}$ * | *LINC00649* | - |

We replicated 22 of the 25 significant associations identified in the population specific and/or trans-ancestry GWAS. Two of the three loci not replicated correspond to rare alleles that may not be well represented in the replication datasets which are dominated by SNP genotyping data. Although not replicated, for rs28368148 (9:21206606:C:G, *IFNA10*) we observed both a consistent direction of effect and odds ratio. The third locus is within the human leukocyte antigen (HLA) locus (see below).

We inferred credible sets of variants using Bayesian fine-mapping with susieR^15^, by analysing the GWAS summaries of 17 3Mbp regions that were flanking groups of lead signals. We obtained 22 independent credible sets of variants for EUR and one for SAS that each had posterior inclusion probability > 0.95.

### Gene burden testing

To assess the contribution of rare variants to critical illness, we performed gene-based analysis using SKAT-O as implemented in SAIGE-GENE^16^, using a subset of 12,982 individuals from our cohort (7,491 individuals with critical COVID-19 and 5,391 controls) for which the genome sequencing data were processed with the same alignment and variant calling pipeline. We tested the burden of rare (MAF<0.5%) variants considering the predicted variant consequence type. We assessed burden using a strict definition for damaging variants (high-confidence loss-of-function (pLoF) variants as identified by LOFTEE^17^) and a lenient definition (pLoF plus missense variants with CADD ≥ 10)^14^, but found no significant associations at a gene-wide significance level. All individual rare variants included in the tests had *P*-values >10^−5^.

We then further examined the association with 13 genes involved in the regulation of type I and III interferon immunity that were implicated in critical COVID-19 pneumonia^3^ but, as with other recent studies^10^, we did not find any significant gene burden test associations (tests for all genes had *P*-value>0.05, Supplementary File AVTsuppinfo.xlsx). We also did not replicate the reported association^10^ for the toll-like receptor 7 (*TLR7*) gene.

### Transcriptome-wide association study

In order to infer the effect of genetically-determined variation in gene expression on disease susceptibility, we performed a transcriptome-wide association study (TWAS) using gene expression data (GTExv8) for two disease-relevant tissues, lung and whole blood. We found 14 genes with significant association between predicted expression and critical COVID-19 in the lung and 6 in whole blood analyses (Supplementary File: TWAS.xlsx). To increase statistical power using eQTLs from multiple tissues, we performed a TWAS meta-analysis using all available tissues in GTExv8, revealing 51 transcriptome-wide significant genes. Since TWAS uses a composite signal derived from multiple eQTLs, we used colocalisation to find specific eQTLs in whole blood (eqtlGen and GTExv8) and lung (GTExv8^18^) which share the same signal with GWAS (EUR) associations. We found 16 genes which significantly colocalise in at least one of the studied tissues, shown in Figure 2.

**Figure 2:**
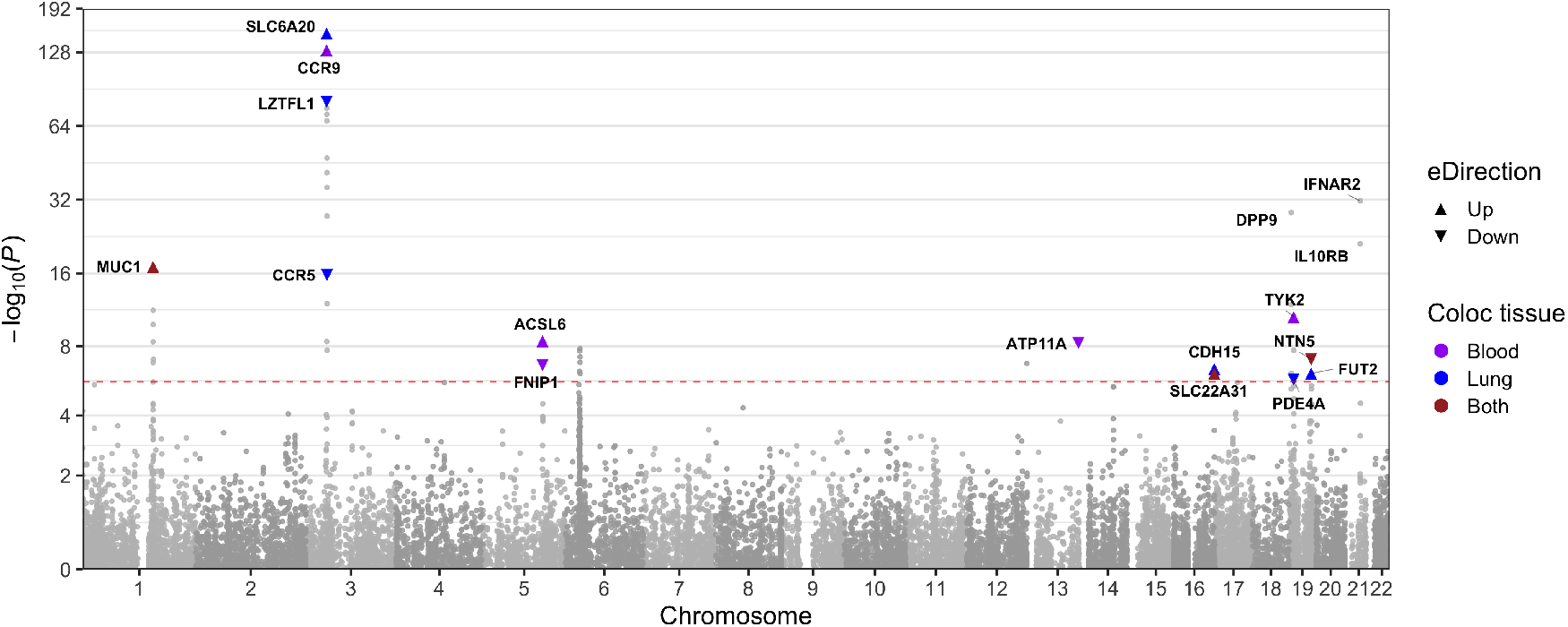
Gene-level Manhattan plot showing results from TWAS meta-analysis and highlighting genes that colocalise with GWAS signals or have strong metaTWAS associations. Highlighting color is different for lung and blood tissue data that were used for colocalisation. Arrows show direction of change in gene expression associated with an increased disease risk. Red dashed line shows significance threshold at *P* < 2.3 × 10^−6^.

We repeated the TWAS analysis using models of intron excision rate from GTExv8 to obtain splicing TWAS. We found 40 signals in lung, affecting 16 genes and 20 signals in whole blood which affect 9 genes. In a meta-analysis of splicing TWAS using all GTExv8 tissues, we found 91 significant introns in a total of 33 genes. Using GTExv8 lung and whole blood sqtls to find colocalising signals with splicing TWAS significant results, we found 11 genes with colocalising splicing signals (Supplementary File: TWAS.xlsx).

### HLA region

To investigate the contribution of specific HLA alleles to the observed association in the HLA region, we imputed HLA alleles at a four digit (two-field) level using HIBAG^19^. The only allele that reached genome-wide significance was HLA-DRB1*04:01 (*OR* = 0.80, 95%*CI* = 0.75 − 0.86, *P* = 1.6 × 10^−10^ in EUR), which has a stronger *P*-value than the lead SNP in the region (*OR* : 0.88, 95%*CIs* : 0.84 − 0.92, *P* = 3.3 × 10^−9^ in EUR) and is a better fit to the data (*AIC*_*DRB*1*04:01_ = 30241.34, *AIC*_*leadSNP*_ = 30252.93). Results are shown in supplementary figure 25.

## Discussion

We report 22 replicated genetic associations with life-threatening COVID-19, and 3 additional loci, discovered in only 7,491 cases. This demonstrates the efficiency of the design of the GenOMICC study, which is an open-source international research programme^20^ focusing on critically-ill patients with infectious disease and other critical illness phenotypes (https://genomicc.org). By using whole genome sequencing we were able to detect multiple distinct signals with high confidence for several of the associated loci, in some cases implicating different biological mechanisms.

Several variants associated with life-threatening disease are linked to interferon signalling. A coding variant in a ligand, *IFNA10A*, and reduced expression of its receptor *IFNAR2* (Figure 2), were associated with critical COVID-19. The narrow failure of replication for the *IFNA10* variant (rs28368148, replication *P* = 0.00243, significance threshold *P* < 0.002) may be due to limited power in the replication cohort. The lead variant in *TYK2* in whole genome sequencing is a well-studied protein-coding variant with reduced phosphorylation activity, consistent with that reported recently,^2^ but associated with significantly increased *TYK2* expression (Figure 2, Methods). Fine mapping reveals a significant critical illness association with an independent missense variant in *IL10RB*, a receptor for Type III (lambda) interferons (rs8178521, Trp164Cys, Table 1). Overall, variants predicted to be associated with reduction in interferon signalling are associated with critical disease. Importantly, systemic administration of interferon in a large clinical trial, albeit late in disease, did not reduce mortality.^21^

Phospholipid scramblase 1 (*PLSCR1* ; chr3:146517122:G:A) functions as a nuclear signal for the antiviral effect of interferon,^22^ and has been shown to control replication of other RNA viruses including vesicular stomatitis virus, encephalomyocarditis virus and Influenza A virus.^23;22^ The risk allele at the lead variant (chr3:146517122:G:A, rs343320) encodes a substitution, H262Y, which is predicted to disrupt the non-canonical nuclear localisation signal^24^ by eliminating a hydrogen bond with importin (Figure 3). Deletion of this nuclear localisation signal has been shown to prevent neutrophil maturation.^25^ Although *PLSCR1* is strongly up-regulated when membrane lipid asymmetry is lost (see below), it may not act directly on this process.^26^

**Figure 3:**
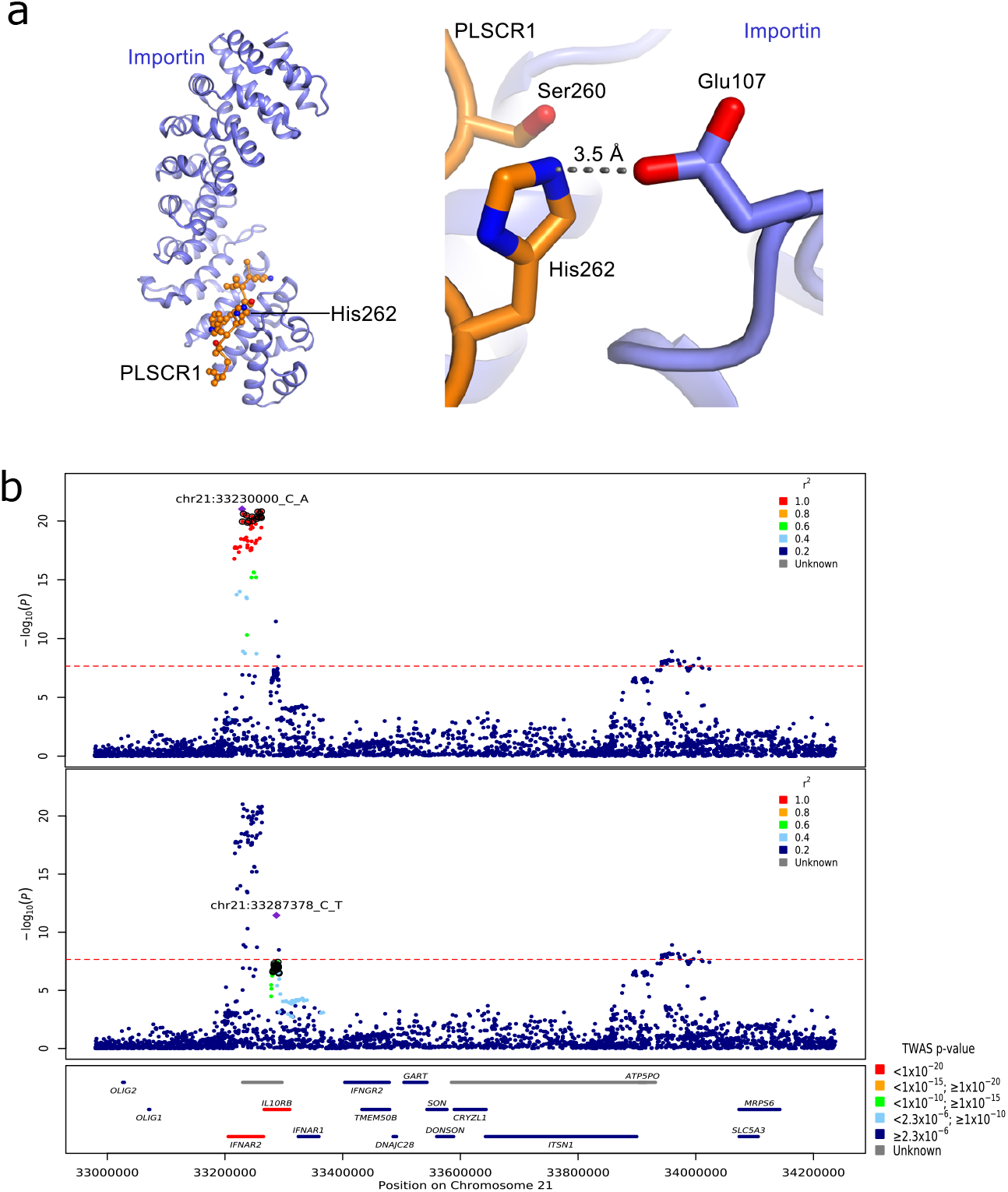
(a) Predicted structural consequences of lead variant at *PLSCR1*. Left panel shows the crystal structure of PLSCR1 nuclear localization signal (orange, Gly257–Ile266, numbering correspond to UniProt entry O15162) in complex with Importin *α* (blue), Protein Data Bank (PDB) ID 1Y2A. Side chains of PLSCR1 are shown as connected spheres with carbon atoms coloured in orange, nitrogens in blue and oxygens in red. Hydrogen atoms were not determined at this resolution (2.20 Å) and are not shown. Right panel: a closeup view showing side chains of PLSCR1 Ser260, His262 and Importin Glu107 as sticks. Distance (in Å) between selected atoms (PLSCR1 His262 *Nϵ*2 and Importin Glu107 carboxyl O) is indicated. A hydrogen bond between PLSCR1 His262 and Importin Glu107 is indicated with a dashed line. The risk variant is predicted to eliminate this bond, disrupting nuclear import, an essential step for effect on antiviral signalling^22^ and neutrophil maturation.^25^ (b) Regional detail showing fine-mapping to separate two adjacent independent signals. Top two panels: variants in linkage disequilibrium with the lead variants shown. The loci that are included in two independent credible sets are displayed with black outline circles. Bottom panel: locations of protein-coding genes, coloured by TWAS *P*-value.

We report significant associations in several genes implicated in B-cell lymphopoesis and differentiation of myeloid cells. *BCL11A* is essential in B- and T-lymphopoiesis^27^ and promotes plasmacytoid dendritic cell differentiation.^28^ *TAC4*, reported previously,^2^ encodes a regulator of B-cell lymphopoesis^29^ and antibody production,^30^ and promotes survival of dendritic cells.^31^ Finally, although the strongest fine mapping signal at 5q31.1 (chr5:131995059:C:T, rs56162149) is in an intron of *ACSL6* (locus, p), the credible set includes a missense variant in *CSF2* of uncertain significance (chr5:132075767:T:C). *CSF2* encodes granulocyte-macrophage colony stimulating factor, a key differentiation factor in the mononuclear phagocyte system which is strongly up-regulated in critical COVID-19,^32^ and is already under investigation as a target for therapy.^33^

Several new genetic associations implicate genes known to be involved in lung disease. The second variant in the credible set at 13q14 (chr13:112882313:A:G, rs1278769, in *ATP11A*), has been reported as a lead variant for idiopathic pulmonary fibrosis.^34^ *ATP11A* encodes a flippase which maintains the asymmetric distribution of phospholipids in cell membranes;^35^ disruption of this asymmetry is a phagocytic signal on apoptotic cells, and is required for platelet activation.^36;37^ TWAS and colocalisation demonstrate that genetic variants predicted to decrease expression of *ATP11A* in lung are associated with critical illness. A combination of fine mapping, colocalisation with eQTL signals (GTEx and eQTLgen) and TWAS results provide evidence in support of *MUC1* as the mediator of the association with rs41264915 (Table 1). This may indicate an important role for mucins in the development of critical illness in COVID-19. The direction of effect (Figure 2) suggests that agents that reduce *MUC1* expression, and by extension its abundance, may be a therapeutic option. Finally, the association on 11p13 (rs61882275) includes GTEx eQTL for the lung fibroblast transcription factor *ELF5* in lung tissue, and the gene encoding the antioxidant enzyme catalase (*CAT*) in whole blood with evidence of colocalisation in both signals (supplementary material: TWAS.xlsx).^18^ The protective allele at this locus is weakly associated with reduced lung function in a previous GWAS.^38^

*FUT2* encodes alpha-(1,2)fucosyltransferase, which controls the secretion of ABO blood type glycans into body fluids and expression on epithelial surfaces. An association with critical COVID-19 was reported previously in a candidate gene association study by Mankelow et al.^39^ The credible set for the *FUT2* locus includes rs492602 (chr19:48703160:A:G) which is linked to a stop codon gain mutation (chr19:48703417:G:A), leading to the well-described non-secretor phenotype in homozygotes.^40;41^ We show that the stop-gain, non-secretor allele is protective against life-threatening COVID-19. The protective variant in our study has been previously reported to protect against other viruses (rotavirus,^42^ mumps and common colds^43^), to enhance antibody responses to polyomavirus BK^44^ and to increase susceptibility to infection with some encapsulated bacteria.^45^

### Limitations

In contrast to microarray genotyping, whole genome sequencing is rapidly evolving and a relatively new technology for genome-wide association studies, with relatively few sources of population controls. We used selected controls from the 100,000 genomes project, sequenced on a different platform (illumina HiSeqX) from the cases (illumina NovaSeq6000)(Supplementary Table 1). To minimise the risk of false positive associations arising due to sequencing or genotyping errors, we required all significant associations to be supported by local variants in linkage disequilibrium, which may be excessively stringent (see Methods). Although this approach may remove some true associations, our priority is to maximise confidence in the reported signals. Of 25 variants meeting this requirement, 22 are replicated in an independent study, and the remaining 3 may well be true associations that have failed due to a lack of coverage or power in the replication dataset.

The design of our study incorporates genetic signals for every stage in the disease progression into a single phenotype. This includes exposure, viral replication, inflammatory lung injury and hypoxaemic respiratory failure. Although we can have considerable confidence that the replicated associations with critical COVID-19 we report are robust, we cannot determine at which stage in the disease process, or in which tissue, the relevant biological mechanisms are active, which can have therapeutic implications.

## Conclusions

The genetic associations here implicate new biological mechanisms underlying the development of life-threatening COVID-19, several of which may be amenable to therapeutic targeting. In the context of the ongoing global pandemic, translation to clinical practice is an urgent priority. As with our previous work, large-scale randomised trials are essential before translating our findings into clinical practice.

## Supporting information

Aggregate variant testing additional results tables

GWAS additional results tables

HLA analysis additional results tables

TWAS additional results tables

## Data Availability

Summary statistics will be shared openly with international collaborators to accelerate discovery. Data can be obtained from https://genomicc.org/data/
Individual-level data will be available in the UK Outbreak Analysis Platform at the University of Edinburgh and through the Genomics England research environment.

## Acknowledgements

We thank the patients and their loved ones who volunteered to contribute to this study at one of the most difficult times in their lives, and the research staff in every intensive care unit who recruited patients at personal risk during the most extreme conditions we have ever witnessed in UK hospitals.

GenOMICC was funded by the Department of Health and Social Care (DHSC), LifeArc, the Medical Research Council, UKRI, Sepsis Research (the Fiona Elizabeth Agnew Trust), the Intensive Care Society, a Wellcome-Beit Prize award to J. K. Baillie (Wellcome Trust 103258/Z/13/A) and a BBSRC Institute Program Support Grant to the Roslin Institute (BBS/E/D/20002172, BBS/E/D/10002070 and BBS/E/D/30002275). Whole-genome sequencing was performed by Illumina at the NHS Genomic Sequencing Centre in partnership and was overseen by Genomics England. We would like to thank all at Genomics England who have contributed to the supporting the processing of the sequencing and clinical data. We thank DHSC, the Medical Research Council, UKRI, LifeArc, Genomics England Ltd and Illumina Inc for funding sequencing. Genomics England and the 100,000 Genomes Project was funded by the National Institute for Health Research, the Wellcome Trust, the Medical Research Council, Cancer Research UK, the Department of Health and Social Care and NHS England. We are grateful for the support from Professor Dame Sue Hill and the team in NHS England and the 13 Genomic Medicine Centres that successfully delivered the 100,000 Genomes Project which provide the control sequences for this study. We thank the participants of the 100,000 Genomes Project who made this study possible and the Genomics England Participant Panel for their strategic advice, involvement and engagement. We acknowledge NHS Digital, Public Health England and the Intensive Care National Audit and Research Centre who provided life course longitudinal clinical data on the participants. This work forms part of the portfolio of research of the NIHR Biomedical Research Centre at Barts. Mark Caulfield is an NIHR Senior Investigator. This study owes a great deal to the National Institute of Healthcare Research Clinical Research Network (NIHR CRN) and the Chief Scientist Office (Scotland), who facilitate recruitment into research studies in NHS hospitals, and to the global ISARIC and InFACT consortia.

The views expressed are those of the authors and not necessarily those of the DHSC, DID, NIHR, MRC, Wellcome Trust or PHE.

The Genotype-Tissue Expression (GTEx) Project was supported by the Common Fund of the Office of the Director of the National Institutes of Health, and by NCI, NHGRI, NHLBI, NIDA, NIMH, and NINDS. The data used for the analyses described in this manuscript were obtained from the GTEx Portal on August 22nd, 2021 (GTEx Analysis Release V8 (dbGaP Accession phs000424.v8.p2).

## Data availability

Summary statistics will be shared openly with international collaborators to accelerate discovery. Data can be obtained from genomicc.org/data

Individual-level data will be available in the UK Outbreak Analysis Platform at the University of Edinburgh and through the Genomics England research environment.

## Contributions

AK, EP-C, KR, AS, CAO, SW, TM, KSE, BW, DR, LK, MZ, NP, ADB, YW, JY, SC, LMo, AL and JKB contributed to data analysis. AK, EP-C, KR, AS, CAO, SW, CDR, JM, AR, SC, LMo and AL contributed to bioinformatics. AK, EP-C, KR, CDR, JM, DM, AN, MGS, SC, LMo, MJC and JKB contributed to writing and reviewing the manuscript. EP-C, KR, KM, SK, AF, LM, KRo, CPP, VV, JFW, SC, AL, MJC and JKB contributed to design. SW, FG, WO, PG and SD contributed to project management. FG, WO, KM, SK, PG, SD, DM, AN, MGS, SS, JK, TAF, MS-H, CS, CH, PH, DMc, HM, PJO, PE, TW, AF, LM, KRo, CPP, RHS, SC and AL contributed to oversight. FG, WO, FM-C and JKB contributed to ethics and governance. KM, ASi, AF and LM contributed to sample handling and sequencing. and ASi contributed to data collection. and TZ contributed to sample handing. TZ and GE contributed to sequencing. and LT contributed to recruitment of controls. GC, PA, KRo and AL contributed to clinical data management. KRo, CPP, SC and JKB contributed to conception. KRo, CPP, VV and JFW contributed to reviewing the manuscript. MJC and JKB contributed to scientific leadership.

## Conflict of interest

All authors declare that they have no conflicts of interest relating to this work.

Genomics England Ltd is a wholly owned Department of Health and Social Care company created in 2013 to work with the NHS to introduce advanced genomic technologies and analytics into healthcare. All Genomics England affiliated authors are, or were, salaried by Genomics England during this programme.

## Materials and Methods

### Ethics

GenOMICC was both approved by the following research ethics committees: Scotland “A” Research Ethics Committee, 15/SS/0110; Coventry and Warwickshire Research Ethics Committtee (England, Wales and Northern Ireland), 19/WM/0247). Current and previous versions of the study protocol are available at genomicc.org/protocol. All participants gave informed consent.

### Recruitment of cases

Patients recruited to the GenOMICC study (genomicc.org) had confirmed COVID-19 according to local clinical testing and were deemed, in the view of the treating clinician, to require continuous cardiorespiratory monitoring. In UK practice this kind of monitoring is undertaken in high-dependency or intensive care units. This study was approved by research ethics committees in the recruiting countries (Scotland 15/SS/0110, England, Wales and Northern Ireland: 19/WM/0247). Current and previous versions of the study protocol are available at genomicc.org/protocol. All participants gave informed consent.

### Recruitment of controls

#### Mild/asymptomatic controls

Participants were recruited to the mild COVID-19 cohort on the basis of having experienced mild (non-hospitalised) or asymptomatic COVID-19. Participants volunteered to take part in the study via a microsite and were required to self-report the details of a positive COVID-19 test. Volunteers were prioritised for genome sequencing based on demographic matching with the critical COVID-19 cohort considering self-reported ancestry, sex, age and location within the UK. We refer to this cohort as the covid-mild cohort.

#### 100,000 Genomes project controls

Participants were enrolled in the 100,000 Genomes Project from families with a broad range of rare diseases, cancers and infection by 13 regional NHS Genomic Medicine Centres across England and in Northern Ireland, Scotland and Wales. For this analysis, participants for whom a positive SARS-CoV-2 test had been recorded as of March, 2021 were not included due to uncertainty in the severity of COVID-19 symptoms. Only participants for whom genome sequencing was performed from blood derived DNA were included and participants with haematological malignancies were excluded to avoid potential tumour contamination.

#### DNA extraction

For severe COVID-19 cases and mild cohort controls, DNA was extracted from whole blood either manually using Nucleon Kit (Cytiva) and re-suspended in 1 ml TE buffer pH 7.5 (10mM Tris-Cl pH 7.5, 1mM EDTA pH 8.0), or automated on the Chemagic 360 platform using Chemagic DNA blood kit (Perkin Elmer) and re-suspended in 400*µ*L Elution Buffer. The yield of the DNA was measured using Qubit and normalised to 50ng/*µ*l before sequencing.

#### WGS sequencing

For all three cohorts, DNA was extracted from whole-blood using standard protocols. Sequencing libraries were generating using the Illumina TruSeq DNA PCR-Free High Throughput Sample Preparation kit and sequenced with 150bp paired-end reads in a single lane of an Illumina Hiseq X instrument (for 100,000 Genomes Project samples) or NovaSeq instrument (for the COVID-19 critical and mild cohorts).

#### Sequencing data QC

All genome sequencing data were required to meet minimum quality metrics and quality control measures were applied for all genomes as part of the bioinformatics pipeline. The minimum data requirements for all genomes were > 85×10^−9^ bases with *Q* ≥ 30 and ≥ 95% of the autosomal genome covered at ≥ 15*x* calculated from reads with mapping quality > 10 after removing duplicate reads and overlapping bases, after adaptor and quality trimming. Assessment of germline cross-sample contamination was performed using VerifyBamID and samples with > 3% contamination were excluded. Sex checks were performed to confirm that the sex reported for a participant was concordant with the sex inferred from the genomic data.

### WGS Alignment and variant calling

#### COVID-19 cohorts

For the critical and mild COVID-19 cohorts, sequencing data alignment and variant calling was performed with Genomics England pipeline 2.0 which uses the DRAGEN software (v3.2.22). Alignment was performed to genome reference GRCh38 including decoy contigs and alternate haplotypes (ALT contigs), with ALT-aware mapping and variant calling to improve specificity.

#### 100,000 Genome Project cohort (100K-genomes)

All genomes from the 100,000 Genomes Project cohort were analysed with the Illumina North Star Version 4 Whole Genome Sequencing Workflow (NSV4, version 2.6.53.23); which is comprised of the iSAAC Aligner (version 03.16.02.19) and Starling Small Variant Caller (version 2.4.7). Samples were aligned to the Homo Sapiens NCBI GRCh38 assembly with decoys.

A subset of the genomes from the Cancer program of the 100,000 Genomes Project were reprocessed (alignment and variants calling) using the same pipeline used for the COVID-19 cohorts (DRAGEN v3.2.22) for equity of alignment and variant calling.

#### Aggregation

Aggregation was conducted separately for the samples analysed with Genomics England pipeline 2.0 (severe-cohort, mild-cohort, cancer-realigned-100K), and those analysed with the Illumina North Star Version 4 pipeline (100K-Genomes).

For the first three, the WGS data were aggregated from single sample gVCF files to multi-sample VCF files using GVCFGenotyper (GG) v3.8.1, which accepts gVCF files generated via the DRAGEN pipeline as input. GG outputs multi-allelic variants (several ALT variants per position on the same row), and for downstream analyses the output was decomposed to bi-allelic variants per row using software vt v0.57721. We refer to the aggregate as aggCOVID_vX, where X is the specific freeze. The analysis in this manuscript uses data from freeze v4.2 and the respective aggregate is referred to as aggCOVID_v4.2.

Aggregation for the 100K-Genomes cohort was performed using Illumina’s gvcfgenotyper v2019.02.26, merged with bcftools v1.10.2 and normalised with vt v0.57721.

#### Sample Quality Control (QC)

Samples that failed any of the following four BAM-level QC filters: freemix contamination (>3%), mean autosomal coverage (<25X), percent mapped reads (<90%), and percent chimeric reads (>5%) were excluded from the analysis.

Additionally, a set of VCF-level QC filters were applied post-aggregation on all autosomal bi-allelic SNVs (akin to gnomAD v3.1^17^). Samples were filtered out based on the residuals of eleven QC metrics (calculated using bcftools) after regressing out the effects of sequencing platform and the first three ancestry assignment principal components (including all linear, quadratic, and interaction terms) taken from the sample projections onto the SNP loadings from the individuals of 1000 Genomes Project phase 3 (1KGP3). Samples were removed that were four median absolute deviations (MADs) above or below the median for the following metrics: ratio heterozygous-homozygous, ratio insertions-deletions, ratio transitions-transversions, total deletions, total insertions, total heterozygous snps, total homozygous snps, total transitions, total transversions. For the number of total singletons (snps), samples were removed that were more than 8 MADs above the median. For the ratio of heterozygous to homozygous alternate snps, samples were removed that were more than 4 MADs above the median.

After quality control, 79,803 individuals were included in the analysis with the breakdown according to cohort shown in Supplementary Table 2.

#### Selection of high-quality (HQ) independent SNPs

We selected high-quality independent variants for inferring kinship coefficients, performing PCA, assigning ancestry and for the conditioning on the Genetic Relatedness matrix by the logistic mixed model of SAIGE and SAIGE-GENE. To avoid capturing platform and/or analysis pipeline effects for these analyses, we performed very stringent variant QC as described below.

#### HQ common SNPs

We started with autosomal, bi-allelic SNPs which had frequency > 5% in aggV2 (100K participant aggregate) and in the 1KGP3. We then restricted to variants that had missingness <1%, median genotype quality QC>30, median depth (DP) >=30 and >= 90% of heterozygote genotypes passing an ABratio binomial test with *P*-value > 10^−2^ for aggV2 participants. We also excluded variants in complex regions from the list available in, and variants where the ref/alt combination was CG or AT (C/G, G/C, A/T, T/A). We also removed all SNPs which were out of Hardy Weinberg Equilibrium (HWE) in any of the AFR, EAS, EUR or SAS super-populations of aggV2, with a *P*-value cutoff of pHWE < 10^−5^. We then LD-pruned using plink v1.9 with an *r*^2^ = 0.1 and in 500kb windows. This resulted in a total of 63,523 high-quality sites from aggV2.

We then extracted these high-quality sites from the aggCOVID_v4.2 aggregate and further applied variant quality filters (missingness <1%, median QC>30, median depth >=30 and >= 90% of heterozygote genotypes passing an ABratio binomial test with *P*-value > 10^−2^), per batch of sequencing platform (i.e, HiseqX, NovaSeq6000).

After applying variant filters in aggV2 and aggCOVID_v4.2, we merged the genomic data from the two aggregates for the intersection of the variants which resulted in a final total of 58,925 sites.

#### HQ rare SNPs

We selected high-quality rare (MAF< 0.005) bi-allelic SNPs to be used with SAIGE for aggregate variant testing analysis. To create this set, we applied the same variant QC procedure as with the common variants: We selected variants that had missingness <1%, median QC>30, median depth >=30 and >= 90% of heterozygote genotypes passing an ABratio binomial test with *P* –value > 10^−2^ per batch of sequencing and genotyping platform (i.e, HiSeq+NSV4, HiSeq+Pipeline 2.0, NovaSeq+Pipeline 2.0). We then subsetted those to the following groups of MAC/MAF categories: MAC 1, 2, 3, 4, 5, 6-10, 11-20, MAC 20 - MAF 0.001, MAF 0.001 - 0.005.

### Relatedness, ancestry and principal components

#### Kinship

We calculated kinship coefficients among all pairs of samples using software plink2 and its implementation of the KING robust algorithm. We used a kinship cutoff < 0.0442 to select unrelated individuals with argument “–king-cutoff”.

#### Genetic Ancestry Prediction

To infer the ancestry of each individual we performed principal components analysis (PCA) on unrelated 1KGP3 individuals with GCTA v1.93.1_beta software using HQ common SNPs and inferred the first 20 PCs. We calculated loadings for each SNP which we used to project aggV2 and aggCOVID_v4.2 individuals onto the 1KGP3 PCs. We then trained a random forest algorithm from R-package randomForest with the first 10 1KGP3 PCs as features and the super-population ancestry of each individual as labels. These were ‘AFR’ for individuals of African ancestry, ‘AMR’ for individuals of American ancestry, ‘EAS’ for individuals of East Asian ancestry, ‘EUR’ for individuals of European ancestry, and ‘SAS’ for individuals of South Asian ancestry. We used 500 trees for the training. We then used the trained model to assign probability of belonging to a certain super-population class for each individual in our cohorts. We assigned individuals to a super-population when class probability >=0.8. Individuals for which no class had probability >=0.8 were labelled as “unassigned” and were not included in the analyses.

#### Principal component analysis

After labelling each individual with predicted genetic ancestry, we calculated ancestry-specific PCs using GCTA v1.93.1_beta, *i*.*e*.. We computed 20 PCs for each of the ancestries that were used in the association analyses (AFR, EAS, EUR, and SAS).

#### Variant Quality Control

Variant QC was performed to ensure high quality of variants and to minimise batch effects due to using samples from different sequencing platforms (NovaSeq6000 and HiseqX) and different variant callers (Strelka2 and DRAGEN). We first masked low-quality genotypes setting them to missing, merged aggregate files and then performed additional variant quality control separately for the two major types of association analyses, GWAS and AVT, which concerned common and rare variants, respectively.

#### Masking

Prior to any analysis we masked low quality genotypes using bcftools setGT module. Genotypes with DP<10, GQ<20, and heterozygote genotypes failing an AB-ratio binomial test with *P*-value < 10^−3^ were set to missing.

We then converted the masked VCF files to plink and bgen format using plink v.2.0.

#### Merging of aggregate samples

Merging of aggV2 and aggCOVID_v4.2 samples was done using plink files with masked genotypes and the merge function of plink v.1.9.^46^ for variants that were found in both aggregates.

### GWAS analyses

#### Variant QC

We restricted all GWAS analyses to common variants applying the following filters using plink v1.9: MAF > 0 in both cases and controls, MAF> 0.5% and MAC >20, missingness < 2%, Differential missingness between cases and controls, mid-*P*-value < 10^−5^, HWE deviations on unrelated controls, mid-*P*-value < 10^−6^, Multi-allelic variants were additionally required to have MAF > 0.1% in both aggV2 and aggCOVID_v4.2.

#### Control-control QC filter

100K aggV2 samples that were aligned and genotype called with the Illumina North Star Version 4 pipeline represented the majority of control samples in our GWAS analyses, whereas all of the cases were aligned and called with Genomics England pipeline 2.0 (Supplementary Table 1). Therefore, the alignment and genotyping pipelines partially match the case/control status which necessitates additional filtering for adjusting for between-pipeline differences in alignment and variant calling. To control for potential batch effects, we used the overlap of 3,954 samples from the Genomics England 100K participants that were aligned and called with both pipelines. For each variant, we computed and compared between platforms the inferred allele frequency for the population samples. We then filtered out all variants that had > 1% relative difference in allele frequency between platforms. The relative difference was computed on a per-population basis for EUR (n=3,157), SAS (n=373), AFR (n=354) and EAS (n=81).

#### Model

We used a 2-step logistic mixed model regression approach as implemented in SAIGE v0.44.5 for single variant association analyses. In step 1, SAIGE fits the null mixed model and covariates. In step 2, single variant association tests are performed with the saddlepoint approximation (SPA) correction to calibrate unbalanced case-control ratios. We used the HQ common variant sites for fitting the null model and *sex, age, age*^2^, *age* * *sex* and 20 principal components as covariates in step 1. The principal components were computed separately by predicted genetic ancestry (i.e, EUR-specific, AFR-specific, etc.), to capture subtle structure effects.

#### Analyses

All analyses were done on unrelated individuals with pairwise kinship coefficient < 0.0442. We conducted GWAS analyses per genetic ancestry, for all populations for which we had >100 cases and >100 controls (AFR, EAS, EUR, and SAS).

#### Multiple testing correction

As our study is testing variants that were directly sequenced by WGS and not imputed, we calculated the *P*-value significance threshold by estimating the effective number of tests. After selecting the final filtered set of tested variants for each population, we LD-pruned in a window of 250Kb and *r*^2^ = 0.8 with plink 1.9. We then computed the Bonferroni-corrected *P*-value threshold as 0.05 divided by the number of LD-pruned variants. The *P*-value thresholds that were used for declaring statistical significance are given in Supplementary Table 3.

#### LD-clumping

We used plink1.9 to do clumping of variants that were genome-wide significant for each analysis with *P* 1 set to per-population *P*-value from table X, *P* 2 = 0.01, clump distance 1500Mb and *r*^2^ = 0.1.

#### Conditional analysis

To find the set of independent variants in the per-population analyses, we performed a step-wise conditional analysis with the GWAS summary statistics for each population using GTCA 1.9.3 –cojo-slct function. The parameters for the function were *pval* = 2.2 × 10^−8^, a distance of 10,000 kb and a colinear threshold of 0.9^47^.

#### Fine-mapping

We performed fine-mapping for genome-wide significant signals using Rpackage SusieR v0.11.42^48^. For each genome-wide significant variant locus, we selected the variants 1.5 Mbp on each side and computed the correlation matrix among them with plink v1.9. We then run the susieR summary-statistics based function susie_rss and provided the summary z-scores from SAIGE (i.e, effect size divided by its standard error) and the correlation matrix computed with the same samples that were used for the corresponding GWAS. We required coverage >0.95 for each identified credible set and minimum and median correlation coefficients (purity) of r=0.1 and 0.5, respectively.

#### Functional annotation of credible sets

We annotated all variants included in each credible set identified by SusieR using VEP v99. We also selected the worst consequence across transcripts using bcftools +split-vep -s worst. We also ranked each variant within each credible set according to the predicted consequence and the ranking was based on the table provided by Ensembl: https://www.ensembl.org/info/genome/variation/prediction/predicted_data.html.

#### Trans-ancestry meta-analysis

We performed a meta-analysis across all ancestries using a inverse-variance weighted method and control for population stratification for each separate analysis in the METAL software^13^. The meta-analysed variants were filtered for variants with heterogeneity *P*-value *p* < 2.22 × 10^−8^ and variants that are not present in at least half of the individuals. We used the meta R package to plot forest plots of the clumped trans-ancestry meta-analysis variants^49^.

#### LD-based validation of lead GWAS signals

In order to quantify the support for genome-wide significant signals from nearby variants in LD, we assessed the internal consistency of GWAS results of the lead variants and their surroundings. To this end, we compared observed z-scores at lead variants with the expected z-scores based on those observed at neighbouring variants. Specifically, we computed the observed z-score for a variant *i* as 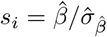 and, following the approach of^50^, the imputed z-score at a target variant *t* as

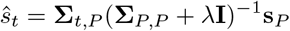

where **s**_*P*_ are the observed z-scores at a set *P* of predictor variants, **Σ**_*x,y*_ is the empirical correlation matrix of dosage coded genotypes computed on the GWAS sample between the variants in *x* and *y*, and *λ* is a regularization parameter set to 10^−5^. The set *P* of predictor variants consisted of all variants within 100 kb of the target variant with a genotype correlation with the target variant greater than 0.25. This approach is similar to one proposed recently by Chen et al.^51^

#### Replication

We used the Host Genetic Initiative (HGI) GWAS meta-analysis round 6 hospitalised COVID vs population (B2 analysis), including all genetic ancestries. In order to remove overlapping signals we performed a mathematical subtraction of the GenOMICC GWAS of European genetic ancestry. The HGI data was downloaded from https://www.covid19hg.org/results/r6/. The subtraction was performed using MetaSubtract package (version 1.60) for R (version 4.0.2) after removing variants with the same genomic position and using the lambda.cohortswith genomic inflation calculated on the GenOMICC summary statistics. Then, we calculated a trans-ancestry meta-analysis for the three ancestries with summary statistics in 23andMe: African, Latino and European using variants that passed the 23andMe ancestry QC, with imputation score > 0.6 and with maf > 0.005. And finally we performed a final meta-analysis of 23andMe and HGI B2 without GenOMICC to create the final replication set. Meta-analysis were performed using METAL^13^, with the inverse-variance weighting method (STDERR mode) and genomic control ON. We considered that a hit was replicating if the direction of effect in the GenOMICC-subtracted HGI summary statistics was the same as in our GWAS, and the *P*-value was significant after Bonferroni correction for the number of attempted replications (*pval* < 0.05*/*25). If the main hit was not present in the HGI-23andMe meta-analysis or if the hit was not replicating we looked for replication in variants in high LD with the top variant (*r*^2^ > 0.9), which helped replicate two regions.

#### Stratified analysis

We also performed sex-specific analysis (male and females separately) as well as analysis stratified by age (*i*.*e*., participants <60 and >=60 years old) for each super-population set. To compare effect of variants within groups for the age and sex stratified analysis we first adjusted the effect and error of each variant for the standard deviation of the trait in each stratified group and then used the following t-statistic, as in previous studies^52;53^

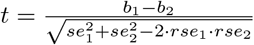

where *b*_1_ is the adjusted effect for group 1, *b*_2_ is the adjusted effect for group 2, *se*_1_ and *se*_2_ are the adjusted standard errors for group 1 and 2 respectively and *r* is the Spearman rank correlation between groups across all genetic variants.

#### HLA Imputation and Association Analysis

HLA types were imputed at two field (4-digit) resolution for all samples within aggV2 and aggCOVID_v4.2 for the following seven loci: HLA-A, HLA-C, HLA-B, HLA-DRB1, HLA-DQA1, HLA-DQB1, and HLA-DPB1 using the HIBAG package in R^19^. At time of writing, HLA types were also imputed for 82% of samples using HLA*LA^54^. Inferred HLA alleles between HIBAG and HLA*LA were >96% identical at 4-digit resolution. HLA association analysis was run under an additive model using SAIGE; in an identical fashion to the SNV GWAS. The multi-sample VCF of aggregated HLA type calls from HIBAG were used as input where any allele call with posterior probability (*T*) < 0.5 were set to missing.

#### Aggregate variant testing (AVT)

Aggregate variant testing on aggCOVID_v4.2 was performed using SKAT-O as implemented in SAIGE-GENE v0.44.5^16^ on all protein-coding genes. Variant and sample QC for the preparation and masking of the aggregate files has been described elsewhere. We further excluded SNPs with differential missingness between cases and controls (mid-P value < 10^−5^) or a site-wide missingness above 5%. Only bi-allelic SNPs with a MAF<0.5% were included.

We filtered the variants to include in the aggregate variant testing by applying two functional annotation filters: A putative loss of function (*pLoF*) filter, where only variants that are annotated by LOFTEE^17^ as high confidence loss of function were included, and a more lenient (*missense*) filter where variants that have a consequence of missense or worse as annotated by VEP, with a CADD_PHRED score of ≥ 10, were also included. All variants were annotated using VEP v99. SAIGE-GENE was run with the same covariates used in the single variant analysis: *sex, age, age*^2^, *age* * *sex* and 20 (population-specific) principal components generated from common variants (MAF ≥ 5%).

We ran the tests separately by genetically predicted ancestry, as well as across all four ancestries as a mega-analysis. We considered a gene-wide significant threshold on the basis of the genes tested per ancestry, correcting for the two masks (*pLoF* and *missense*, Supplementary Table 4).

### Post-GWAS analysis

#### Transcriptome-wide Association Studies (TWAS)

We performed TWAS in the MetaXcan framework and the GTExv8 eQTL and sQTL MASHR-M models available for download in (http://predictdb.org/). We first calculated, using the European summary statistics, individual TWAS for whole blood and lung with the S-PrediXcan function^55;56^. Then we performed a metaTWAS including data from all tissues to increase statistical power using s-MultiXcan^57^. We applied Bonferroni correction to the results in order to choose significant genes and introns for each analysis.

#### Colocalisation analysis

Significant genes from TWAS, splicing TWAS, metaTWAS and splicing metaTWAS, as well as genes where one of the top variants was a significant eQTL or sQTL were selected for a colocalisation analysis using the coloc R package^58^. We chose the lead SNPS from the European ancestry GWAS summary statistics and a region of *±* 200 kb around each SNP to do the colocalisation with the identified genes in the region. GTExv8 whole blood and lung tissue summary statistics and eqtlGen (which has blood eQTL summary statistics for > 30, 000 individuals) were used for the analysis^18;59^. We first performed a sensitivity analysis of the posterior probability of colocalisation (PPH4) on the prior probability of colocalisation (p12), going from *p*12 = 10^−^8 to *p*12 = 10^−^4 with the default threshold being *p*12 = 10^−^5. eQTL signal and GWAS signals were deemed to colocalise if these two criteria were met: (1) At *P*12 = 5 × 10^−^5 the probability of colocalisation *PPH*4 > 0.5 and (2) At *p*12 = 10^−^5 the probability of independent signal (PPH3) was not the main hypothesis (*PPH*3 < 0.5). These criteria were chosen to allow eQTLs with weaker *P*-values due to lack of power in GTExv8, to be colocalised with the signal when the main hypothesis using small priors was that there was not any signal in the eQTL data.

As the chromosome 3 associated interval is larger than 200 kb, we performed additional colocalisation including a region up to 500 kb, but no further colocalisations were found.

