## Supplementary information for "Whole genome sequencing identifies multiple loci for critical illness caused by COVID-19"

### Supplementary material

**Meta-analysis by information content (MAIC)** **41**

**References** **42**

**GenOMICC Investigators** **42**

**Covid-19 Human Genetics Initiative** **63**

**23andMe Investigators** **63**

### Overview of analysis

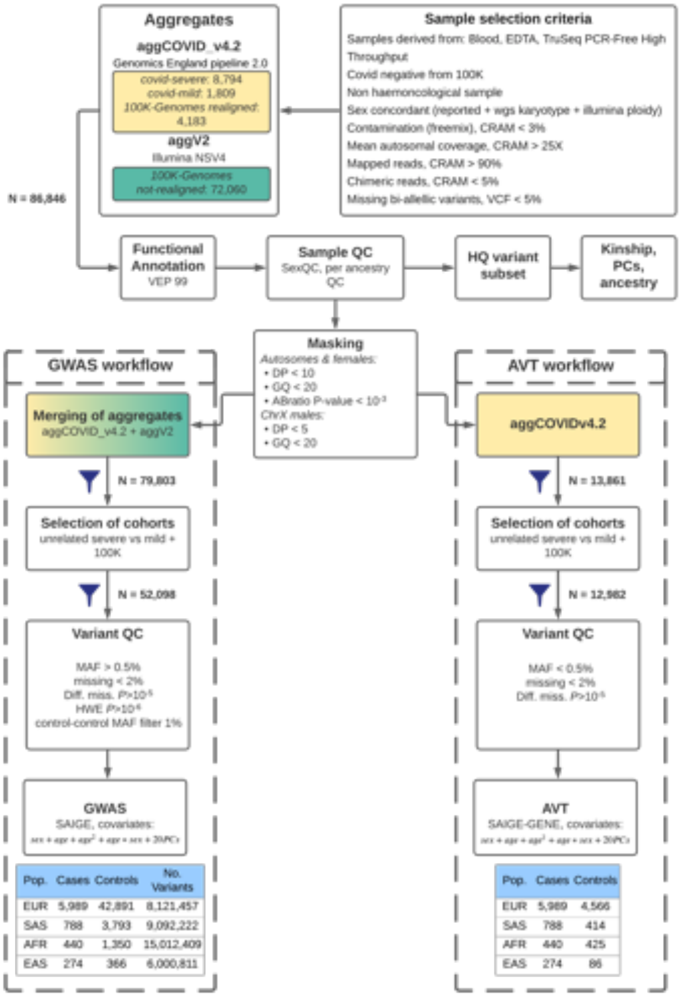

Figure 1: Diagram showing the analysis workflow for genome-wide association study (GWAS) and aggregate variant testing (AVT) analyses of this study.

### Cohort characteristics

#### Pre-QC breakdown

| Cohort | Instrument | Alignment & variant calling workflow | N (pre-QC) | Included in aggregate |
|---|---|---|---|---|
| *covid - severe* | NovaSeq | Genomics England pipeline 2.0 | 8,794 | aggCOVID_v4.2 |
| *covid - mild* | NovaSeq | Genomics England pipeline 2.0 | 1,809 | aggCOVID_v4.2 |
| *100K-Genomes(not realigned)* | Hiseq X | Illumina North Star Version 4 (NSV4, version 2.6.53.23) | 72,060 | aggV2 |
| *100K-Genomes(realigned)* | Hiseq X | Genomics England pipeline 2.0 | 4,183 | aggCOVID_v4.2 |

Table 1: Breakdown of the cohorts included in this study

#### SampleQC

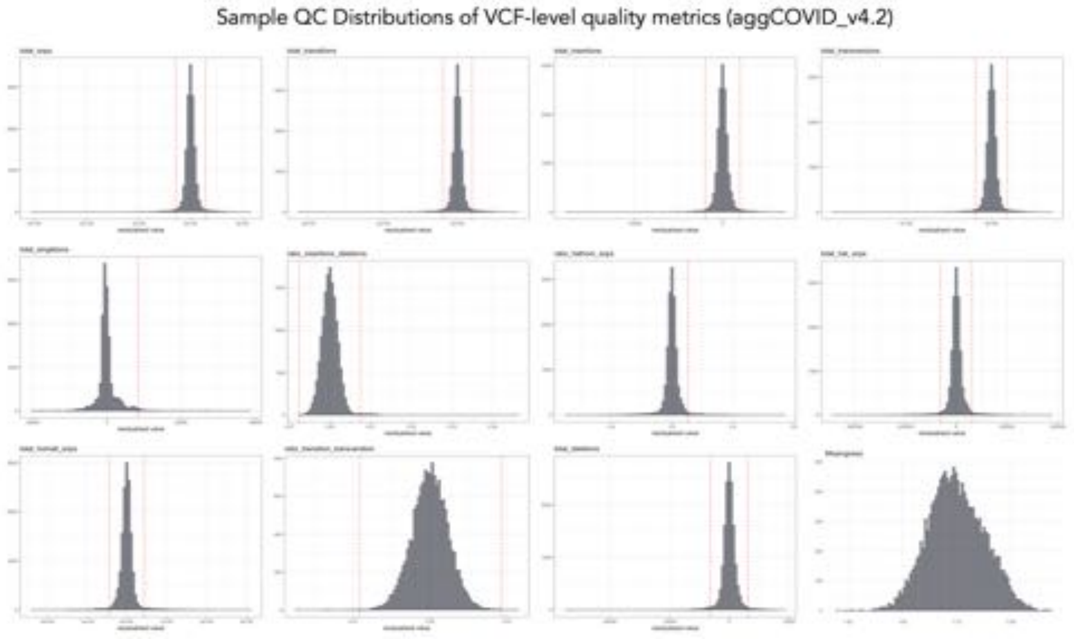

Figure 2: Each histogram shows the distribution of samples in the aggCOVID v4.2 data-set for a particular VCF-level quality metric, following adjustment for sequencing platform and the first three ancestry assignment principal components (as described in Methods). All metrics are calculated from autosomal bi-allelic SNVs. The dashed read lines indicate the threshold for sample exclusion. Samples were removed that were four median absolute deviations (MADs) above or below the median for the following metrics: ratio heterozygous-homozygous, ratio insertions-deletions, ratio transitions-transversions, total deletions, total insertions, total heterozygous snps, total homozygous snps, total transitions, total transversions. For the number of total singletons (snps), samples were removed that were more than 8 MADs above the median. For the ratio of heterozygous to homozygous alternate snps, samples were removed that were more than 4 MADs above the median. For sample-missingness (bottom-right panel), a hard cut-off of 0.05 was applied (no adjustment for sequencing platform or ancestry).

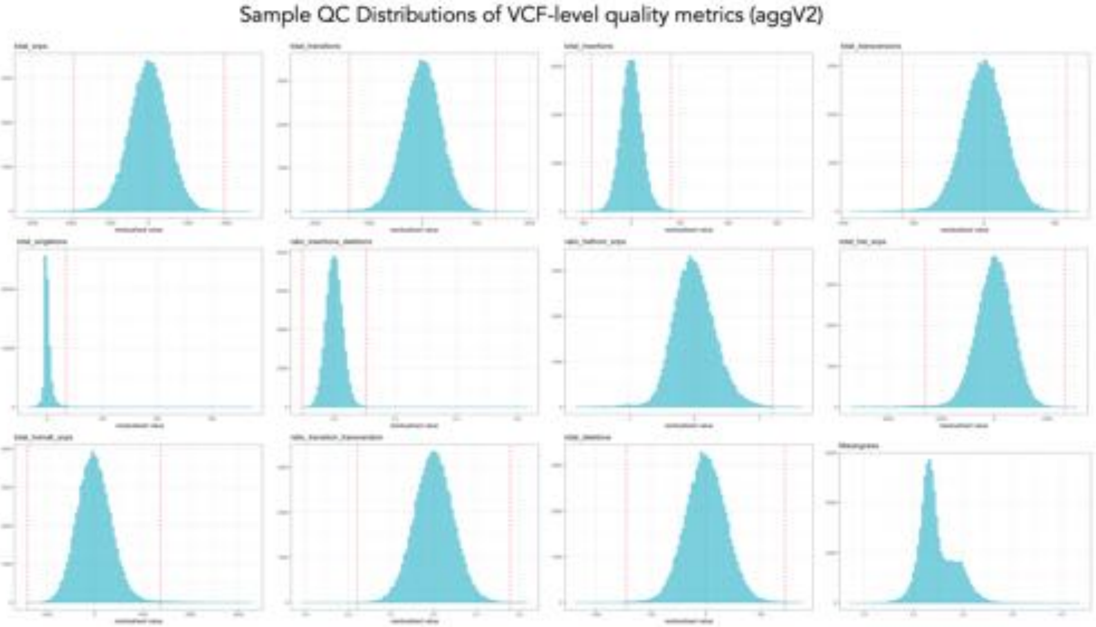

Figure 3: Each histogram shows the distribution of samples in the aggV2 data-set for a particular VCF-level quality metric, following adjustment for sequencing platform and the first three ancestry assignment principal components (as described in Methods). All metrics are calculated from autosomal bi-allelic SNVs. The dashed read lines indicate the threshold for sample exclusion. Samples were removed that were four median absolute deviations (MADs) above or below the median for the following metrics: ratio heterozygous-homozygous, ratio insertions-deletions, ratio transitions-transversions, total deletions, total insertions, total heterozygous snps, total homozygous snps, total transitions, total transversions. For the number of total singletons (snps), samples were removed that were more than 8 MADs above the median. For the ratio of heterozygous to homozygous alternate snps, samples were removed that were more than 4 MADs above the median. For sample-missingness (bottom-right panel), a hard cut-off of 0.05 was applied (no adjustment for sequencing platform or ancestry).

#### PCA and ancestry

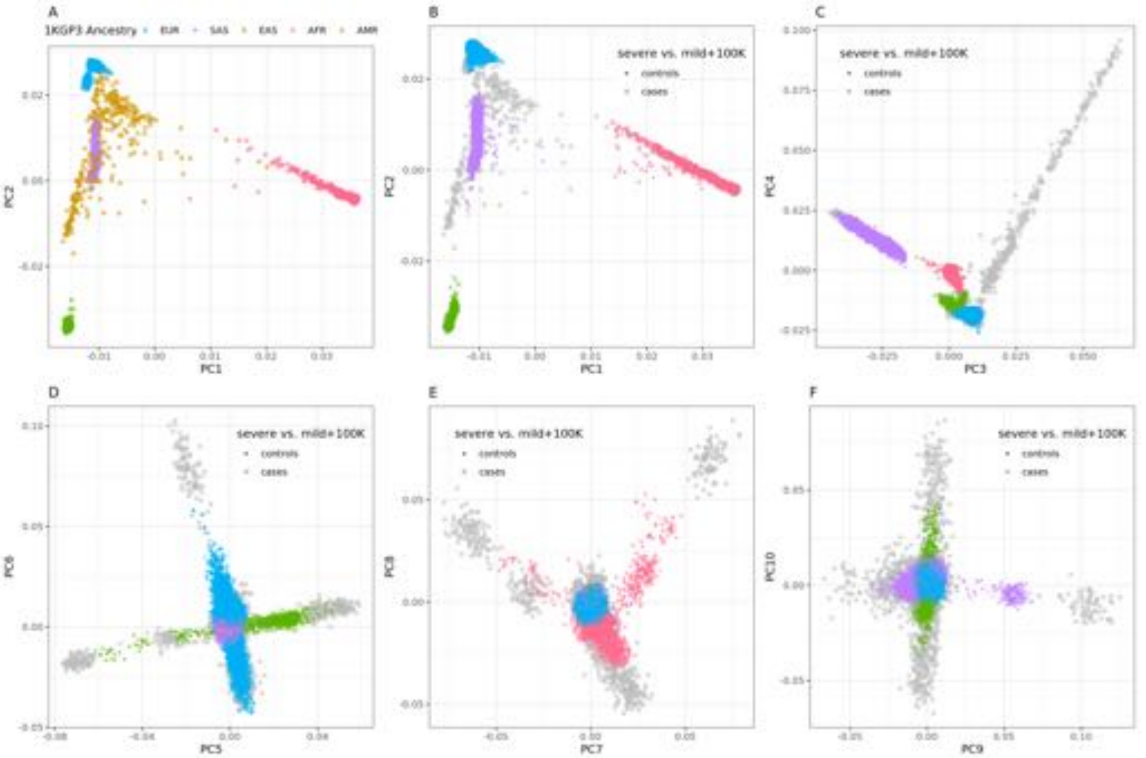

Figure 4: Projection of severe, mild and 100K individuals onto the PCs of the 1KGP3. (A) PCs 1 & 2 for 1KGP3 unrelated individuals using the high quality independent SNP set. (B-F). Projected PCs 1-10 for severe (cases) and mild + 100K individuals (controls). 1KGP3 reference individuals are shown in grey.

#### Post-QC breakdown

#### Age and sex breakdown

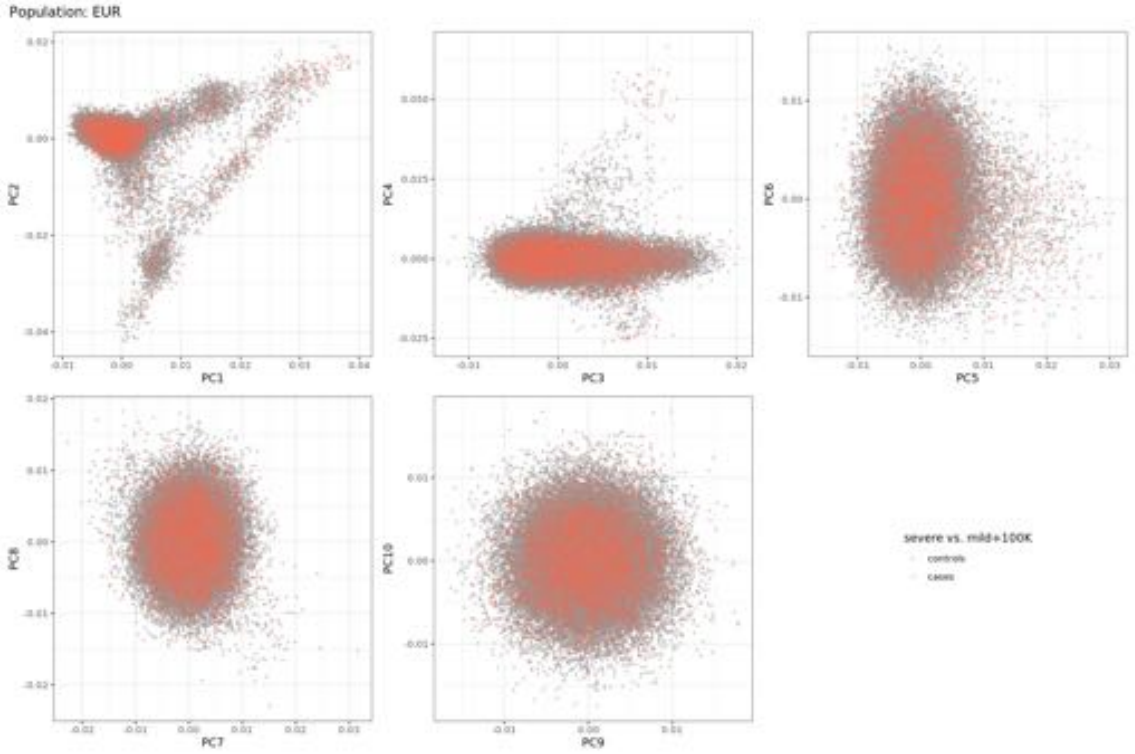

Figure 5: EUR PCs 1-10 for severe (cases) and mild + 100K individuals (controls).

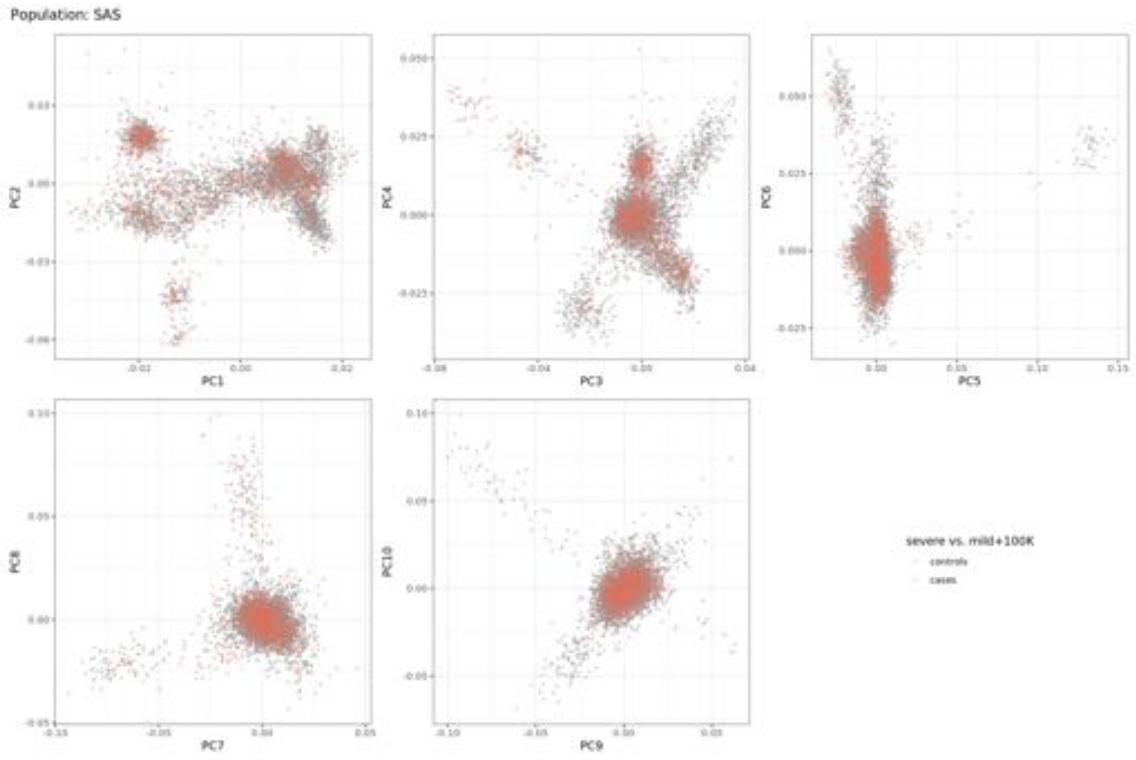

Figure 6: SAS PCs 1-10 for severe (cases) and mild + 100K individuals (controls).

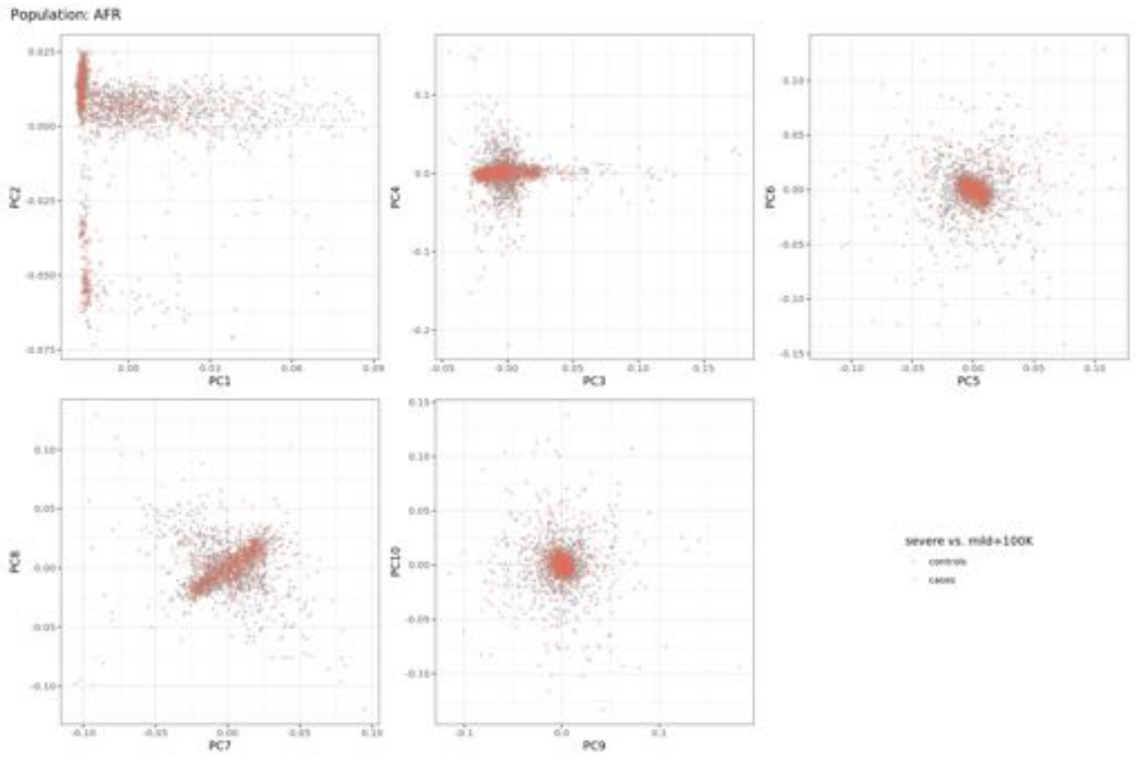

Figure 7: AFR PCs 1-10 for severe (cases) and mild + 100K individuals (controls).

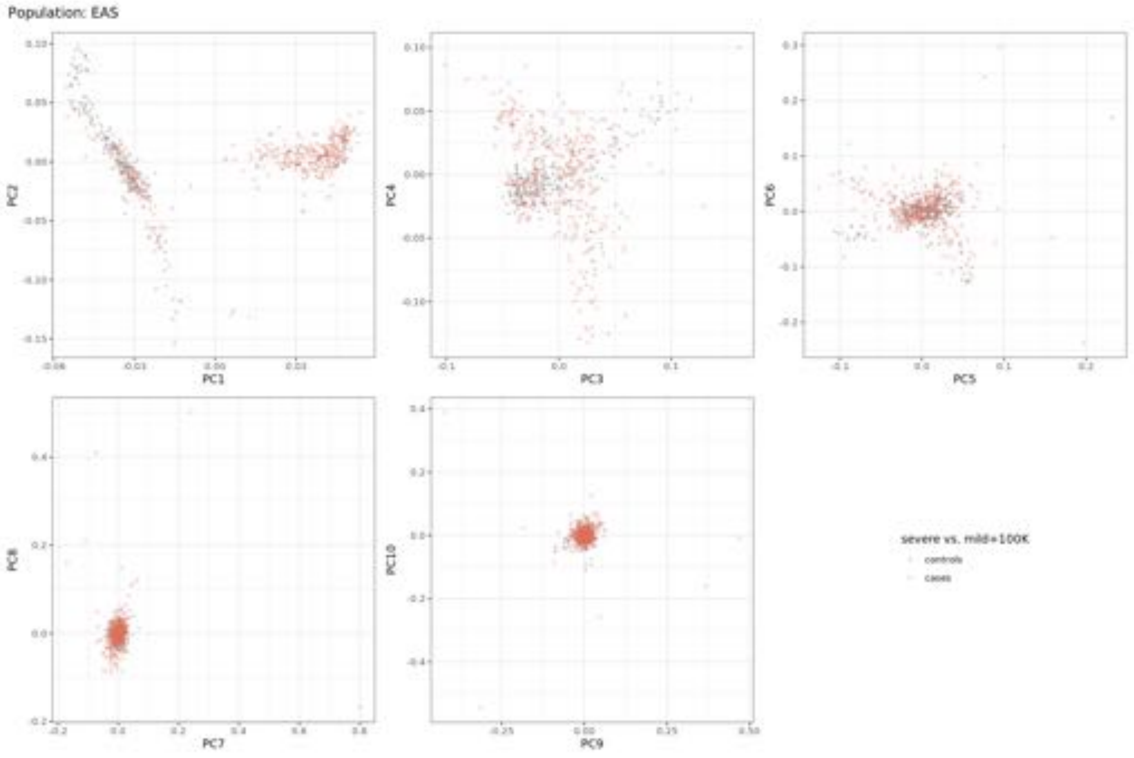

Figure 8: EAS PCs 1-10 for severe (cases) and mild + 100K individuals (controls).

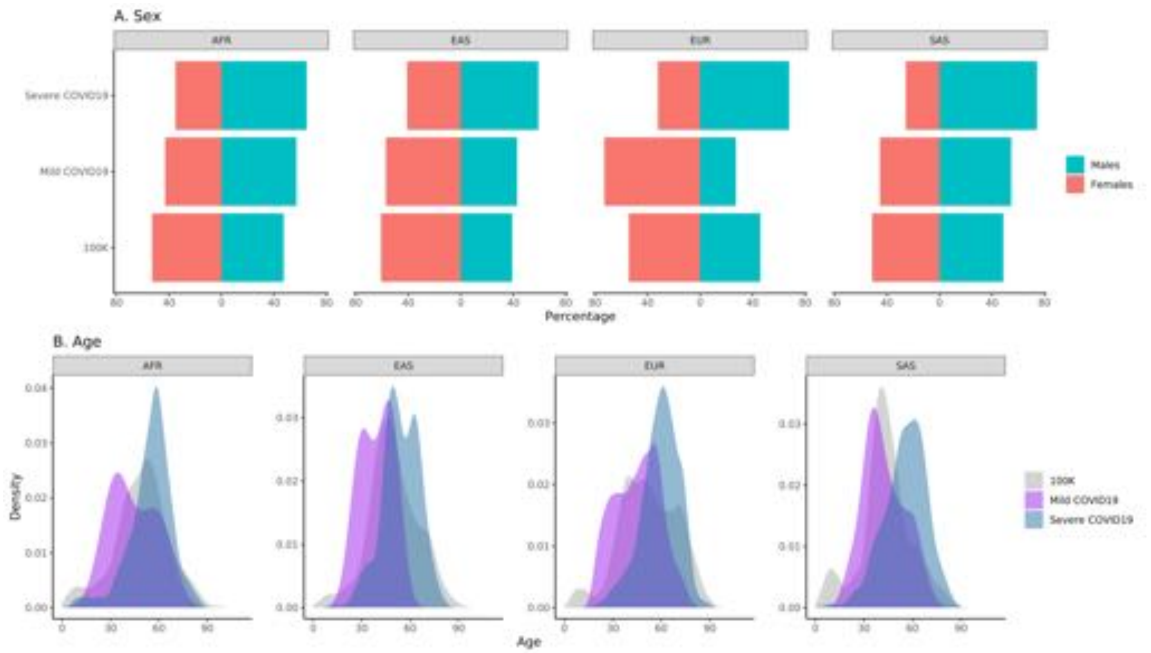

Figure 9: Breakdown of sex and age for severe, mild and 100K cohorts.

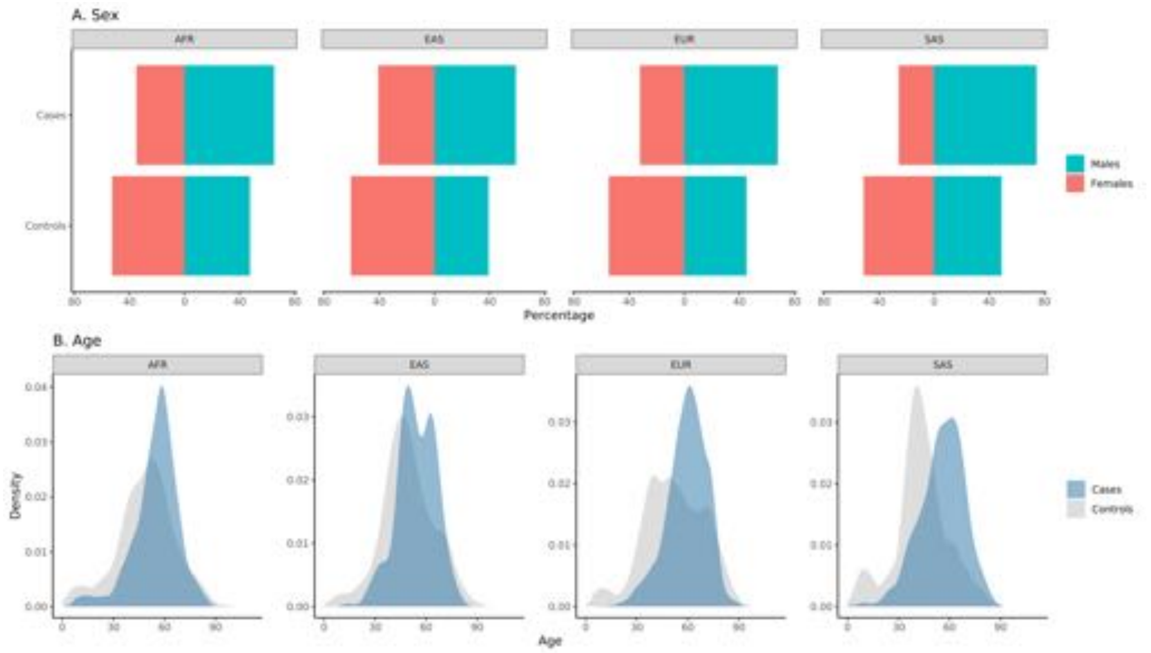

Figure 10: Breakdown of sex and age for cases and controls of the severe (cases) vs. mild+100K (controls) cohorts.

| Predicted Ancestry | *covid-severe* | *covid-mild* | *100K-Genomes (not realigned)* | *100K-Genomes (realigned)* |
|---|---|---|---|---|
| EUR | 5,989 | 1,507 | 38,325 | 3,059 |
| SAS | 788 | 95 | 3379 | 319 |
| AFR | 440 | 14 | 1025 | 311 |
| EAS | 274 | 14 | 280 | 72 |

Table 2: Breakdown of the cohorts included in this study by predicted ancestry, after sample QC and removal of related individuals.

### GWAS

#### Per-population GWAS results

| Population | No. of ld-pruned variants | Bonferroni-corrected P-value threshold |
|---|---|---|
| EUR | 2264479 | 2.2e-08 |
| SAS | 2729540 | 1.8e-08 |
| AFR | 5370001 | 9.3e-09 |
| EAS | 1264431 | 4e-08 |

Table 3: The Bonferoni-corrected $P$-values for the per-population GWAS analyses. The $P$-value significance threshold was calculated by estimating the effective number of tests. After selecting the final filtered set of tested variants for each population, we LD-pruned in a window of 250Kb and $r^2 = 0.8$ with plink 1.9.

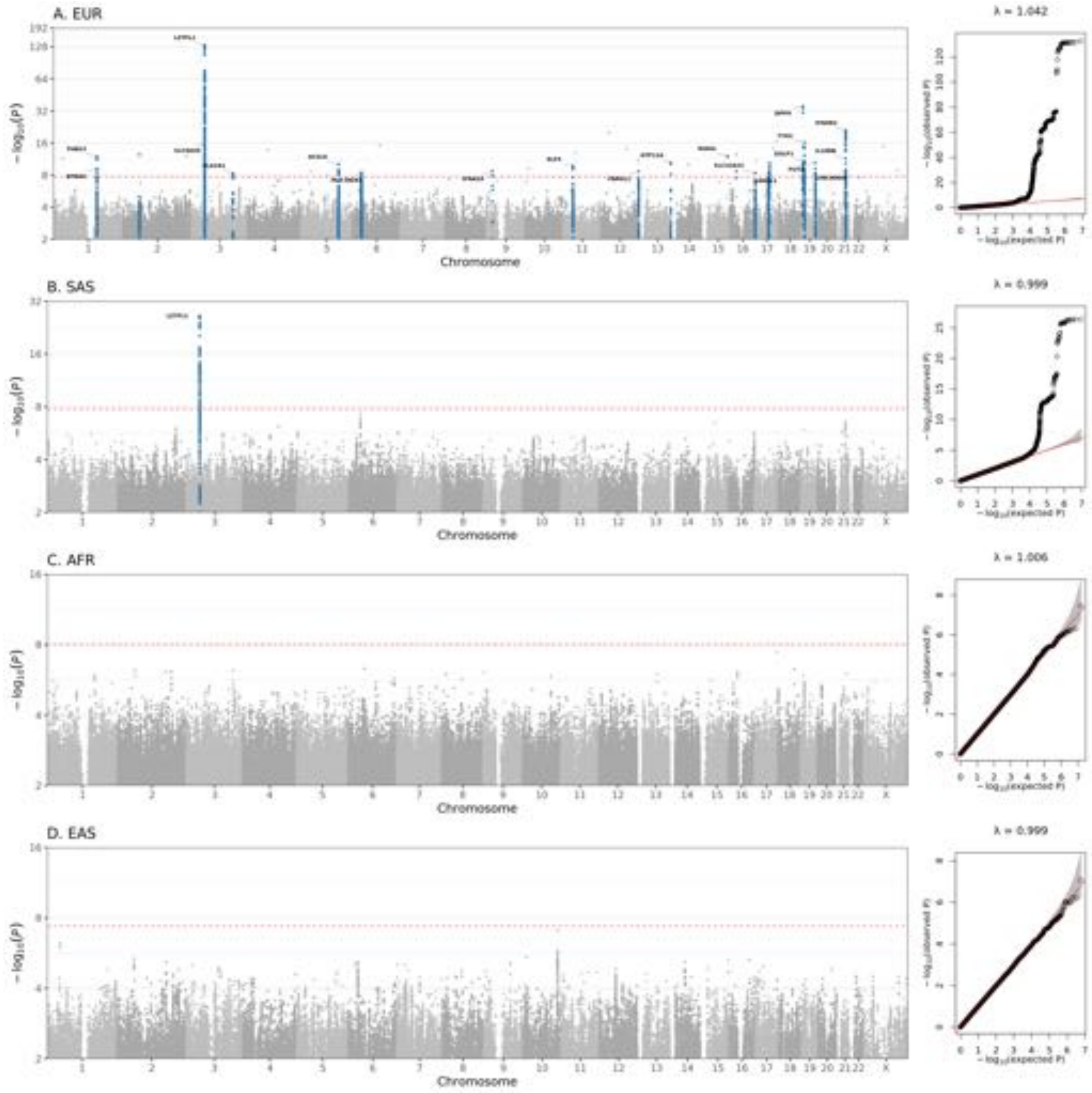

Figure 11: Manhattan plots showing GWAS results for each population cohort. The highlighted results with blue are the variants that are LD clumped ($r^2$=0.1, $P_2$=0.01 in each population) with each lead variant. Red dashed line is the Bonferroni-corrected $P$-value according to the number of estimated independent tests in each population.

#### LD-based validation of lead GWAS signals

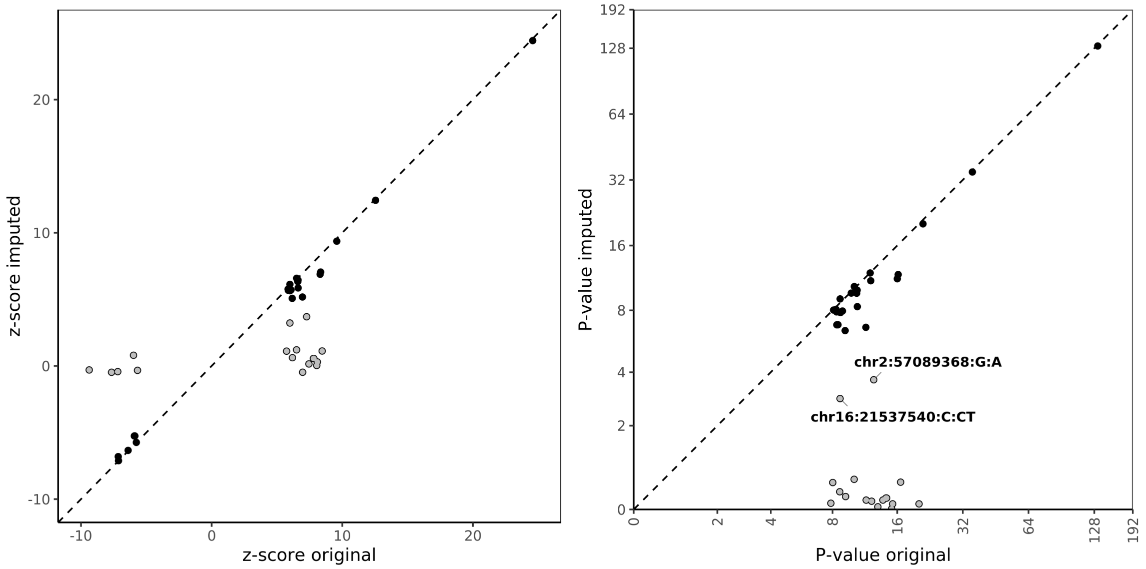

Figure 12: Original and imputed zscores and respective $P$-values with leave-one procedure for lead variants of the EUR analysis. Variants with low support from neighbouring variants are highlighted with grey.

### Association signal forest plots

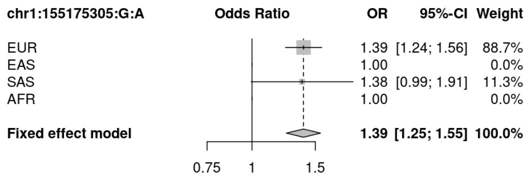

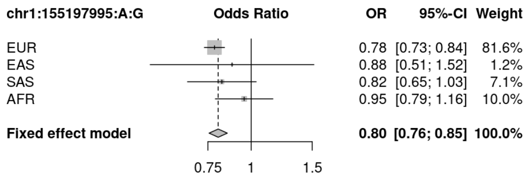

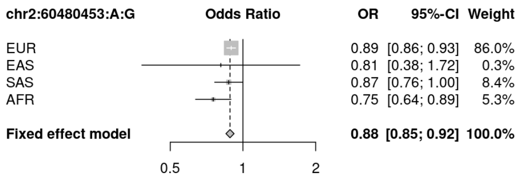

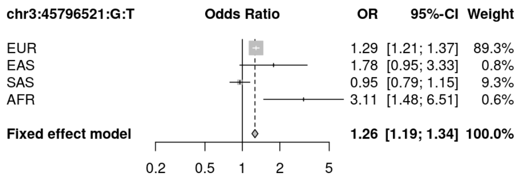

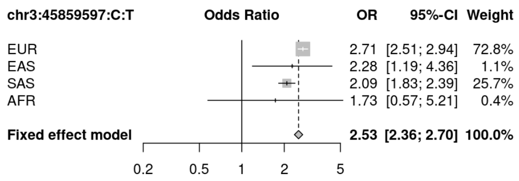

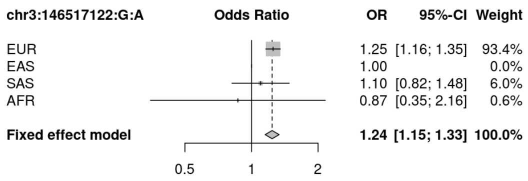

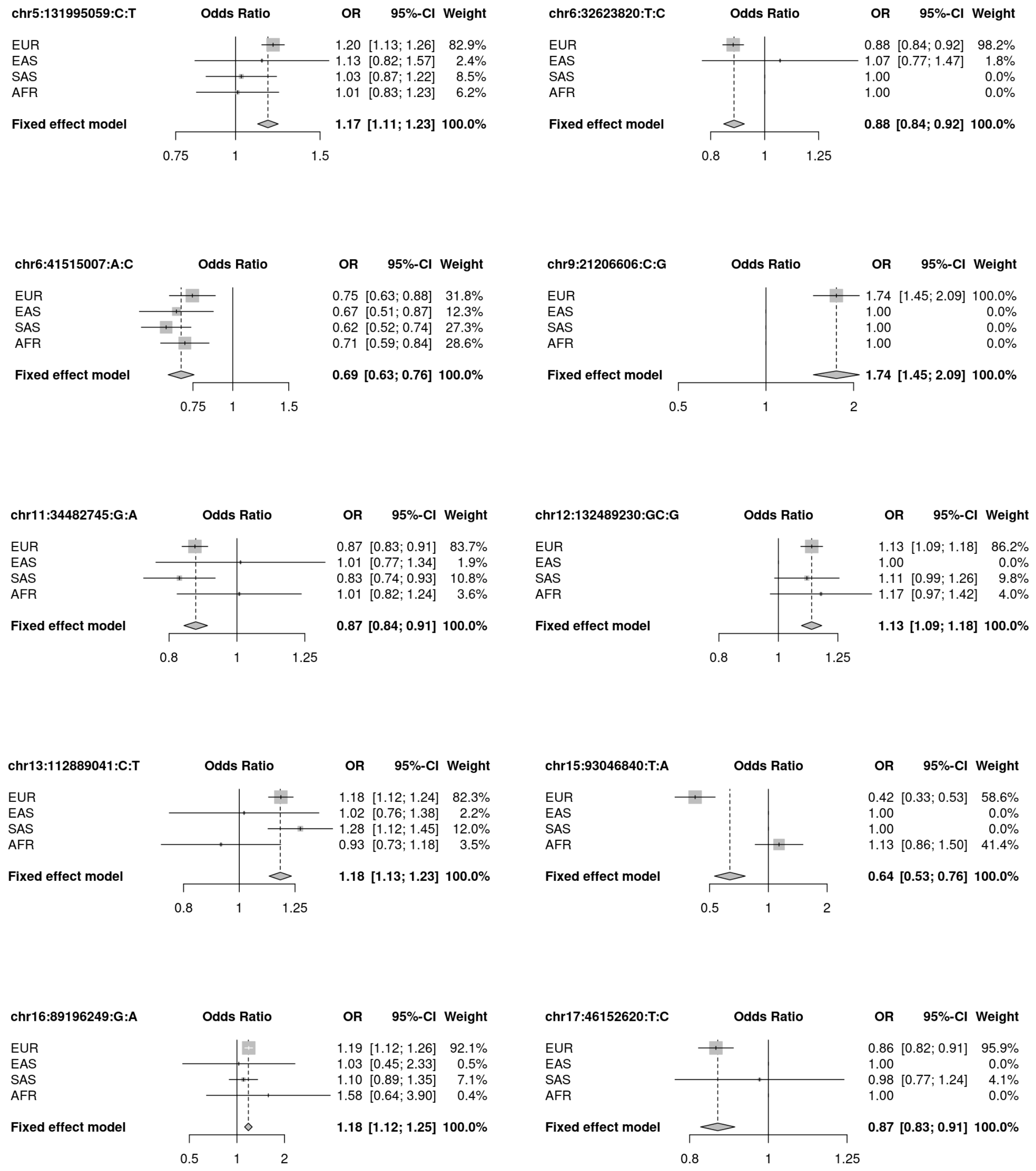
chr5:131995059:C:T Odds Ratio OR 95%-CI Weight
EUR 1.20 [1.13; 1.26] 82.9%
EAS 1.13 [0.82; 1.57] 2.4%
SAS 1.03 [0.87; 1.22] 8.5%
AFR 1.01 [0.83; 1.23] 6.2%
Fixed effect model 1.17 [1.11; 1.23] 100.0%
0.75 1 1.5
chr6:32623820:T:C Odds Ratio OR 95%-CI Weight
EUR 0.88 [0.84; 0.92] 98.2%
EAS 1.07 [0.77; 1.47] 1.8%
SAS 1.00 0.0%
AFR 1.00 0.0%
Fixed effect model 0.88 [0.84; 0.92] 100.0%
0.8 1 1.25
chr6:41515007:A:C Odds Ratio OR 95%-CI Weight
EUR 0.75 [0.63; 0.88] 31.8%
EAS 0.67 [0.51; 0.87] 12.3%
SAS 0.62 [0.52; 0.74] 27.3%
AFR 0.71 [0.59; 0.84] 28.6%
Fixed effect model 0.69 [0.63; 0.76] 100.0%
0.75 1 1.5
chr9:21206606:C:G Odds Ratio OR 95%-CI Weight
EUR 1.74 [1.45; 2.09] 100.0%
EAS 1.00 0.0%
SAS 1.00 0.0%
AFR 1.00 0.0%
Fixed effect model 1.74 [1.45; 2.09] 100.0%
0.5 1 2
chr11:34482745:G:A Odds Ratio OR 95%-CI Weight
EUR 0.87 [0.83; 0.91] 83.7%
EAS 1.01 [0.77; 1.34] 1.9%
SAS 0.83 [0.74; 0.93] 10.8%
AFR 1.01 [0.82; 1.24] 3.6%
Fixed effect model 0.87 [0.84; 0.91] 100.0%
0.8 1 1.25
chr12:132489230:GC:G Odds Ratio OR 95%-CI Weight
EUR 1.13 [1.09; 1.18] 86.2%
EAS 1.00 0.0%
SAS 1.11 [0.99; 1.26] 9.8%
AFR 1.17 [0.97; 1.42] 4.0%
Fixed effect model 1.13 [1.09; 1.18] 100.0%
0.8 1 1.25
chr13:112889041:C:T Odds Ratio OR 95%-CI Weight
EUR 1.18 [1.12; 1.24] 82.3%
EAS 1.02 [0.76; 1.38] 2.2%
SAS 1.28 [1.12; 1.45] 12.0%
AFR 0.93 [0.73; 1.18] 3.5%
Fixed effect model 1.18 [1.13; 1.23] 100.0%
0.8 1 1.25
chr15:93046840:T:A Odds Ratio OR 95%-CI Weight
EUR 0.42 [0.33; 0.53] 58.6%
EAS 1.00 0.0%
SAS 1.00 0.0%
AFR 1.13 [0.86; 1.50] 41.4%
Fixed effect model 0.64 [0.53; 0.76] 100.0%
0.5 1 2
chr16:89196249:G:A Odds Ratio OR 95%-CI Weight
EUR 1.19 [1.12; 1.26] 92.1%
EAS 1.03 [0.45; 2.33] 0.5%
SAS 1.10 [0.89; 1.35] 7.1%
AFR 1.58 [0.64; 3.90] 0.4%
Fixed effect model 1.18 [1.12; 1.25] 100.0%
0.5 1 2
chr17:46152620:T:C Odds Ratio OR 95%-CI Weight
EUR 0.86 [0.82; 0.91] 95.9%
EAS 1.00 0.0%
SAS 0.98 [0.77; 1.24] 4.1%
AFR 1.00 0.0%
Fixed effect model 0.87 [0.83; 0.91] 100.0%
0.8 1 1.25

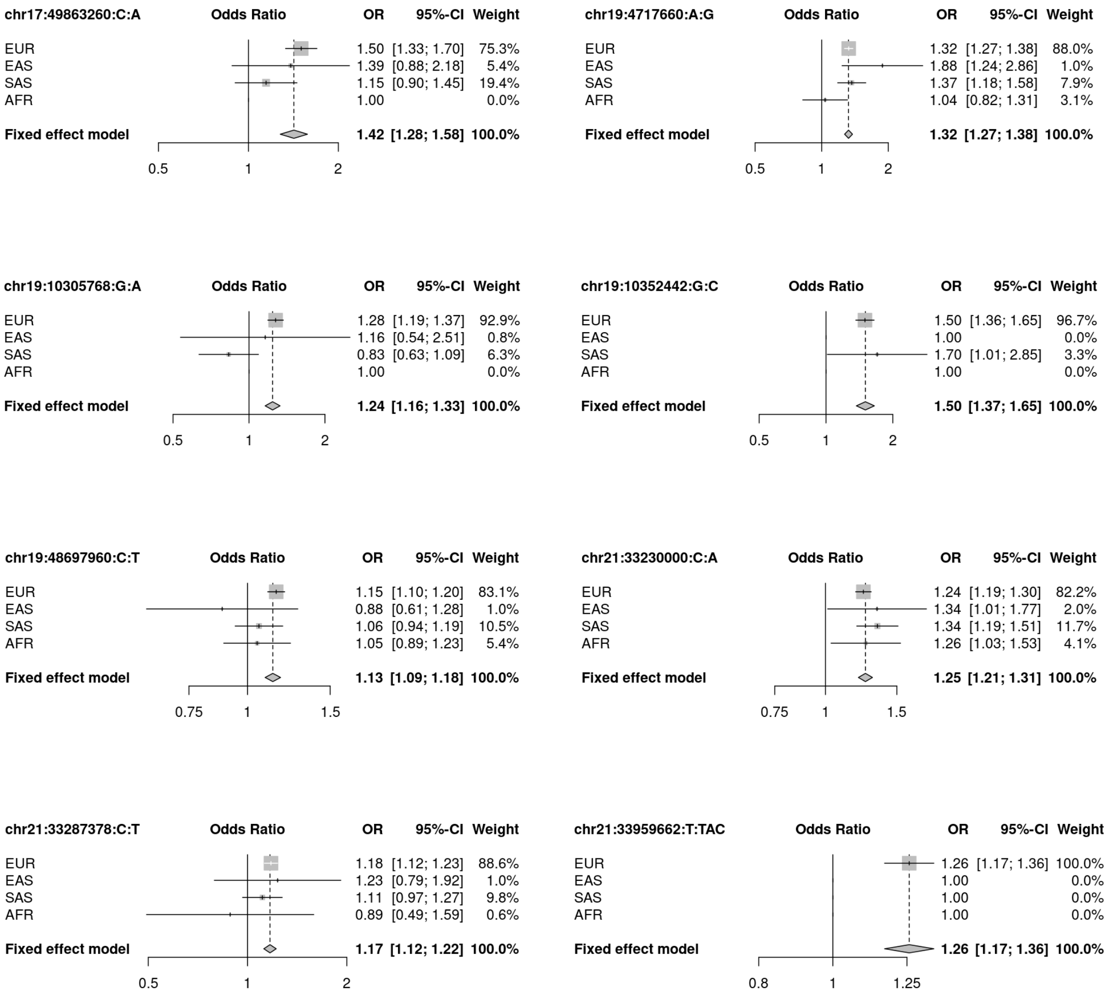

Figure 13: Forest plots for all lead signals.

### Replication forestplots

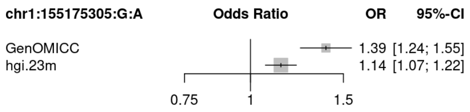

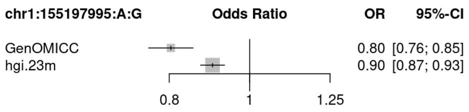

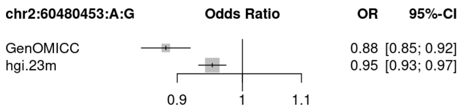

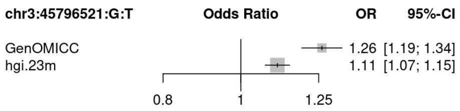

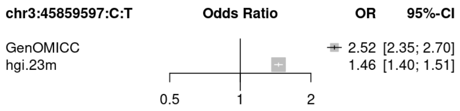

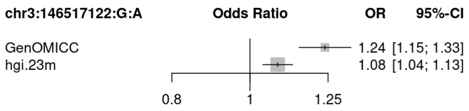

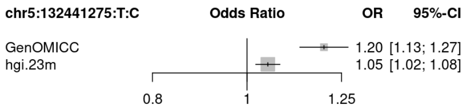

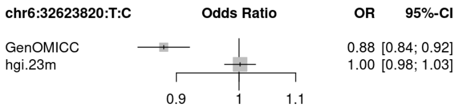

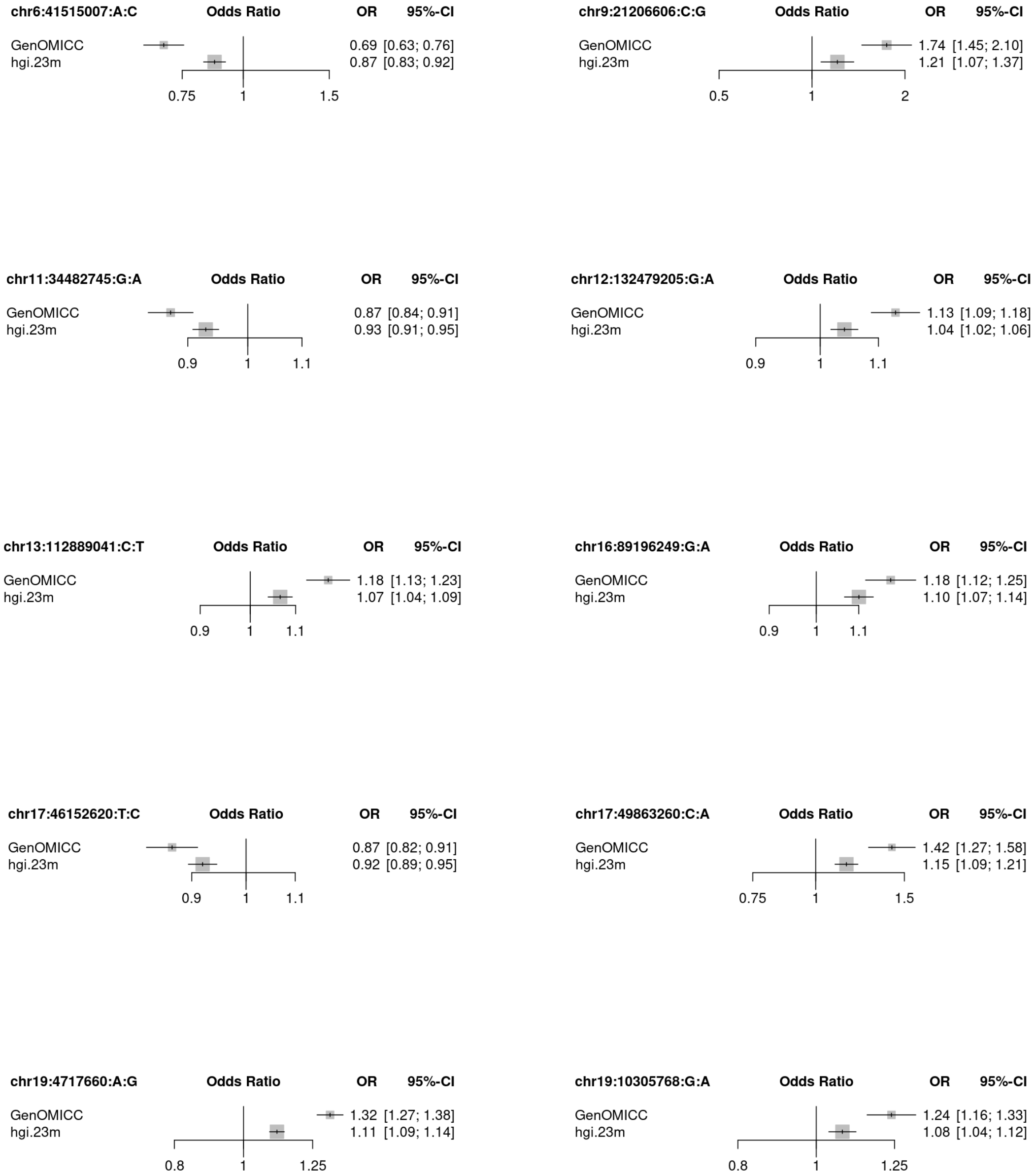
chr6:41515007:A:C
Odds Ratio
OR
95%-CI
GenOMICC
0.69 [0.63; 0.76]
hgi.23m
0.87 [0.83; 0.92]
0.75
1
1.5
chr9:21206606:C:G
Odds Ratio
OR
95%-CI
GenOMICC
1.74 [1.45; 2.10]
hgi.23m
1.21 [1.07; 1.37]
0.5
1
2
chr11:34482745:G:A
Odds Ratio
OR
95%-CI
GenOMICC
0.87 [0.84; 0.91]
hgi.23m
0.93 [0.91; 0.95]
0.9
1
1.1
chr12:132479205:G:A
Odds Ratio
OR
95%-CI
GenOMICC
1.13 [1.09; 1.18]
hgi.23m
1.04 [1.02; 1.06]
0.9
1
1.1
chr13:112889041:C:T
Odds Ratio
OR
95%-CI
GenOMICC
1.18 [1.13; 1.23]
hgi.23m
1.07 [1.04; 1.09]
0.9
1
1.1
chr16:89196249:G:A
Odds Ratio
OR
95%-CI
GenOMICC
1.18 [1.12; 1.25]
hgi.23m
1.10 [1.07; 1.14]
0.9
1
1.1
chr17:46152620:T:C
Odds Ratio
OR
95%-CI
GenOMICC
0.87 [0.82; 0.91]
hgi.23m
0.92 [0.89; 0.95]
0.9
1
1.1
chr17:49863260:C:A
Odds Ratio
OR
95%-CI
GenOMICC
1.42 [1.27; 1.58]
hgi.23m
1.15 [1.09; 1.21]
0.75
1
1.5
chr19:4717660:A:G
Odds Ratio
OR
95%-CI
GenOMICC
1.32 [1.27; 1.38]
hgi.23m
1.11 [1.09; 1.14]
0.8
1
1.25
chr19:10305768:G:A
Odds Ratio
OR
95%-CI
GenOMICC
1.24 [1.16; 1.33]
hgi.23m
1.08 [1.04; 1.12]
0.8
1
1.25

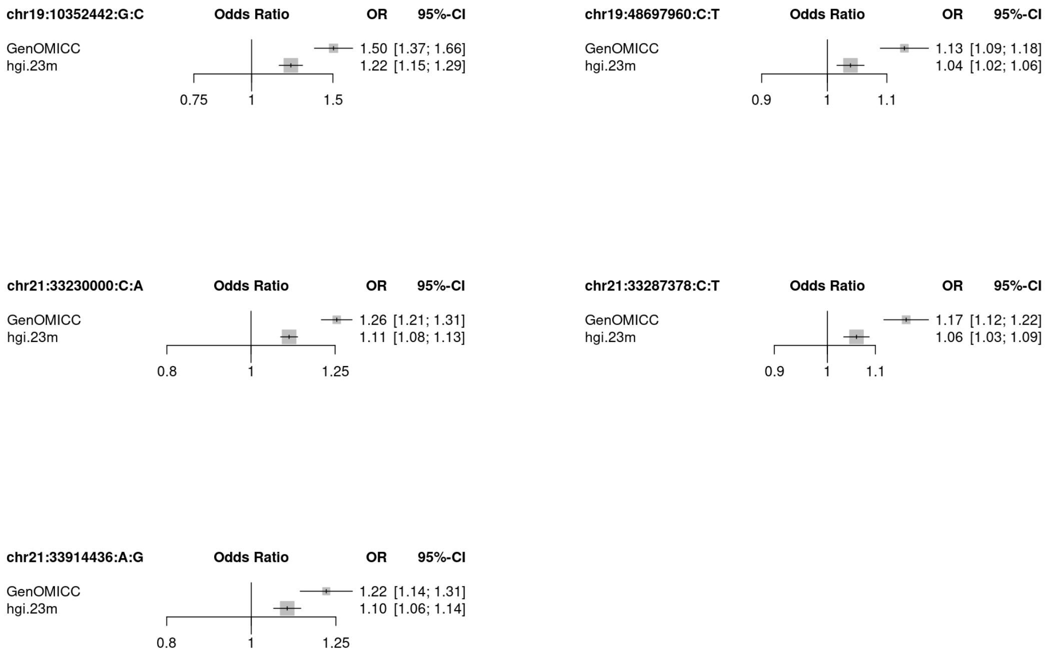

Figure 14: Forest plots comparing the Odds ratios for all the lead signals with a combined meta-analysis of HGI freeze 6 B2 and 23andMe. The HGI summaries were produced with new meta-analysis that removed the GENOMICC cases to ensure statistical independence. The Genomics England 100K participants (i.e, controls) summary data contributed to HGI C2 analysis and not B2 used here.

#### Genetic fine-mapping and colocalisation

In the EUR ancestry group, we found two independent signals for the association at 1q22. The lead variant for the first fine-mapped locus is a synonymous variant in *EFNA4* (chr1:155066988:C:T), while the lead variant of the second independent signal (chr1:155197995:A:G) is an intronic variant in *THSB3*, in close proximity to the most significant single-tissue eQTLs for *MUC1* (chr1:155199564:G:T, chr1:155199139:G:A) in GTEx v8 , which are also in the same intronic region. Fine mapping the trans-ancestry meta-analysis revealed a third independent signal at this locus, with the lead variant (chr1:155175305:G:A, rs7528026, OR:1.39, 95% CIs:[1.24-1.55]) being in an intron of *TRIM46*. This variant is an sQTL and eQTL for *MUC1* in lung and and whole blood tissue, respectively, in GTEx v8[1] (Supplementary File TWAS.xlsx).

Meta-analysis across genetically inferred ancestries revealed a novel locus at 2p16.1, with the lead variant (chr2:60480453:A:G, OR:0.88, 95 %CIs:[0.85,0.92]) being in an intron of *BCL11A*.

We fine-mapped the signal in the 3p21.31 region, first reported by Ellighaus *et al*,[2] into two independent associations. The lead variant for the first association is in the 5' UTR region of *SLC6A20* (chr3:45796521:G:T, OR:1.29, 95%CIs:[1.21,1.37]). The second association in the chr3p21.31 region is seen at genome-wide significance in both the EUR and SAS cohorts, with two of the three highest ranked variants in the fine-mapped region in the two populations shared between the two cohorts (chr3:45818159:G:A, chr3:45859597:C:T) and residing in downstream and intronic regions of *LZTFL1*.

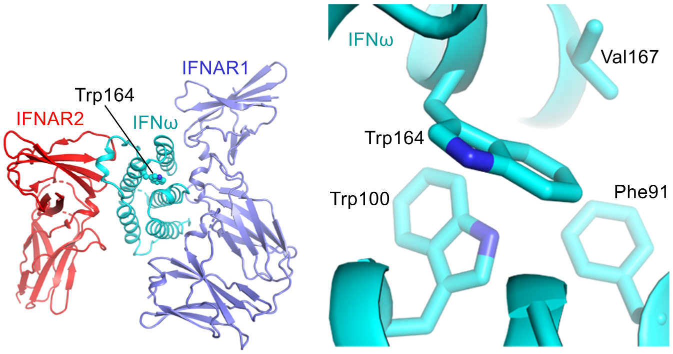

Figure 15: Structural predictions for impact of risk variant on IFNA10

The credible set for the 3q24 association included 9 variant and the lead variant (chr3:146517122:G:A, rs343320,p.His262Tyr, OR:1.24, 95%CIs [1.15-1.33]) is a missense variant in PLSCR1, predicted to be damaging by CADD (CADD:22.6).

At 5q31.1, the lead variant (chr5:131995059:C:T, rs56162149, OR:1.17, 95%CIs:[1.11,1.23]) is in an intron of *ACSL6*. The credible set for this locus contains 33 variants that span 484 kb including variants in genes *CSF2* and *IRF1-AS1*, with chr5:132075767:T:C being a missense variant in *CSF2* and chr5:131991772:C:G being missense in *ACSL6* and only intronic variants for *IRF1-AS1*.

The previously reported signal at 6p21.1, linked to *FOXP4*[3], is stronger in the SAS cohort but has a consistent effect across ancestries ($P_{het}$=0.49).

We fine mapped the signal at 9p21.3 to three variants with lead variant (chr9:21206606:C:G, rs28368148,p.Trp164Cys, OR:1.74, 95% CIs [1.45-2.09]) being a missense variant in IFNA10 that is predicted to be damaging by CADD (CADD:23.9) with potential functional impact Supplementary Figure 15.

The signal in the 11p13 region was fine-mapped to four variants (lead variant chr11:34482745:G:A, rs61882275, OR:0.87, 95%CIs:[0.84-0.91]), all four of which are in an intron of *ELF5*.

The credible set for the signal in the 12q24.33 region includes 24 variants spanning 95 kb, of which the lead (chr12:132489230:GC:G, rs56106917, OR:1.13, 95% CIs:[1.09-1.18]) lies upsdtream *FBRSL1*.

The signal at 13q34 was fine-mapped to four variants, with lead variant (13:112889041:C:T, rs9577175, OR:1.18, 95%CIs [1.12-1.24]) lying downstream of *ATP11A* and upstream of *MCF2L* genes.

The association at 15q26.1 was fine mapped to two variants with lead variant (chr15:93046840:T:A, rs4424872, OR: 2.37, 95% CIs:[ 1.87-3.01] in an intron of RGMA. This a low frequency (allele frequency <1%) variant that was not replicated due to lack of coverage in the available replication data and further validation in an independent dataset is recommended.

The credible set for the association with lead variant at chr17:46152620:T:C (rs2532300, OR:1.16, 95% CIs:[1.10,1.22]) includes 1430 variants and spans 658 kb, indicating an association with the known inversion haplotype at 17q21.31[4].

We fine mapped the signal at 17q21.33 to five variants, with lead variant (chr17:49863260:C:A, rs3848456, OR:1.5, 95% CIs:[1.33-1.70] residing in a regulatory element (ENSR00001010694) 15.5 kb upstream of the TAC4 gene.

In the 19p13.3 region, which we reported in 2020,[5], we fine-mapped the signal to a single variant in *DPP9* (chr19:4717660:A:G). This variant is a missense variant in transcript ENST00000599248 but intronic in other transcripts including the MANE transcript (ENST00000262960.14).

In the 19p13.2 region, where we previously reported a variant associated with *TYK2*,[5] we find two independent signals, one of which is a damaging missense variant in *TYK2*, chr19:10352442:G:C (rs34536443, OR:1.50, 95% CIs:[1.36,1.65], CADD=25.1), and the second is an intronic variant of *ZGLP1* (19:10305768:G:A, rs73510898, OR:1.28, 95% CIs:[1.19,1.37]).

The signal at 19q13.33 was fine-mapped to ten variants, with the lead variant (chr19:48697960:C:T, rs368565, OR:1.15, 95%CIs [1.1-1.2]) in an intron of *FUT2* and variant chr19:48705753:T:C (rs503279) being in the 3' UTR of the MANE transcript for this gene.

In the 21q22.11 region, we described previously,[5] fine-mapping revealed three independent signals, for which the lead variants reside in the 5' UTR of *IFNAR2* (chr21:33230000:C:G,rs17860115, OR:1.24, 95% CIs:[1.19-1.30], CADD=10.1), an intronic region of *IL10RB* (chr21:33287378:C:T, rs8178521, OR:1.18, 95% CIs:[1.12,1.23]) and in a downstream long non-coding RNA (chr21:33959662:T:TAC, rs35370143, OR:1.26, 95% CIs:[1.17,1.36]).

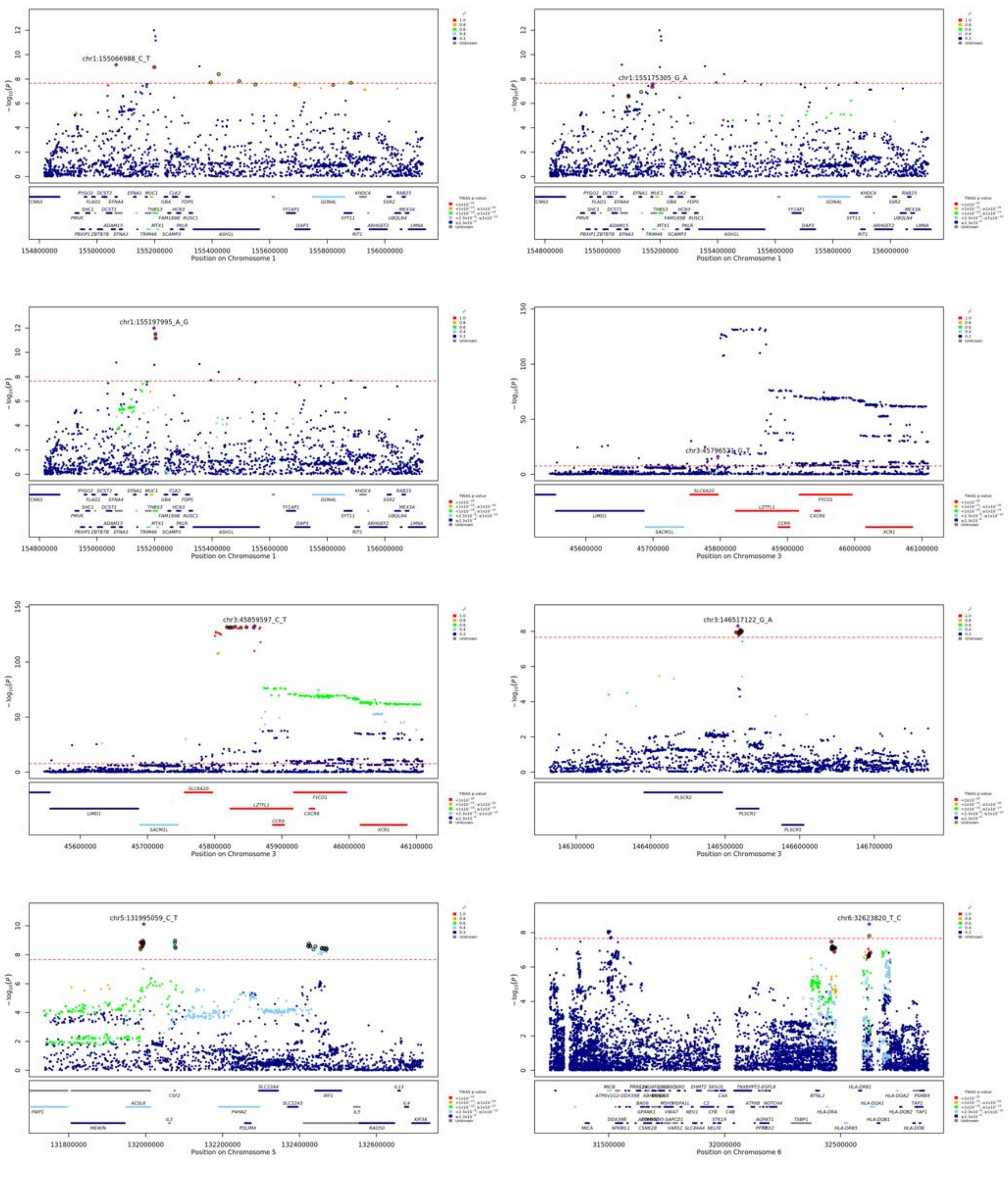
chr1:155066988_C_T
chr1:155175305_G_A
chr1:155197995_A_G
chr3:45796521_G_T
chr3:45859597_C_T
chr3:146517122_G_A
chr5:131995059_C_T
chr6:32623820_T_C
Position on Chromosome 1
Position on Chromosome 3
Position on Chromosome 5
Position on Chromosome 6

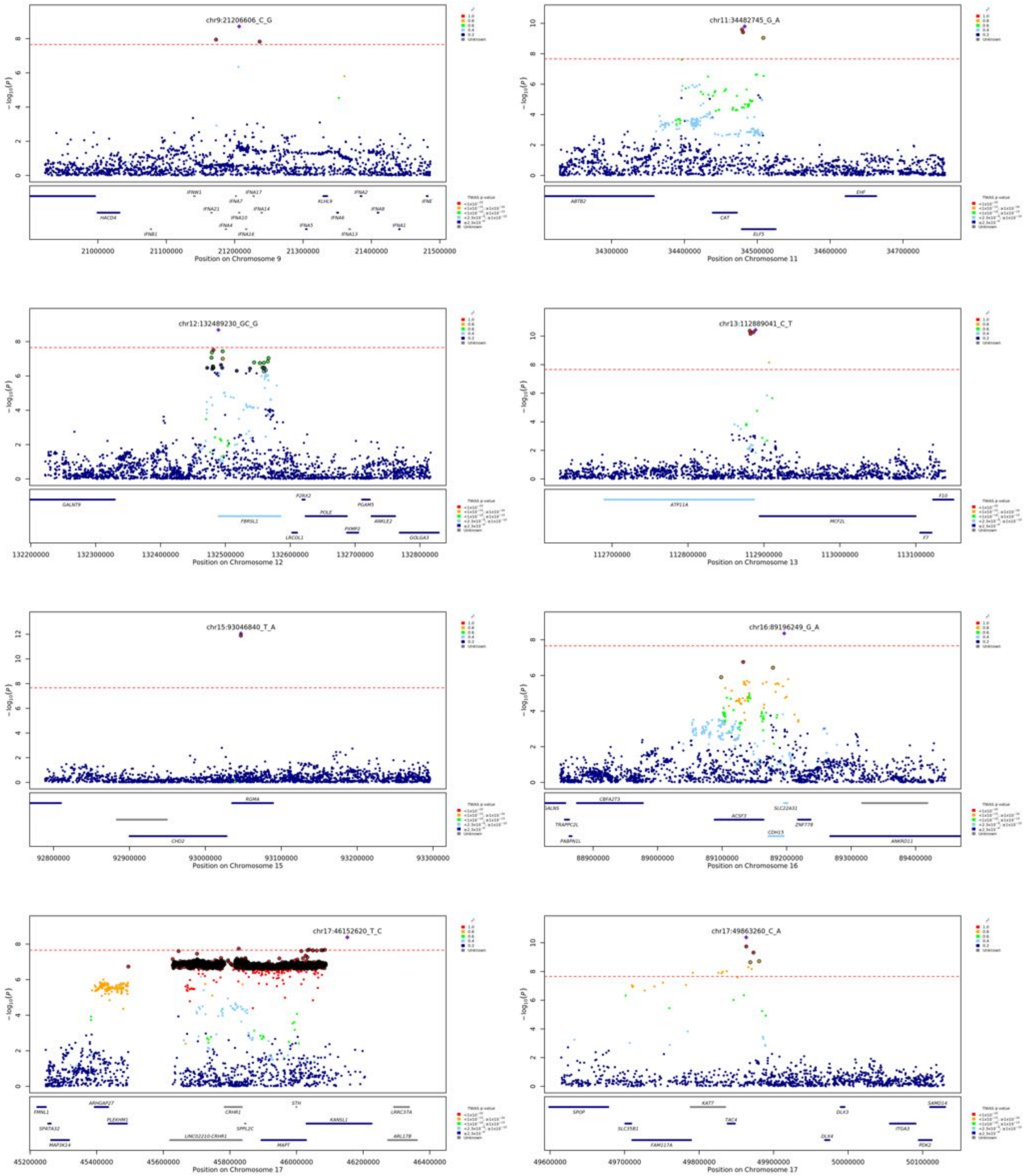
chr9:21206606_C_G
Position on Chromosome 9
chr11:34482745_G_A
Position on Chromosome 11
chr12:132489230_GC_G
Position on Chromosome 12
chr13:112889041_C_T
Position on Chromosome 13
chr15:93046840_T_A
Position on Chromosome 15
chr16:89196249_G_A
Position on Chromosome 16
chr17:46152620_T_C
Position on Chromosome 17
chr17:49863260_C_A
Position on Chromosome 17

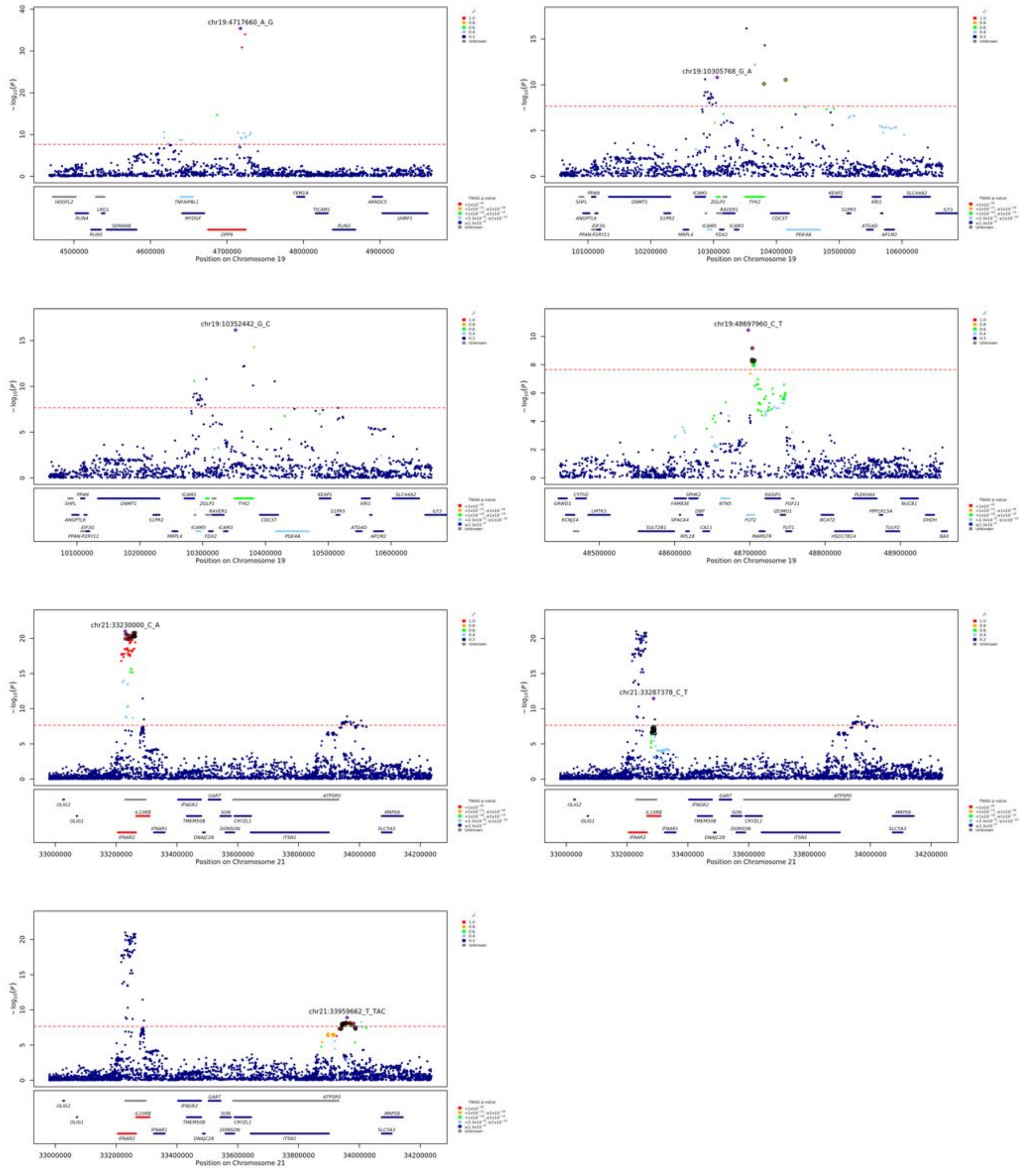

Figure 16: Locuszoom figures for the signals found in the per-population analyses. Upper panels show lead signals and LD calculated in EUR with all other loci in the window shown. Credible sets for each displayed signal that were inferred with susieR are displayed with outline black circles. On the bottom panels an hg38 gene track is displayed with colors matching significance from the metaTWAS analysis in discrete bins shown.

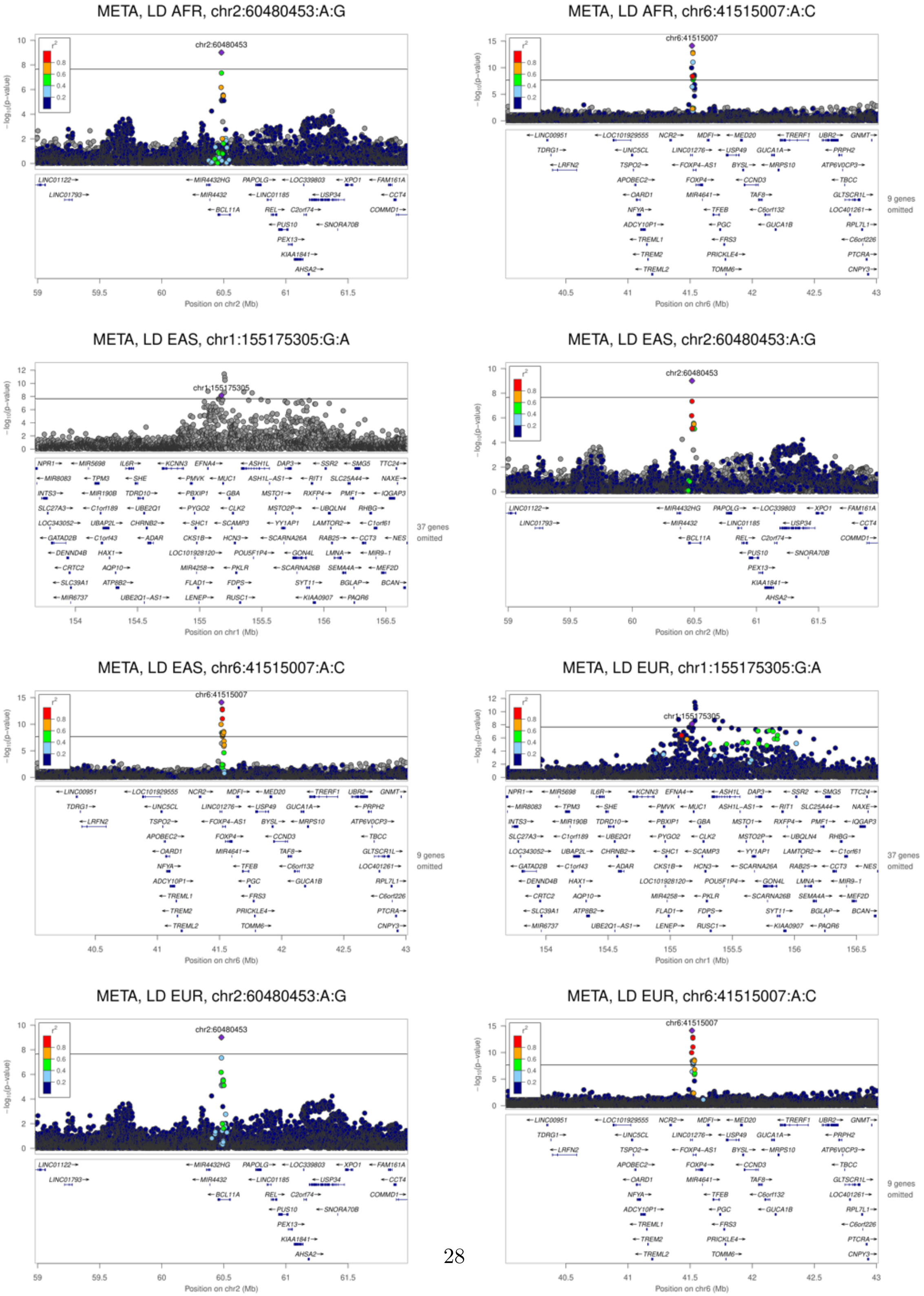

META, LD AFR, chr2:60480453:A:G
META, LD AFR, chr6:41515007:A:C
META, LD EAS, chr1:155175305:G:A
META, LD EAS, chr2:60480453:A:G
META, LD EAS, chr6:41515007:A:C
META, LD EUR, chr1:155175305:G:A
META, LD EUR, chr2:60480453:A:G
META, LD EUR, chr6:41515007:A:C

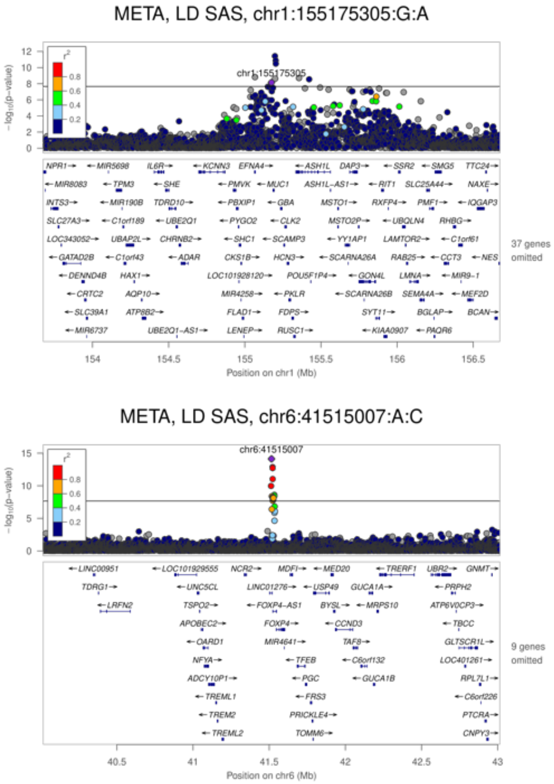

Figure 17: Locuszoom figures for the transethnic meta-analysis signals with different panels for LD calculated in the four populations of this study (AFR, EAS, EUR, SAS).

#### Stratified analysis

We performed sex- ($< 60$ vs. $\geq 60$) and age-stratified analyses. We did not obtain significant evidence for sex- specific effects (Supplementary Figure 18). The locus at chr3:45801750:G:A (rs13071258) in the European population had a significantly stronger effect in the younger age group ($OR = 3.34, 95\%CI = 2.98 - 3.75$ vs. $OR = 2.1, 95\%CI = 1.88 - 2.34$).

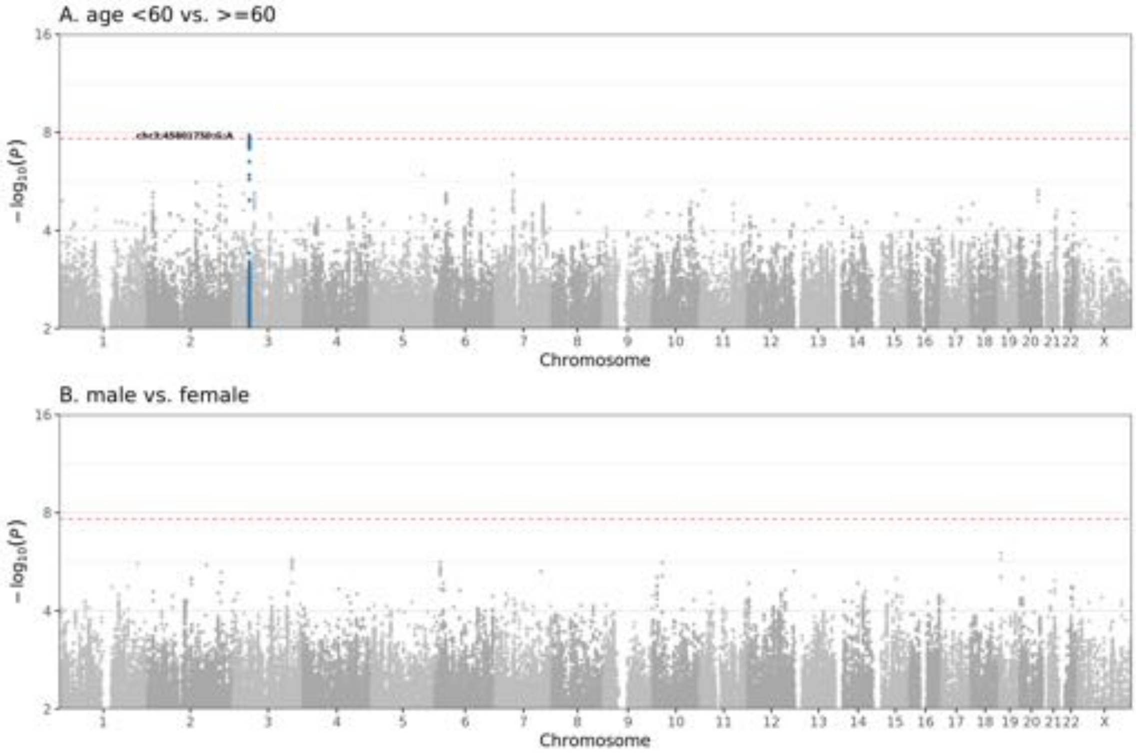

Figure 18: Manhattan plot for $t$-test $P$-values obtained from comparison of stratified GWAS analyses by age and sex. GWAS analyses were run for stratified subsets of the severe vs. mild+100K analysis for individuals with (A) age $>60$ vs. $\geq 60$ and (B) males vs. females. For each analysis we then perfomed a $t$-test comparing between-group effect sizes per variant. Red dashed line corresponds to Bonferroni-corrected $P$-value $= 2 \times 10^{-8}$.

### TWAS

#### eQTL TWAS

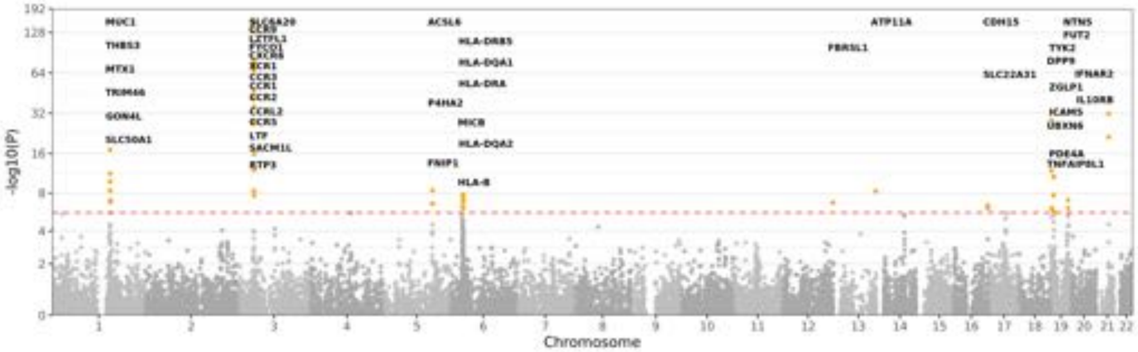

Figure 19: TWAS results from meta-analysis of eQTL models from all tissues in GTEXv8. All significant genes at $P < 2.3 \times 10^{-6}$ (red dashed line) are annotated.

#### Colocalisation sensitivity analysis

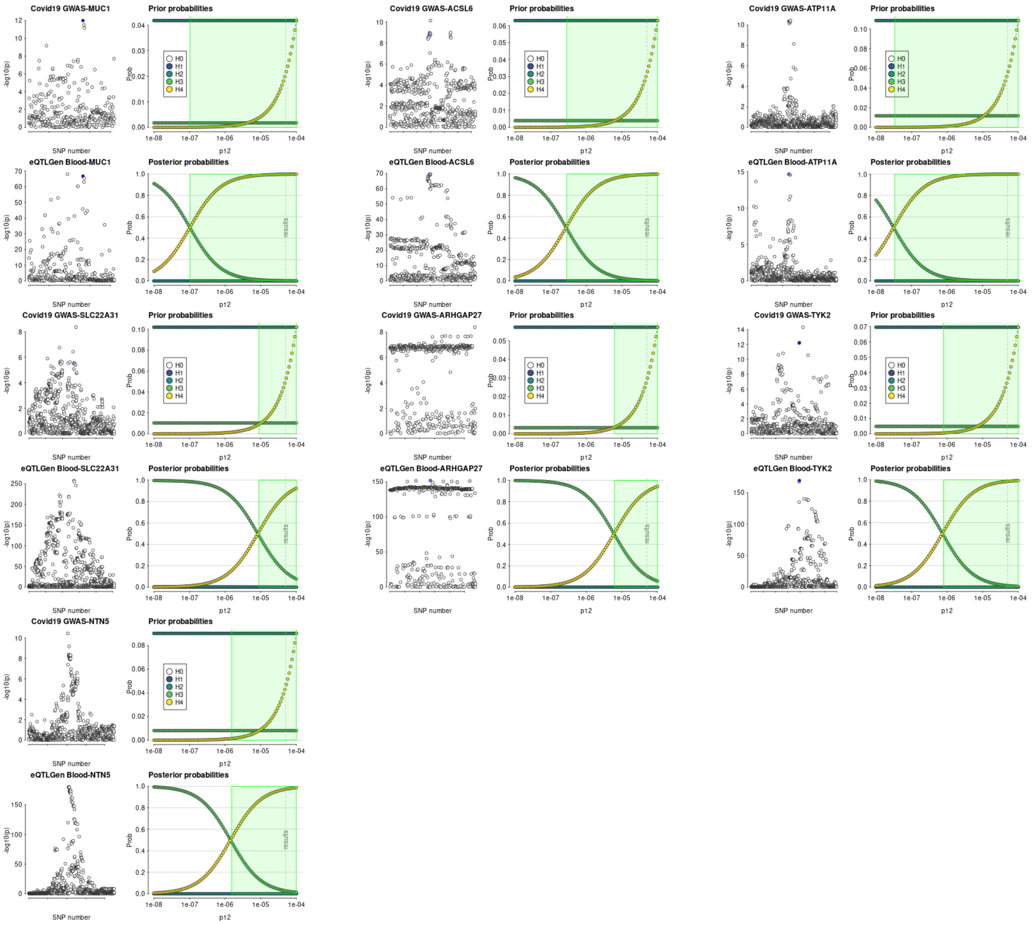

Figure 20: Sensitivity analysis for colocalisation of TWAS-significant loci in blood (eQTLgen data) with GWAS signals. Evidence for the following hypotheses is plotted across a range of prior probabilities: H0 - neither trait has a genetic association in the region; H1 - only trait 1 has a genetic association in the region; H2 - only trait 2 has a genetic association in the region; H3 - both traits are associated, but with different causal variants; H4 - both traits are associated and share a single causal variant.

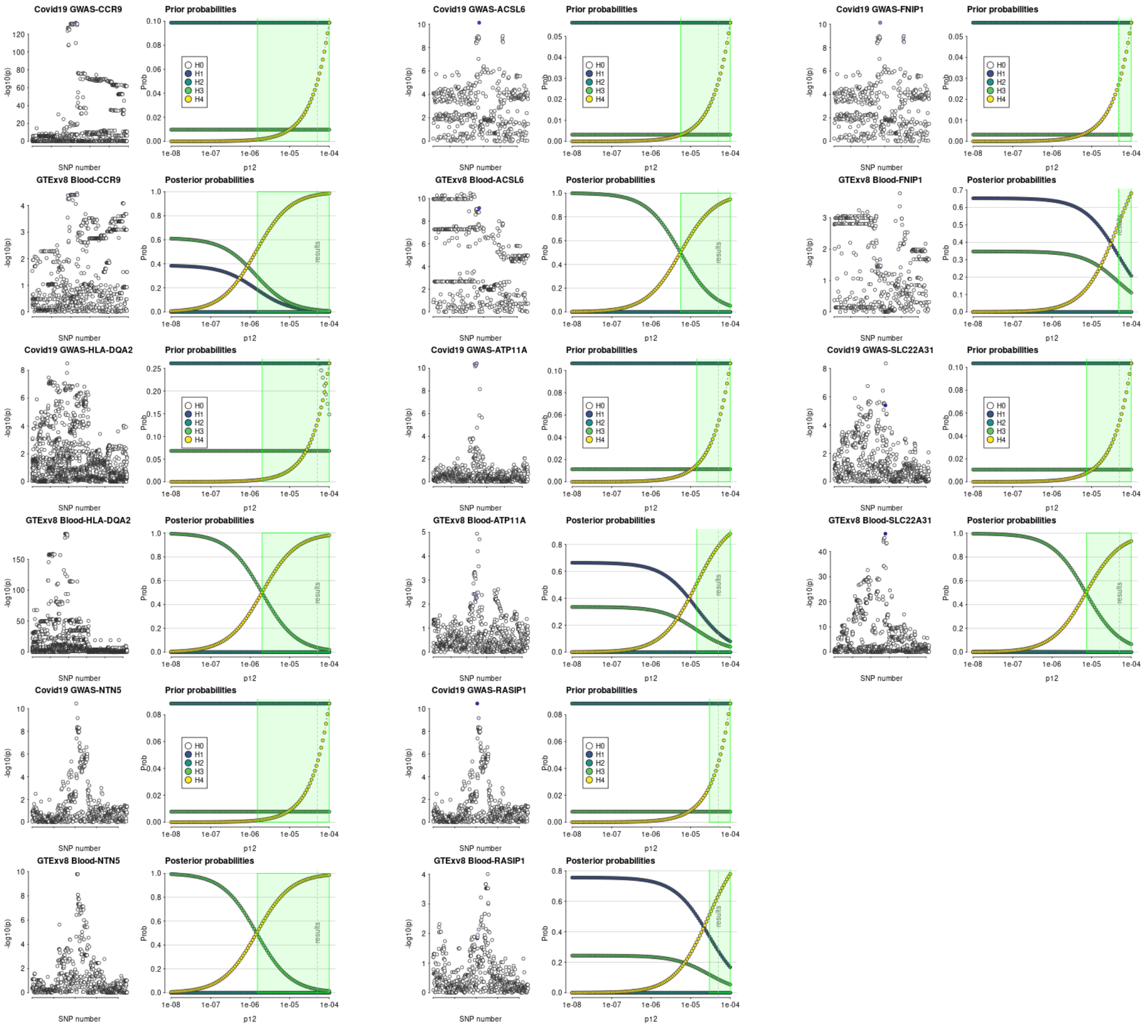

Figure 21: Sensitivity analysis for colocalisation of TWAS-significant loci in blood (GTEXv8 data) with GWAS signals. Evidence for the following hypotheses is plotted across a range of prior probabilities: H0 - neither trait has a genetic association in the region; H1 - only trait 1 has a genetic association in the region; H2 - only trait 2 has a genetic association in the region; H3 - both traits are associated, but with different causal variants; H4 - both traits are associated and share a single causal variant.

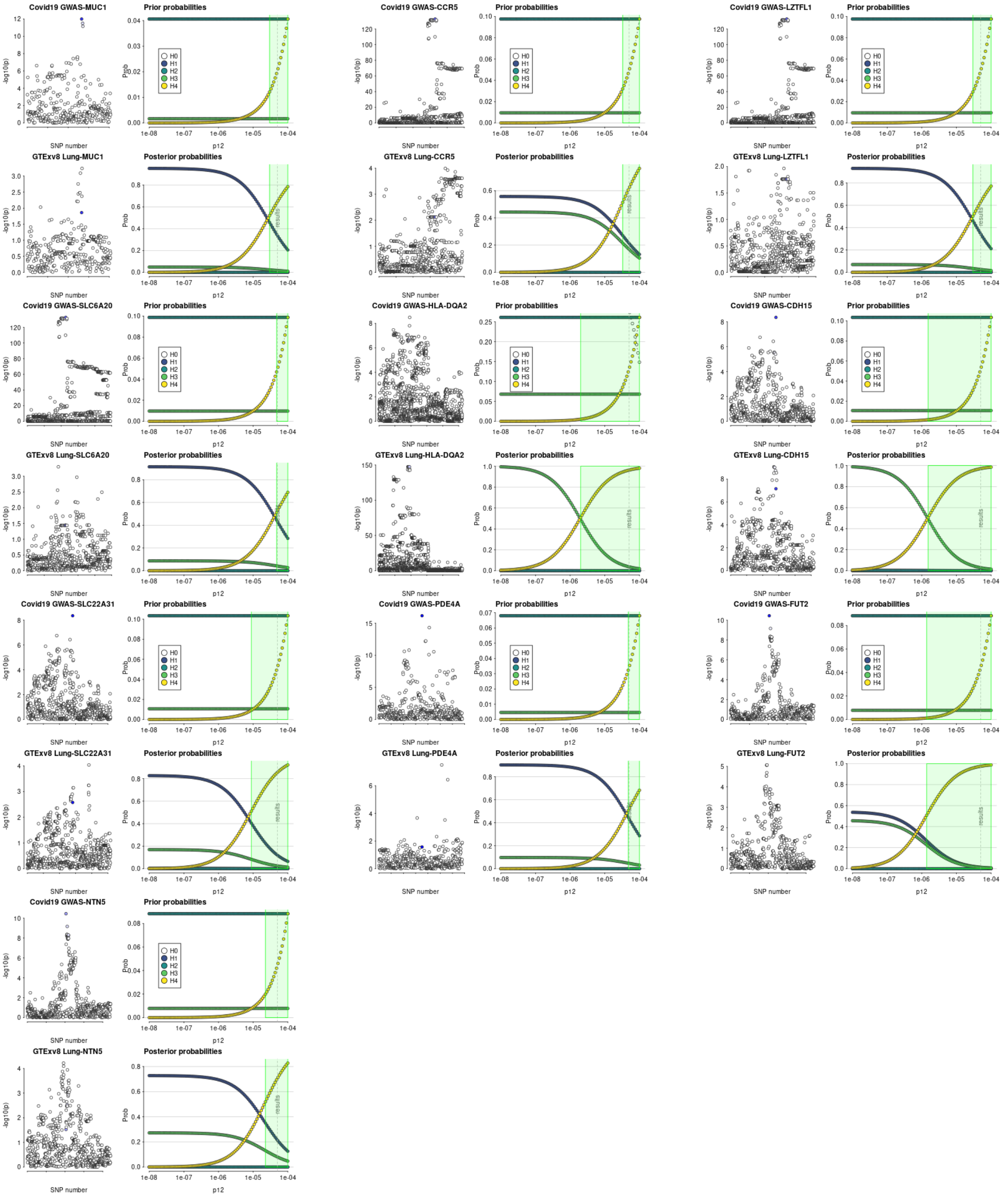

Figure 22: Sensitivity analysis for colocalisation of TWAS-significant loci in lung (GTEXv8 data) with GWAS signals. Evidence for the following hypotheses is plotted across a range of prior probabilities: H0 - neither trait has a genetic association in the region; H1 - only trait 1 has a genetic association in the region; H2 - only trait 2 has a genetic association in the region; H3 - both traits are associated, but with different causal variants; H4 - both traits are associated and share a single causal variant.

### Aggregate variant testing (AVT)

Aggregate variant testing on aggCOVID_v4.2 was performed using SKAT-O as implemented in SAIGE-GENE v0.44.5[6]. Variant and sample QC for the preparation of the aggregate files has been described elsewhere. In addition, the following filters were applied to the masked aggregate dataset:

- Bi-allelic SNPs only
- Minor allele frequency $< 0.005$
- Site wide missingness $< 0.05$
- Differential missingness between cases and controls, mid-p value $< 10^{-5}$

All the variants in the dataset were annotated using VEP v99.

#### Masks and Model

Two functional annotation masks were applied on top of the filters detailed. The first is a strict putative loss of function (*pLoF*) filter, where only variants that are annotated by Loftee as high confidence loss of function are included. The second is a more lenient filter (*missense*) where all variants from the strict filter are included, together with all variants that have a consequence of missense or worse as annotated by VEP, with a CADD_PHRED score of $\geq 10$ (CADD version 1.5). The covariates used in the model were the same as for the single variant analysis: $sex$, $age$, $age^2$, $age * sex$ and 20 (population-specific) principal components generated from common variants (MAF $\geq 5\%$).

The tests were run separately by genetically predicted ancestry, on all protein-coding genes as annotated by Ensembl.

#### AVT results

Supplementary table 4 shows the number of tested genes per mask per predicted ancestry. These numbers were used to apply a Bonferonni correction on the SKAT-O $P$-values from SAIGE-GENE on a per population basis. The $P$-value thresholds for gene-wide significance were taken as $0.05/n*2$, with $n$ being the number of tested genes in that population, divided by 2 (the number of masks used). This makes the assumption that each gene was tested by both masks, which is conservative for the missense threshold.

| **Cohort** | **Tested genes, pLof mask** | **$P$-value threshold, pLof mask** | **Tested genes, missense mask** | **$P$-value threshold, missense mask** |
|---|---|---|---|---|
| EUR | 7,352 | 3.4e-06 | 18,631 | 1.3e-06 |
| SAS | 1,435 | 1.7e-05 | 17,291 | 1.4e-06 |
| AFR | 763 | 3.3e-05 | 16,125 | 1.6e-06 |
| EAS | 265 | 9.4e-05 | 12,519 | 2.0e-06 |

Table 4: Number of tested genes per mask per predicted ancestry, with $P$-values used to assess gene-wide significance.

No significant associations were found across any of the populations. Supplementary figure 23 shows the Manhattan and Q-Q plots for each predicted ancestry using the *pLoF* mask, and supplementary figure 24 shows the Manhattan and Q-Q plots for each predicted ancestry using the *missense* mask. Supplementary File AVTsuppinfo.xlsx, sheet A and sheet B, show the top ten genes by $P$-value for each predicted ancestry and all combined ancestries, respectively.

Supplementary File AVTsuppinfo.xlsx, sheet C, shows the top genes that were highlighted as part of the GWAS analysis, ranked by p value per predicted ancestry. Supplementary File AVTsuppinfo.xlsx, sheet D shows the SKAT-O $P$-values for the 13 genes involved in the regulation of type I and III interferon immunity that were implicated in severe COVID-19 pneumonia[7], ranked by p value per predicted ancestry.

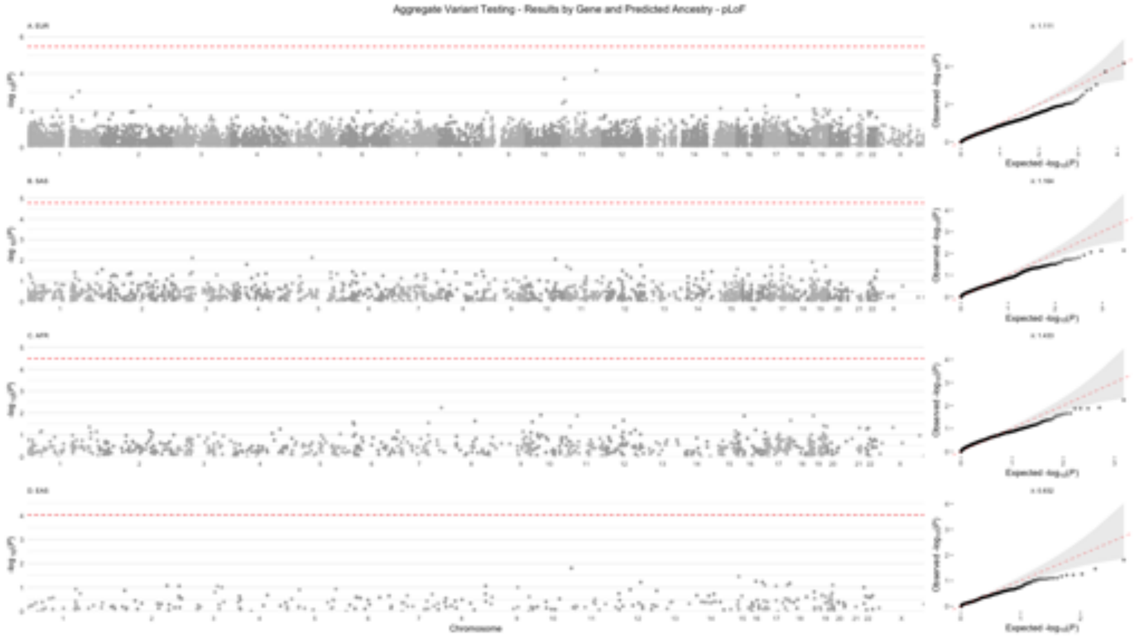

Figure 23: Gene-level manhattan and Q-Q plots per predicted ancestry for the *pLof* mask. Each point in the manhattan plot represents a gene. Panels from top to bottom are for EUR (A), SAS (B), AFR (C) and EAS (D). The red dashed lines indicate Bonferonni corrected "gene-wide" $P$-values (see Table 4).

### HLA Inference and Association Tests

#### HLA imputation using HIBAG

HLA types were imputed at two field (4-digit) resolution for all samples within aggV2 and aggCOVID_v4.2 for the following seven loci: HLA-A, HLA-C, HLA-B, HLA-DRB1, HLA-DQA1, HLA-DQB1, and HLA-DPB1 using the HIBAG package in R[8]. We used ancestry specific pre-fit classifiers trained on the Illumina 1M Duo genotyping array on individuals of either European, Asian, and African ancestry dependent on the assigned ancestry of the sample in hand. The list of HLA alleles represented in the reference panel is shown in Supplementary File HLAsuppinfo.xlsx

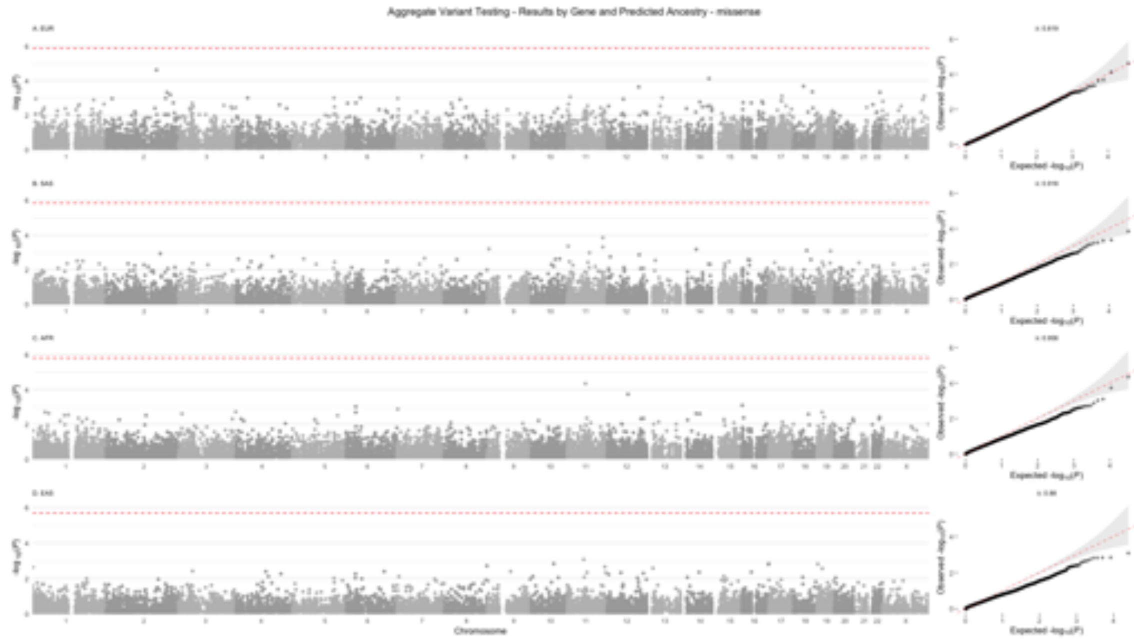

Figure 24: Gene-level manhattan and Q-Q plots per predicted ancestry for the *missense* mask. Each point in the manhattan plot represents a gene. Panels from top to bottom are for EUR (A), SAS (B), AFR (C) and EAS (D). The red dashed lines indicate Bonferonni corrected "gene-wide" $P$-values (see Table 4).

(Sheet A). HIBAG requires genotyped data in PLINK format as input. We lifted over the GRCh38 variant calls from aggV2 and aggCOVID_v4.2 for the extended (xMHC) region to hg19, keeping the variants included in the pre-trained classifiers for the seven HLA loci which were present in both the aggV2 and aggCOVID_v4.2 call-sets to ensure that the variants used for the imputation were the same across the two datasets. We applied a threshold of $T$=0.5 on the posterior probabilities returned by HIBAG, as in the original publication.

#### HLA inference using HLA*LA and concordance between HIBAG and HLA*LA callsets

We used a second HLA inference method, HLA*LA [9], to assess concordance and ensure call rates were comparable between the two methods. HLA*LA (version fe00f82) was used with GRCh38 IMGT population reference graphs to infer classical HLA types at G-group resolution for the three class I genes (HLA-A, HLA-C, HLA-B) and four class II genes (HLA-DRB1, HLA-DQA1, HLA-DQB1, HLA-DPB1) that were also umputed with HIBAG. HLA*LA implements a graph alignment model for HLA type inference, based on the projection of linear alignments onto a variation graph. Whole-genome sequencing BAM/CRAM files including unmapped reads were used as input. Where CRAM files were used (alignments from aggCOVID_v4.2 cohort), the reference genome FASTA file used for the original alignment was also provided.

Note that at time of writing, only 82% of aggV2 and aggCOVID_v4.2 samples had their HLA types inferred by HLA*LA (Supplementary File HLAsuppinfo.xlsx (Sheet B)). All samples had their HLA types imputed using HIBAG.

For samples for which we had both HIBAG and HLA*LA calls (n=45,796), we compared the 4-digit resolution alleles called from HIBAG with the alleles called from HLA*LA. As HLA*LA calls alleles at G-group resolution, we took all 4-digit alleles belonging to each G-Group and compared these to the HIBAG calls. For example, if a sample is called as A*01:01:01G, the mapped HLA alleles at 4-digit resolution within HLA*LA are 01:01, 01:04, 01:10, 01:13, 01:14, 01:15, 01:22, 01:32, 01:37, 01:45, 01:56, 01:81, 01:87. These were compared against the HIBAG 4-digit calls. If the 4-digits matched exactly (in either diploid combination - i.e. 01:01 / 01:04 vs 01:04 / 01:01), then sample alleles were deemed concordant. We found that >96% of calls were identical between HIBAG and HLA*LA.

The percentage of concordant calls between HIBAG and HLA*LA by ancestry and locus is shown in Supplementary File HLAsuppinfo.xlsx (Sheet C).

#### HLA Association Tests

HLA calls from HIBAG were aggregated into a single multi-sample VCF file containing sample genotypes for all observed HLA calls. Per sample, the genotypes of any allele call with posterior probability $< 0.5$ were set to missing. If a sample then had a missing genotype for a particular allele, all other alleles at that locus were also set to missing. For each locus, samples that did not harbour a specific HLA allele (either in a heterozygous or homozygous alternate state), were set to homozygous reference; unless already set to missing from the above mask.

HLA association analysis (single variant association tests) was run under an additive model using SAIGE (logistic mixed-model regression) version 0.44.5; in an identical fashion to the SNV GWAS.

The multi-sample VCF of aggregated HLA type calls from HIBAG were used as input. The set of 60K high-quality common SNPs aggCOVID_v4.2_aggV2_HQSNPs were used to create the GRM and variance ratio files. HLA association tests were run per ancestry (EUR, SAS, EAS, AFR) on unrelated individuals for the sev_vs_mld_aggV2 cohort. Each HLA association test was run using: $sex$, $age$, $age^2$, $age \times sex$, and the first 20 ancestral principle components as covariates. No minimum minor allele count / frequency threshold was set. Results can be seen in Supplementary File HLAsuppinfo.xlsx (Sheet D). Note this table combines the results for all ancestries (EUR, SAS, EAS, and AFR) - which is referenced in the first column of the table. The table is sorted by ancestry and alphabetically by allele.

HLA-DRB1*:04:01 was the only genome-wide significant HLA allele ($OR = 0.80, 95\%CI = 0.75 - 0.86, P = 1.6 \times 10^{-10}$, in EUR), having a protective effect (casesMAF: 9.6%, controlsMAF: 11.7%). In Europeans, the DRB1*04:01 allele had a low rate of missingness (call rate >0.92 at T=0.5 posterior probability threshold), and was in Hardy-Weinberg equilibrium for both cases (p=0.32) and controls (p=0.17). The observed allele frequency for HLA-DRB1*04:01 was 0.1%, 0.08%, and 0.02% for AFR, EAS, and SAS cohorts respectively. A meta-analysis was performed using METAL with an inverse-variance weighted method across the four populations. DRB1*04:01 remained the only significant association ($OR = 0.80, 95\%CI = 0.75 - 0.86, P = 1.4 \times 10^{-10}$), Supplementary File HLAsuppinfo.xlsx (Sheet E).

#### Conditional Analysis

We conducted a conditional analysis, controlling for the HLA-DRB1*04:01 allele, on the main GWAS results within the extended MHC region. This analysis was performed on the European sev_vs_mld_aggV2 cohort. To do this, we firstly regressed out the effect of DRB1*04:01 (including $age$, $sex$, $age \times sex$, $age^2$, and the first 20 population PCs), and performed linear regression on the residuals using the GWAS variant genotypes as the dependent variable. No variants within the extended MHC remained genome-wide significant upon conditioning on DRB1*04:01. The top GWAS signal (chr6:32623820 T/C; $OR = 0.88, 95\%CI = 0.84 - 0.92, P = 3.3 \times 10^{-9}$) was attenuated following conditional analysis ($P = 0.001$). Figure 25 shows the results of the combined HLA and GWAS association results and the conditional analysis.

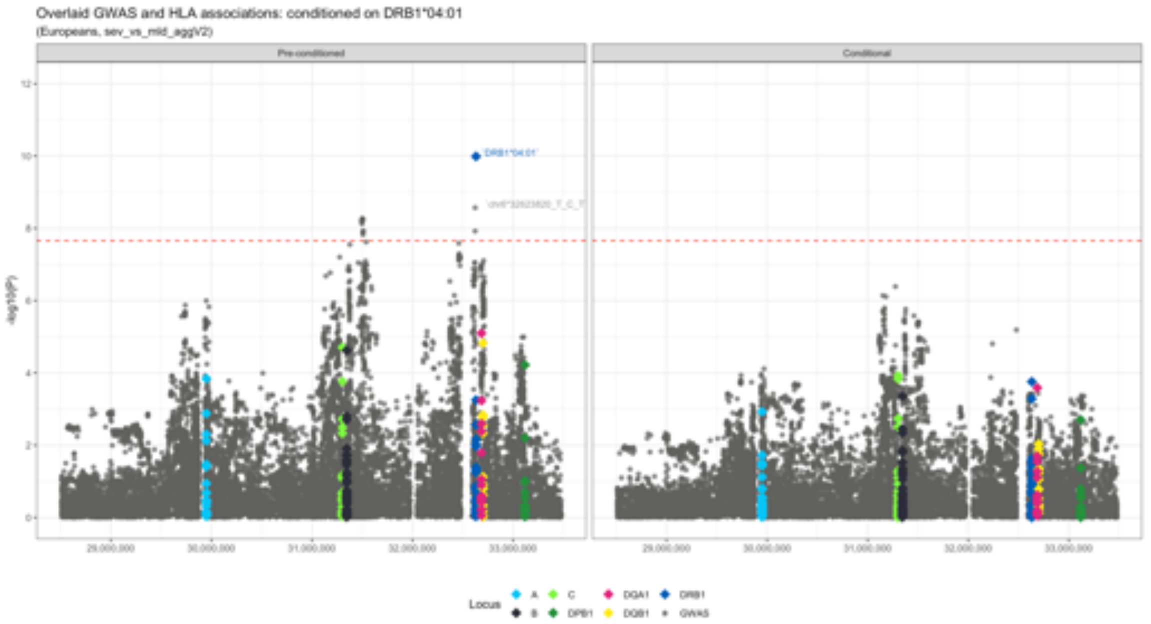

Figure 25: Manhattan plot of HLA and GWAS signal across the extended MHC region for the European sev_vs_mld_aggV2 cohort. Grey circles mark the GWAS (small variant) associations and diamonds represent the HLA each allele association, coloured by locus. The lead variant from the GWAS and lead allele from HLA are labelled. The left-panel shows the raw association -log10(p-values) per variant - prior to conditional analysis. The right-panel shows the -log10(p-values) per variant following conditioning on DRB1*04:01. The dashed red line is the genome-wide significance threshold for Europeans.

### Meta-analysis by information content (MAIC)

In order to put the results in the context of existing knowledge of host genes implicated in SARS-CoV-2 replication or pathophysiology of COVID-19, we use meta-analyis by information content (MAIC)[10] to incorporate lists of named genes from a large systematic review of in vitro and in vivo studies.[11] Remarkably, the top 2000 named genes in our metaTWAS contributes 19.5% of the total information content in this composite analysis (Supplementary Figure 26). Full results are available at baillielab.net/maic/covid.

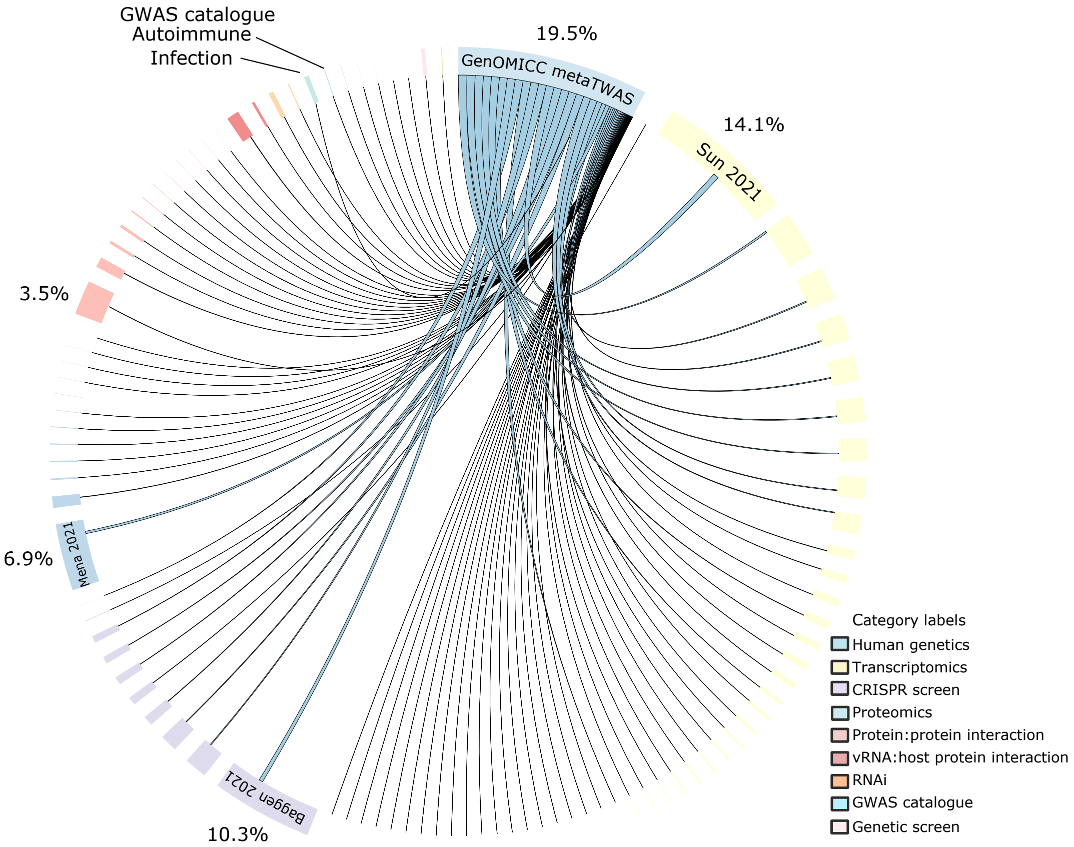

Figure 26: Circular diagram of shared information content among data sources using MAIC analysis. Each data source is represented by a coloured block on the outer ring of the circle; the size of data source blocks is proportional to the summed information content of the input list—that is, the total contribution that this data source makes to the aggregate, calculated as the sum of the MAIC gene scores contributed by that list and represented numerically for datasets with the highest information content. Lines are coloured according to the dominant data source. Data sources within the same category share the same colour (legend). The largest categories and data sources are labelled. An interactive version of this figure is available at baillielab.net/maic/covid.

### GenOMICC Investigators

**GenOMICC Co-Investigator** J. Kenneth Baillie[1,2], Colin Begg[3], Sara Clohisey[1], Charles Hinds[4], Peter Horby[5], Julian Knight[6], David Maslove[7], Danny McAuley[8,9], Johnny Millar[1], Hugh Montgomery[10], Alistair Nichol[11], Peter J.M. Openshaw[12,13], Chris P Ponting[14], Kathy Rowan[15], Malcolm G Semple[16,17], Manu Shankar-Hari[18], Charlotte Summers[19], Timothy Walsh[2].

**Management and Laboratory team** Ruth Armstrong[1], J. Kenneth Baillie[1,2], Heather Biggs[20], Ceilia Boz[1], Adam Brown[1], Richard Clark[21], Sara Clohisey[1], Audrey Coutts[21], Judy Coyle[1], Louise Cullum[1], Nicky Day[1], Lorna Donnelly[21], Esther Duncan[1], Angie Fawkes[21], Paul Finernan[1], Max Head Fourman[1], Anita Furlong[20], James Furniss[1], Bernadette Gallagher[1], Tammy Gilchrist[21], Ailsa Golightly[1], Fiona Griffiths[1], Katarzyna Hafezi[21], Debbie Hamilton[1], Ross Hendry[1], Andy Law[1], Dawn Law[1], Rachel Law[1], Sarah Law[1], Rebecca Lidstone-Scott[1], Louise Macgillivray[21], Alan Maclean[21], Hanning Mal[1], Sarah McCafferty[21], Ellie Mcmaster[1], Jen Meikle[1], Shona C Moore[16], Kirstie Morrice[21], Lee Murphy[21], Wilna Oosthuyzen[1], Nick Parkinson[1], Trevor Paterson[1], Katherine Schon[20], Andrew Stenhouse[1], Maaike Swets[1,22], Helen Szoor-McElhinney[1], Filip Taneski[1], Lance Turtle[16], Tony Wackett[1], Mairi Ward[1], Jane Weaver[1], Nicola Wrobel[21], Marie Zechner[1], Mybaya Hellen[1].

**Guys and St Thomas' Hospital, London, UK** Gill Arbane[23], Aneta Bociek[23], Sara Campos[‡,23], Neus Grau[23], Tim Owen Jones[23], Rosario Lim[23], Martina Marotti[23], Marlies Ostermann[*,23], Manu Shankar-Hari[*,23], Christopher Whitton[23].

**Barts Health NHS Trust, London, UK** Zoe Alldis[24], Raine Astin-Chamberlain[24], Fatima Bibi[24], Jack Biddle[24], Sarah Blow[24], Matthew Bolton[24], Catherine Borra[24], Ruth Bowles[24], Maudrian Burton[24], Yasmin Choudhury[24], David Collier[*,24], Amber Cox[24], Amy Easthope[24], Patrizia Ebano[24], Stavros Fotiadis[24], Jana Gurasashvili[24], Rosslyn Halls[24], Pippa Hartridge[24], Delordson Kallon[24], Jamila Kassam[24], Ivone Lancoma-Malcolm[24], Maninderpal Matharu[24], Peter May[24], Oliver Mitchelmore[24], Tabitha Newman[24], Mital Patel[24], Jane Pheby[24], Irene Pinzuti[24], Zoe Prime[24], Oleksandra Prysyazhna[24], Julian Shiel[24], Melanie Taylor[24], Carey Tierney[24], Suzanne Wood[‡,24], Anne Zak[‡,24], Olivier Zongo[24].

**James Cook University Hospital, Middlesbrough, UK** Stephen Bonner[*,25], Keith Hugill[25], Jessica Jones[25], Steven Liggett[25], Evie Headlam[25].

**Royal Stoke University Hospital, Staffordshire, UK** Nageswar Bandla[*,26], Minnie Gellamucho[‡,26], Michelle Davies[26], Christopher Thompson[26].

**North Middlesex University Hospital NHS trust, London, UK** Marwa Abdelrazik[27], Dhanalakshmi Bakthavatsalam[‡,27], Munzir Elhassan[27], Arunkumar Ganesan[27], Anne Haldeos[*,27], Jeronimo Moreno-Cuesta[*,27], Dharam Purohit[27], Rachel Vincent[‡,27], Kugan Xavier[27], kumar Rohit[28], Frater Alasdair[27], Malik Saleem[27], Carter David[27], Jenkins Samuel[27], Zoe Lamond[27], Wall Alanna[27].

**The Royal Liverpool University Hospital, Liverpool, UK** Jaime Fernandez-Roman[29], David O. Hamilton[29], Emily Johnson[29], Brian Johnston[29], Maria Lopez Martinez[29], Suleman Mulla[29], David Shaw[29], Alicia A.C. Waite[29], Victoria Waugh[‡,29], Ingeborg D. Welters[*,29], Karen Williams[29].

**King's College Hospital, London, UK** Anna Cavazza[30], Maeve Cockrell[30], Eleanor Corcoran[30], Maria Depante[30], Clare Finney[30], Ellen Jerome[30], Mark McPhail[*,30], Monalisa Nayak[30], Harriet Noble[30], Kevin O'Reilly[30], Evita Pappa[30], Rohit Saha[30], Sian Saha[30], John Smith[30], Abigail Knighton[30].

**Charing Cross Hospital, St Mary's Hospital and Hammersmith Hospital, London, UK** David Antcliffe[*,31], Dorota Banach[31], Stephen Brett[31], Phoebe Coghlan[31], Ziortza Fernandez[31], Anthony Gordon[*,31], Roceld Rojo[31], Sonia Sousa Arias[31], Maie Templeton[‡,31].

**Nottingham University Hospital, Nottingham, UK** Megan Meredith[*,32], Lucy Morris[*,32], Lucy Ryan[32], Amy Clark[32], Julia Sampson[32], Cecilia Peters[32], Martin Dent[32], Margaret Langley[32], Saima Ashraf[32], Shuying Wei[32], Angela Andrew[32].

**John Radcliffe Hospital, Oxford, UK** Archana Bashyal[33], Neil Davidson[33], Paula Hutton[‡,33], Stuart McKechnie[*,33], Jean Wilson[33].

**Kingston Hospital, Surrey, UK** David Baptista[34], Rebecca Crowe[34], Rita Fernandes[34], Rosaleen Herdman-Grant[‡,34], Anna Joseph[*,34], Adam Loveridge[34], India McKenley[34], Eriko Morino[34], Andres Naranjo[34], Richard Simms[34], Kathryn Sollesta[34], Andrew Swain[34], Harish Venkatesh[34], Jacyntha Khera[34], Jonathan Fox[34].

**Royal Infirmary of Edinburgh, Edinburgh, UK** Gillian Andrew[35], J. Kenneth Baillie[*,35], Lucy Barclay[‡,35], Marie Callaghan[35], Rachael Campbell[35], Sarah Clark[35], Dave Hope[35], Lucy Marshall[35], Corrienne McCulloch[35], Kate Briton[35], Jo Singleton[35], Sohphie Birch [35].

**Queen Alexandra Hospital, Portsmouth, UK** Lutece Brimfield[36], Zoe Daly[36], David Pogson[*,36], Steve Rose[‡,36].

**Morriston Hospital, Swansea, UK** Ceri Battle[*,37], Elaine Brinkworth[37], Rachel Harford[37], Carl Murphy[37], Luke Newey[*,37], Tabitha Rees[‡,37], Marie Williams[37], Sophie Arnold[37].

**Addenbrooke's Hospital, Cambridge, UK** Petra Polgarova[‡,38], Katerina Stroud[38], Charlotte Summers[*,38], Eoghan Meaney[38], Megan Jones[38], Anthony Ng[38], Shruti Agrawal[38], Nazima Pathan[38], Deborah White[38], Esther Daubney[38], Kay Elston[38].

**BHRUT (Barking Havering) - Queens Hospital and King George Hospital, Essex, UK** Lina Grauslyte[39], Musarat Hussain[*,39], Mandeep Phull[*,39], Tatiana Pogreban[‡,39], Lace Rosaroso[‡,39], Erika Salciute[39], George Franke[39], Joanna Wong[39], Aparna George[39].

**Royal Sussex County Hospital, Brighton, UK** Laura Ortiz-Ruiz de Gordoa[40], Emily Peasgood[40], Claire Phillips[*,40], Laura Ortiz-Ruiz de Gordoa[40], Emily Peasgood[40], Claire Phillips[*,40].

**Queen Elizabeth Hospital, Birmingham, UK** Michelle Bates[41], Jo Dasgin[41], Jaspret Gill[41], Annette Nilsson[41], James Scriven[*,41].

**St George's Hospital, London, UK** Carlos Castro Delgado[42], Deborah Dawson[42], Lijun Ding[42], Georgia Durrant[42], Obiageri Ezeobu[42], Sarah Farnell-Ward[*,42], Abiola Harrison[42], Rebecca Kanu[42], Susannah Leaver[*,42], elena Maccacari[‡,42], Soumendu Manna[*,42], Romina Pepermans Saluzzio[42], Joana Queiroz[‡,42], Tinashe Samakomva[42], Christine Sicat[42], Joana Texeira[42], Edna Fernandes Da Gloria[42], Ana Lisboa[42], John Rawlins[42], Jisha Mathew[42], Ashley Kinch[42], William James Hurt[42], Nirav Shah[42], Victoria Clark[42], Maria Thanasi[42], Nikki Yun[42], Kamal Patel[42].

**Stepping Hill Hospital, Stockport, UK** Sara Bennett[43], Emma Goodwin[43], Matthew Jackson[*,43], Alissa Kent[43], Clare Tibke[‡,43], Wiesia Woodyatt[43], Ahmed Zaki[*,43].

**Countess of Chester Hospital, Chester, UK** Azmerelda Abraheem[44], Peter Bamford[*,44], Kathryn Cawley[‡,44], Charlie Dunmore[44], Maria Faulkner[‡,44], Rumanah Girach[44], Helen Jeffrey[‡,44], Rhianna Jones[44], Emily London[44], Imrun Nagra[44], Farah Nasir[44], Hannah Sainsbury[44], Clare Smedley[44].

**Royal Blackburn Teaching Hospital, Blackburn, UK** Tahera Patel[45], Matthew Smith[45], Srikanth Chukkambotla[45], Aayesha Kazi[45], Janice Hartley[45], Joseph Dykes[45], Muhammad Hijazi[45], Sarah Keith[45], Meherunnisa Khan[45], Janet Ryan-Smith[45], Philippa Springle[45], Jacqueline Thomas[45], Nick Truman[45], Samuel Saad[45], Dabheoc Coleman[45], Christopher Fine[45], Roseanna Matt[45], Bethan Gay[45], Jack Dalziel[45], Syamlan Ali[45], Drew Goodchild[45], Rhiannan Harling[45], Ravi Bhatterjee[45], Wendy Goddard[45], Chloe Davison[45], Stephen Duberly[45], Jeanette Hargreaves[45], Rachel Bolton[45].

**The Tunbridge Wells Hospital and Maidstone Hospital, Kent, UK** Miriam Davey[*,46], David Golden[*,46], Rebecca Seaman[46].

**Royal Gwent Hospital, Newport, UK** Shiney Cherian[‡,47], Sean Cutler[47], Anne Emma Heron[‡,47], Anna Roynon-Reed[47], Tamas Szakmany[*,47], Gemma Williams[47], Owen Richards[47], Yusuf Cheema[47].

**Pinderfields General Hospital, Wakefield, UK** Hollie Brooke[48], Sarah Buckley[‡,48], Jose Cebrian Suarez[48], Ruth Charlesworth[48], Karen Hansson[48], John Norris[48], Alice Poole[48], Alastair Rose[*,48], Rajdeep Sandhu[48], Brendan Sloan[*,48], Elizabeth Smithson[48], Muthu Thirumaran[48], Veronica Wagstaff[48], Alexandra Metcalfe[48].

**Royal Berkshire NHS Foundation Trust, Berkshire, UK** Mark Brunton[49], Jess Caterson[49], Holly Coles[49], Matthew Frise[*,49], Sabi Gurung Rai[49], Nicola Jacques[‡,49], Liza Keating[49], Emma Tilney[49], Shauna Bartley[49], Parminder Bhuie[49].

**Broomfield Hospital, Chelmsford, UK** Sian Gibson[50], Amanda Lyle[50], Fiona McNeela[50], Jayachandran Radhakrishnan[*,50], Alistair Hughes[*,50].

**Northumbria Healthcare NHS Foundation Trust, North Shields, UK** Bryan Yates[*,51], Jessica Reynolds[‡,51], Helen Campbell[51], Maria Thompsom[51], Steve Dodds[51], Stacey Duffy[51].

**Whiston Hospital, Prescot, UK** Sandra Greer[‡,52], Karen Shuker[‡,52], Ascanio Tridente[*,52].

**Croydon University Hospital, Croydon, UK** Reena Khade[‡,53], Ashok Sundar [*,53], George Tsinaslanidis[53].

**York Hospital, York, UK** Isobel Birkinshaw[‡,54], Joseph Carter[*,54], Kate Howard[54], Joanne Ingham[54], Rosie Joy[54], Harriet Pearson[54], Samantha Roche[54], Zoe Scott[54].

**Heartlands Hospital, Birmingham, UK** Hollie Bancroft[55], Mary Bellamy[55], Margaret Carmody[55], Jacqueline Daglish[55], Faye Moore[55], Joanne Rhodes[55], Mirriam Sangombe[55], Salma Kadiri[55], James Scriven[*,55].

**Ashford and St Peter's Hospital, Surrey, UK** Maria Croft[‡,56], Ian White[*,56], Victoria Frost[56], Maia Aquino[56].

**Barnet Hospital, London, UK** Rajeev Jha[*,57], Vinodh Krishnamurthy[57], Lai Lim[57], Rajeev Jha[*,57], Vinodh Krishnamurthy[57], Li Lim[‡,57].

**East Surrey Hospital, Redhill, UK** Edward Combes[*,58], Teishel Joefield[58], Sonja Monnery[58], Valerie Beech[58], Sallyanne Trotman[58].

**Ninewells Hospital, Dundee, UK** Christine Almaden-Boyle[59], Pauline Austin[59], Louise Cabrelli[‡,59], Stephen Cole[59], Matt Casey[59], Susan Chapman[59], Stephen Cole[*,59], Clare Whyte[59].

**Worthing Hospital, Worthing, UK and St Richard's Hospital, Chichester, UK** Yolanda Baird[‡,60], Aaron Butler[60], Indra Chadbourn[60], Linda Folkes[60], Heather Fox[60], Amy Gardner[60], Raquel Gomez[‡,60], Gillian Hobden[60], Luke Hodgson[*,60], Kirsten King[60], Michael Margarson[*,60], Tim Martindale[60], Emma Meadows[60], Dana Raynard[60], Yvette Thirlwall[60], David Helm[60], Jordi Margalef[60].

**Southampton General Hospital, Southampton, UK** Kristine Criste[61], Rebecca Cusack[*,61], Kim Golder[*,61], Hannah Golding[61], Oliver Jones[‡,61], Samantha Leggett[61], Michelle Male[61], Martyna Marani[61], Kirsty Prager[61], Toran Williams[61], Belinda Roberts[61], Karen Salmon[61].

**The Alexandra Hospital, Redditch and Worcester Royal Hospital, Worcester, UK** Peter Anderson[62], Katie Archer[62], Karen Austin[62], caroline Davis[62], Alison Durie[62], Olivia Kelsall[*,62], Jessica Thrush[62], Charlie Vigurs[62], Laura Wild[62], Hannah-Louise Wood[62], Helen Tranter[62], Alison Harrison[62], Nicholas Cowley[62], Michael McAlindon[62], Andrew Burtenshaw[62], Stephen Digby[62], Emma Low[62], Aled Morgan[62], Naiara Cother[62], Tobias Rankin[62], Sarah Clayton[62], Alex McCurdy[62].

**Sandwell General Hospital and City Hospital, Birmingham, UK** Cecilia Ahmed[63], Balvinder Baines[63], Sarah Clamp[63], Julie Colley[63], Risna Haq[63], Anne Hayes[63], Jonathan Hulme[*,63], Samia Hussain[‡,63], Sibet Joseph[63], Rita Kumar[63], Zahira Maqsood[63], Manjit Purewal[63].

**Blackpool Victoria Hospital, Blackpool, UK** Leonie Benham[‡,64], Zena Bradshaw[64], Joanna Brown[64], Melanie Caswell[64], Jason Cupitt[*,64], Sarah Melling[64], Stephen Preston[64], Nicola Slawson[64], Emma Stoddard[‡,64], Scott Warden[64].

**Royal Glamorgan Hospital, Pontyclun, UK** Bethan Deacon[65], Ceri Lynch[*,65], Carla Pothecary[65], Lisa Roche[‡,65], Gwenllian Sera Howe[65], Jayaprakash Singh[65], Keri Turner[65], Hannah Ellis[65], Natalie Stroud[65].

**The Royal Oldham Hospital, Manchester, UK** Jodie Hunt[66], Joy Dearden[‡,66], Emma Dobson[66], Andy Drummond[*,66], Michelle Mulcahy[66], Sheila Munt[66], Grainne O'Connor[66], Jennifer Philbin[66], Chloe Rishton[66], Redmond Tully[*,66], Sarah Winnard[66].

**Glasgow Royal Infirmary, Glasgow, UK** Susanne Cathcart[‡,67], Katharine Duffy[67], Alex Puxty[*,67], Kathryn Puxty[67], Lynne Turner[67], Jane Ireland[67], Gary Semple[67].

**St James's University Hospital and Leeds General Infirmary, Leeds, UK** Kate Long[68], Simon Whiteley[*,68], Elizabeth Wilby[‡,68], Bethan Ogg[68].

**University Hospital North Durham, Darlington, UK and Darlington Memorial Hospital, Darlington, UK** Amanda Cowton[*,69], Andrea Kay[69], Melanie Kent[69], Kathryn Potts[69], Ami Wilkinson[69], Suzanne Campbell[*,69], Ellen Brown [‡,69].

**Fairfield General Hospital, Bury, UK** Julie Melville[70], Jay Naisbitt[*,70], Rosane Joseph[70], Maria Lazo[70], Olivia Walton[70], Alan Neal[70].

**Wythenshawe Hospital, Manchester, UK** Peter Alexander[*,71], Schvearn Allen[71], Joanne Bradley-Potts[‡,71], Craig Brantwood[*,71], Jasmine Egan[71], Timothy Felton[71], Grace Padden[71], Luke Ward[71], Stuart Moss[71], Susannah Glasgow [71].

**Royal Alexandra Hospital, Paisley, UK** Lynn Abel[‡,72], Michael Brett[72], Brian Digby[72], Lisa Gemmell[72], James Hornsby[72], Patrick MacGoey[72], Pauline O'Neil[72], Richard Price[72], Natalie Rodden[‡,72], Kevin Rooney[*,72], Radha Sundaram[72], Nicola Thomson[‡,72].

**Good Hope Hospital, Birmingham, UK** Bridget Hopkins[73], James Scriven[*,73], Laura Thrasyvoulou[73], Heather Willis[73].

**Tameside General Hospital, Ashton Under Lyne, UK** Martyn Clark[74], Martina Coulding[74], Edward Jude[*,74], Jacqueline McCormick[74], Oliver Mercer[74], Darsh Potla[74], Hafiz Rehman[74], Heather Savill[74], Victoria Turner[‡,74].

**Royal Derby Hospital, Derby, UK** Charlotte Downes[75], Kathleen Holding[*,75], Katie Riches[‡,75], Mary Hilton[75], Mel Hayman[75], Deepak Subramanian[*,75], Priya Daniel[*,75].

**Medway Maritime Hospital, Gillingham, UK** Oluronke Adanini[76], Nikhil Bhatia[*,76], Maines Msiska[‡,76].

**Royal Victoria Infirmary, Newcastle Upon Tyne, UK** Ian Clement[*,77], Bijal Patel [77], A Gulati[77], Carole Hays[‡,77], K Webster[77], Anne Hudson[‡,77], Andrea Webster[77], Elaine Stephenson[77], Louise McCormack[77], Victoria Slater[77], Rachel Nixon[77], Helen Hanson[77], Maggie fearby[77], Sinead Kelly[77], Victoria Bridgett[77], Philip Robinson[77].

**Poole Hospital, Poole, UK** Julie Camsooksai[‡,78], Charlotte Humphrey[78], Sarah Jenkins[78], Henrik Reschreiter[*,78], Beverley Wadams[78], Yasmin Death[78].

**Bedford Hospital, Bedford, UK** Victoria Bastion[79], Daphene Clarke[79], Beena David[‡,79], Harriet Kent[79], Rachel Lorusso[79], Gamu Lubimbi[79], Sophie Murdoch[79], Melchizedek Penacerrada[79], Alastair Thomas[79], Jennifer Valentine[79], Ana Vochin[*,79], Retno Wulandari[79], Brice Djeugam[79].

**Queens Hospital Burton, Burton-On-Trent, UK** Gillian Bell[‡,80], Katy English[*,80], Amro Katary[*,80], Louise Wilcox[‡,80].

**North Manchester General Hospital, Manchester, UK** Michelle Bruce[81], Karen Connolly[81], Tracy Duncan[*,81], Helen T-Michael[81], Gabriella Lindergard[81], Samuel Hey [81], Claire Fox[81], Jordan Alfonso[81], Laura Jayne Durrans[81], Jacinta Guerin[81], Bethan Blackledge[81], Jade Harris[81], Martin Hruska[81], Ayaa Eltayeb[81], Thomas Lamb[81], Tracey Hodgkiss[81], Lisa Cooper[81], Joanne Rothwell[81].

**Aberdeen Royal Infirmary, Aberdeen, UK** Angela Allan[82], Felicity Anderson[‡,82], Callum Kaye[*,82], Jade Liew[82], Jasmine Medhora[82], Teresa Scott[82], Erin Trumper[82], Adriana Botello[82].

**Derriford Hospital, Plymouth, UK** Liana Lankester[‡,83], Nikitas Nikitas[*,83], Colin Wells[83], Bethan Stowe[83], Kayleigh Spencer[83].

**Manchester Royal Infirmary, Manchester, UK** Craig Brandwood[*,84], Lara Smith[84], Richard Clark[84], Katie Birchall[84], Laurel Kolakaluri[84], Deborah Baines [84], Anila Sukumaran[84].

**Salford Royal Hospital, Manchester, UK** Elena Apetri[85], Cathrine Basikolo[85], Bethan Blackledge[85], Laura Catlow[85], Bethan Charles[‡,85], Paul Dark[85], Reece Doonan[85], Jade Harris[85], Alice Harvey[85], Daniel Horner[*,85], Karen Knowles[85], Stephanie Lee[85], Diane Lomas[85], Chloe Lyons[85], Tracy Marsden[85], Danielle McLaughlan[‡,85], Liam McMorrow[85], Jessica Pendlebury[85], Jane Perez[85], Maria Poulaka[85], Nicola Proudfoot[85], Melanie Slaughter[85], Kathryn Slevin[85], Melanie Taylor[85], Vicky Thomas[85], Danielle Walker[85], Angiy Michael [85], Matthew Collis[85].

**William Harvey Hospital, Ashford, UK** Tracey Cosier[86], Gemma Millen[86], Neil Richardson[*,86], Natasha Schumacher[86], Heather Weston[86], James Rand[86].

**Queen Elizabeth University Hospital, Glasgow, UK** Nicola Baxter[87], Steven Henderson[*,87], Sophie Kennedy-Hay[‡,87], Christopher McParland[87], Laura Rooney[87], Malcolm Sim[87], Gordan McCreath[87].

**Bradford Royal Infirmary, Bradford, UK** Louise Akeroyd[‡,88], Shereen Bano[88], Matt Bromley[88], Lucy Gurr[88], Tom Lawton[88], James Morgan[*,88], Kirsten Sellick[88], Deborah Warren[88], Brian Wilkinson[88], Janet McGowan[88], Camilla Ledgard[88], Amelia Stacey[88], Kate Pye[88], Ruth Bellwood[88], Michael Bentley[88].

**Bristol Royal Infirmary, Bristol, UK** Jeremy Bewley[*,89], Zoe Garland[89], Lisa Grimmer[89], Bethany Gumbrill[89], Rebekah Johnson[89], Katie Sweet[‡,89], Denise Webster[89], Georgia Efford[89].

**Norfolk and Norwich University hospital (NNUH), Norwich, UK** Karen Convery[‡,90], Deirdre Fottrell-Gould[90], Lisa Hudig[90], Jocelyn Keshet-Price[90], Georgina Randell[*,90], Katie Stammers[90].

**Queen Elizabeth Hospital Gateshead, Gateshead, UK** Maria Bokhari[91], Vanessa Linnett[*,91], Rachael Lucas[91], Wendy McCormick[91], Jenny Ritzema[‡,91], Amanda Sanderson[‡,91], Helen Wild[91].

**Sunderland Royal Hospital, Sunderland, UK** Anthony Rostron[*,92], Alistair Roy[*,92], Lindsey Woods[‡,92], Sarah Cornell[92], Fiona Wakinshaw[92], Kimberley Rogerson[92], Jordan Jarmain[92].

**Aintree University Hospital, Liverpool, UK** Robert Parker[*,93], Amie Reddy[93], Ian Turner-Bone[‡,93], Laura Wilding[93], Peter Harding[93].

**Hull Royal Infirmary, Hull, UK** Caroline Abernathy[94], Louise Foster[94], Andrew Gratrix[*,94], Vicky Martinson[94], Priyai Parkinson[94], Elizabeth Stones[94], Llucia Carbral-Ortega[95].

**University College Hospital, London, UK** Georgia Bercades[‡,96], David Brealey[*,96], Ingrid Hass[96], Niall MacCallum[*,96], Gladys Martir[96], Eamon Raith[*,96], Anna Reyes[96], Deborah Smyth[96].

**Royal Devon and Exeter Hospital, Exeter, UK** Letizia Zitter[*,97], Sarah Benyon[97], Suzie Marriott[97], Linda Park[97], Samantha Keenan[97], Elizabeth Gordon[97], Helen Quinn[97], Kizzy Baines[97].

**The Royal Papworth Hospital, Cambridge, UK** Lenka Cagova[98], Adama Fofano[98], Lucie Garner[98], Helen Holcombe[98], Sue Mepham[98], Alice Michael Mitchell[98], Lucy Mwaura[*,98], Krithivasan Praman[98], Alain Vuylsteke[*,98], Julie Zamikula[‡,98].

**Ipswich Hospital, Ipswich, UK** Bally Purewal[*,99].

**Southmead Hospital, Bristol, UK** Hayley Blakemore[100], Borislava Borislavova[100], Beverley Faulkner[‡,100], Emma Gendall[100], Elizabeth Goff[100], Kati Hayes[100], Matt Thomas[*,100], Ruth Worner[100], Kerry Smith[100], Deanna Stephens[100].

**Milton Keynes University Hospital, Milton Keynes, UK** Louise Mew[‡,101], Esther Mwaura[‡,101], Richard Stewart[*,101], Felicity Williams[101], Lynn Wren[‡,101], Sara-Beth Sutherland [‡,101].

**Royal Hampshire County Hospital, Hampshire, UK** Emily Bevan[102], Jane Martin[‡,102], Dawn Trodd[102], Geoff Watson[*,102], Caroline Wrey Brown[102].

**Queen Elizabeth Hospital, Woolwich, London, UK** Amy Collins[‡,103], Waqas Khaliq[*,103], Estefania Treus Gude[103].

**Great Ormond St Hospital and UCL Great Ormond St Institute of Child Health NIHR Biomedical Research Centre, London, UK** Olugbenga Akinkugbe[104], Alasdair Bamford[104], Emily Beech[104], Holly Belfield[104], Michael Bell[104], Charlene Davies[104], Gareth A. L. Jones[104], Tara McHugh[104], Hamza Meghari[104], Lauran O'Neill[‡,104], Mark J. Peters[*,104], Samiran Ray[104], Ana Luisa Tomas[104].

**Stoke Mandeville Hospital, Buckinghamshire, UK** Iona Burn[105], Geraldine Hambrook[‡,105], Katarina Manso[105], Ruth Penn[105], Pradeep Shanmugasundaram[*,105], Julie Tebbutt[105], Danielle Thornton[105].

**University Hospital of Wales, Cardiff, UK** Jade Cole[*,106], Michelle Davies[106], Rhys Davies[106], Donna Duffin[106], Helen Hill[106], Ben Player[106], Emma Thomas[106], Angharad Williams[106].

**Basingstoke and North Hampshire Hospital, Basingstoke, UK** Denise Griffin[107], Nycola Muchenje[107], Mcdonald Mupudzi[107], Richard Partridge[*,107], Jo-Anna Conyngham[107], Rachel Thomas[107], Mary Wright[107], Maria Alvarez Corral[107].

**Arrowe Park Hospital, Wirral, UK** Reni Jacob[108], Cathy Jones[108], Craig Denmade[*,108].

**Chesterfield Royal Hospital Foundation Trust, Chesterfield, UK** Sarah Beavis[*,109], Katie Dale[109], Rachel Gascoyne[109], Joanne Hawes[109], Kelly Pritchard[109], Lesley Stevenson[109], Amanda Whileman[‡,109].

**Musgrove Park Hospital, Taunton, UK** Patricia Doble[‡,110], Joanne Hutter[110], corinne Pawley[110], Charmaine Shovelton[110], Marius Vaida[*,110].

**Peterborough City Hospital, Peterborough, UK and Hinchingbrooke Hospital, Huntingdon, UK** Deborah Butcher[111], Susie O'Sullivan[‡,111], Nicola Butterworth-Cowin[*,111].

**Royal Hallamshire Hospital and Northern General Hospital, Sheffield, UK** Norfaizan Ahmad[112], Joann Barker[112], Kris Bauchmuller[112], Sarah Bird[112], Kay Cawthron[112], Kate Harrington[112], Yvonne Jackson[112], Faith Kibutu[112], Becky Lenagh[112], Shamiso Masuko[112], Gary H Mills[*,112], Ajay Raithatha[112], Matthew Wiles[112], Jayne Willson[‡,112], Helen Newell[112], Alison Lye[112], Lorenza Nwafor[112], Claire Jarman[112], Sarah Rowland-Jones[112], David Foote[112], Joby Cole[112], Roger Thompson[112], James Watson[112], Lisa Hesseldon[112], Irene Macharia[112], Luke Chetam [112], Jacqui Smith[112], Amber Ford[112], Samantha Anderson[112], Kathryn Birchall[112], Kay Housley[112], Sara Walker[112], Leanne Milner[112], Helena Hanratty[112], Helen Trower[112], Patrick Phillips[112], Simon Oxspring[112], Ben Donne[112].

**Dumfries and Galloway Royal Infirmary, Dumfries, UK** Catherine Jardine[‡,113], Dewi Williams[*,113], Alasdair Hay[*,113].

**Royal Bolton Hospital, Bolton, UK** Rebecca Flanagan[114], Gareth Hughes[*,114], scott Latham[‡,114], Emma McKenna[‡,114], Jennifer Anderson[‡,114], Robert Hull[114], Kat Rhead[114].

**Lister Hospital, Stevenage, UK** Carina Cruz[‡,115], Natalie Pattison[*,115].

**Craigavon Area Hospital, County Armagh, NI** Rob Charnock[*,116], Denise McFarland[116], Denise Cosgrove[116].

**Southport and Formby District General Hospital, Ormskirk, UK** Ashar Ahmed[*,117], Anna Morris[117], Srinivas Jakkula[*,117].

**Calderdale Royal Hospital, Halifax, UK and Huddersfield Royal Infirmary, Huddersfield, UK** Asifa Ali[118].

**Calderdale Royal Hospital, Halifax, UK** Megan Brady[118], Sam Dale[118], Annalisa Dance[118], Lisa Gledhill[118], Jill Greig[118], Kathryn Hanson[118], Kelly Holdroyd[118], Marie Home[118], Diane Kelly[118], Ross Kitson[*,118], Lear Matapure[118], Deborah Melia[118], Samantha Mellor[118], Tonicha Nortcliffe[118], Jez Pinnell[*,118], Matthew Robinson[118], Lisa Shaw[118], Ryan Shaw[118], Lesley Thomis[118], Alison Wilson[‡,118], Tracy Wood[118], Lee-Ann Bayo[118], Ekta Merwaha[118], Tahira Ishaq[118], Sarah Hanley[118].

**Prince Charles Hospital, Merthyr Tydfil, UK** Bethan Deacon[119], Meg Hibbert[119], Carla Pothecary[119], Dariusz Tetla[*,119], Chrsitopher Woodford[119], Latha Durga[119], Gareth Kennard-Holden [119].

**Royal Bournemouth Hospital, Bournemouth, UK** Debbie Branney[‡,120], Jordan Frankham[120], Sally Pitts[‡,120], Nigel White[120].

**Royal Preston Hospital, Preston, UK** Shondipon Laha[121], Mark Verlander[121], Alexandra Williams[*,121].

**Whittington Hospital, London, UK** Abdelhakim Altabaibeh[*,122], Ana Alvaro[122], Kayleigh Gilbert[122], Louise Ma[122], Loreta Mostoles[‡,122], Chetan Parmar[*,122], Kathryn Simpson[122], Champa Jetha[122], Lauren Booker[122], Anezka Pratley[122].

**Princess Royal Hospital, Telford and Royal Shrewsbury Hospital, Shrewsbury, UK** Colene Adams[123], Anita Agasou[123], Tracie Arden[123], Amy Bowes[123], Pauline Boyle[123], Mandy Beekes[123], Heather Button[123], Nigel Capps[*,123], Mandy Carnahan[123], Anne Carter[123], Danielle Childs[123], Denise Donaldson[‡,123], Kelly Hard[123], Fran Hurford[123], Yasmin Hussain[123], Ayesha Javaid[123], James Jones[123], Sanal Jose[123], Michael Leigh[123], Terry Martin[123], Helen Millward[123], Nichola Motherwell[123], Rachel Rikunenko[123], Jo Stickley[123], Julie Summers[123], Louise Ting[123], Helen Tivenan[123], Louise Tonks[123], Rebecca Wilcox[123].

**Macclesfield District General Hospital, Macclesfield, UK** Maureen Holland[124], Natalie Keenan[124], Marc Lyons[*,124], Helen Wassall[124], Chris Marsh[124], Mervin Mahenthran[124], Emma Carter[124], Thomas Kong[124].

**Royal Surrey County Hospital, Guildford, UK** Helen Blackman[‡,125], Ben Creagh-Brown[*,125], Sinead Donlon[125], Natalia Michalak-Glinska[125], Sheila Mtuwa[125], Veronika Pristopan[125], Armorel Salberg[‡,125], Eleanor Smith[125], Sarah Stone[125], Charles Piercy[125], Jerik Verula[125], Dorota Burda[125], Rugia Montaser[125], Lesley Harden[125], Irving Mayangao[125], Cheryl Marriott[‡,125], Paul Bradley[125], Celia Harris[125].

**Hereford County Hospital, Hereford, UK** Susan Anderson[‡,126], Eleanor Andrews[126], Janine Birch[‡,126], Emma Collins[‡,126], Kate Hammerton[126], Ryan O'Leary[*,126].

**University Hospital of North Tees, Stockton on Tees, UK** Michele Clark[*,127], Sarah Purvis[*,127].

**Lincoln County Hospital, Lincoln, UK** Russell Barber[*,128], Claire Hewitt[128], Annette Hilldrith[128], Karen Jackson-Lawrence[128], Sarah Shepardson[128], Maryanne Wills[128], Susan Butler [128], Silvia Tavares[‡,128], Amy Cunningham [128], Julia Hindale [128], Sarwat Arif [128].

**Royal Cornwall Hospital, Truro, UK** Sarah Bean[129], Karen Burt[129], Michael Spivey[*,129].

**Royal United Hospital, Bath, UK** Carrie Demetriou[130], Charlotte Eckbad[130], Sarah Hierons[‡,130], Lucy Howie[130], Sarah Mitchard[130], Lidia Ramos[130], Alfredo Serrano-Ruiz[*,130], Katie White[130], Fiona Kelly[*,130].

**Royal Brompton Hospital, London, UK** Daniele Cristiano[131], Natalie Dormand[131], Zohreh Farzad[131], Mahitha Gummadi[131], Kamal Liyanage[131], Brijesh Patel[*,131], Sara Salmi[131], Geraldine Sloane[131], Vicky Thwaites[‡,131], Mathew Varghese[131], Anelise C Zborowski[131].

**University Hospital Crosshouse, Kilmarnock, UK** John Allan[132], Tim Geary[132], Gordon Houston[*,132], Alistair Meikle[132], Peter O'Brien[132].

**Basildon Hospital, Basildon, UK** Miranda Forsey[133], Agilan Kaliappan[*,133], Anne Nicholson[133], Joanne Riches[133], Mark Vertue[‡,133], Miranda Forsey[133], Agilan Kaliappan[*,133], Anne Nicholson[133], Joanne Riches[133], Mark Vertue[‡,133].

**Glan Clwyd Hospital, Bodelwyddan, UK** Elizabeth Allan[134], Kate Darlington[134], Ffyon Davies[134], Jack Easton[134], Sumit Kumar[134], Richard Lean[134], Daniel Menzies[*,134], Richard Pugh[*,134], Xinyi Qiu[134], Llinos Davies[134], Hannah Williams [134], Jeremy Scanlon[134], Gwyneth Davies [134], Callum Mackay[134], Joannne Lewis[134], Stephanie Rees[134].

**West Middlesex Hospital, Isleworth, UK** Metod Oblak[135], Monica Popescu[*,135], Mini Thankachen[135].

**Royal Lancaster Infirmary, Lancaster, UK** Andrew Higham[*,136], Kerry Simpson[‡,136], Jayne Craig[136].

**Western General Hospital, Edinburgh, UK** Rosie Baruah[*,137], Sheila Morris[‡,137], Susie Ferguson[137], Amy Shepherd[137].

**Chelsea & Westminster NHS Foundation Trust, London, UK** Luke Stephen Prockter Moore[138], Marcela Paola Vizcaychipi[138], Laura Gomes de Almeida Martins[138], Jaime Carungcong[138].

**The Queen Elizabeth Hospital, King's Lynn, UK** Inthakab Ali Mohamed Ali[139], Karen Beaumont[139], Mark Blunt[*,139], Zoe Coton[‡,139], Hollie Curgenven[139], Mohamed Elsaadany[139], Kay Fernandes[139], Sameena Mohamed Ally[139], Harini Rangarajan[139], Varun Sarathy[139], Sivarupan Selvanayagam[139], Dave Vedage[139], Matthew White[139].

**King's Mill Hospital, Nottingham, UK** Mandy Gill[‡,140], Paul Paul[*,140], Valli Ratnam[*,140], Sarah Shelton[140], Inez Wynter[140].

**Watford General Hospital, Watford, UK** Siobhain Carmody[‡,141], Valerie Joan Page[*,141].

**University Hospital Wishaw, Wishaw, UK** Claire Marie Beith[142], Karen Black[142], Suzanne Clements[142], Alan Morrison[142], Dominic Strachan[*,142], Margaret Taylor[‡,142], Michelle Clarkson[142], Stuart D'Sylva[142], Kathryn Norman[142].

**Forth Valley Royal Hospital, Falkirk, UK** Fiona Auld[143], Joanne Donnachie[143], Ian Edmond[*,143], Lynn Prentice[143], Nikole Runciman[143], Dario Salutous[‡,143], Lesley Symon[143], Anne Todd[143], Patricia Turner[143], Abigail Short[143], Laura Sweeney[143], Euan Murdoch[143], Dhaneesha Senaratne[143].

**George Eliot Hospital NHS Trust, Nuneaton, UK** Michaela Hill[144], Thogulava Kannan[*,144], Wild Laura[144].

**Barnsley Hospital, Barnsley, UK** Rikki Crawley[145], Abigail Crew[145], Mishell Cunningham[145], Allison Daniels[145], Laura Harrison[145], Susan Hope[145], Ken Inweregbu[*,145], Sian Jones[145], Nicola Lancaster[145], Jamie Matthews[145], Alice Nicholson[‡,145], Gemma Wray[145].

**The Great Western Hospital, Swindon, UK** Helen Langton[146], Rachel Prout[*,146], Malcolm Watters[*,146], Catherine Novis[146].

**Harefield Hospital, London, UK** Anthony Barron[147], Ciara Collins[147], Sundeep Kaul[147], Heather Passmore[147], Claire Prendergast[147], Anna Reed[*,147], Paula Rogers[147], Rajvinder Shokkar[147], Meriel Woodruff[147], Hayley Middleton [147], Oliver Polgar[147], Claire Nolan [147], Vicky Thwaites[‡,147], Kanta Mahay[‡,147].

**Rotherham General Hospital, Rotherham, UK** Dawn Collier[148], Anil Hormis[*,148], Rachel Walker[‡,148], Victoria Maynard[148].

**Ysbyty Gwynedd, Bangor, UK** Ellen Knights[149], Alicia Price[149], Alice Thomas[‡,149], Chris Thorpe[*,149].

**Diana Princess of Wales Hospital, Grimsby, UK** Teresa Behan[150], Caroline Burnett[150], Jonathan Hatton[150], Elaine Heeney[150], Atideb Mitra[*,150], Maria Newton[150], Rachel Pollard[150], Rachael Stead[‡,150].

**Russell's Hall Hospital, Dudley, UK** Vishal Amin[*,151], Elena Anastasescu[‡,151], Vikram Anumakonda[151], Komala Karthik[151], Rizwana Kausar[151], Karen Reid[‡,151], Jacqueline Smith[151], Janet Imeson-Wood[151].

**Princess Royal Hospital** Denise Skinner[152], Jane Gaylard[152], Dee Mullan[152], Julie Newman[152].

**Princess Royal Hospital, Haywards Heath, UK** Denise Skinner[152], Jane Gaylard[152], Dee Mullan[152], Julie Newman[152].

**St Mary's Hospital, Newport, UK** Alison Brown[153], Vikki Crickmore[153], Gabor Debreceni[*,153], Joy Wilkins[153], Liz Nicol[153].

**University Hospital Lewisham, London, UK** Waqas Khaliq[*,154], Rosie Reece-Anthony[‡,154], Mark Birt[154].

**Colchester General Hospital, Colchester, UK** Alison Ghosh[*,155], Emma Williams[155].

**Queen Elizabeth the Queen Mother Hospital, Margate, UK** Louise Allen[156], Eva Beranova[156], Nikki Crisp[156], Joanne Deery[156], Tracy Hazelton[156], Alicia Knight[156], Carly Price[156], Sorrell Tilbey[156], Salah Turki[*,156], Sharon Turney[156].

**Royal Albert Edward Infirmary, Wigan, UK** Joshua Cooper[157], Cheryl Finch[‡,157], Sarah Liderth[157], Alison Quinn[*,157], Natalia Waddington[157].

**Victoria Hospital, Kirkcaldy, UK** Tina Coventry[158], Susan Fowler[‡,158], Michael MacMahon[*,158], Amanda McGregor[158].

**Eastbourne District General Hospital, East Sussex, UK and Conquest Hospital, East Sussex, UK** Anne Cowley[‡,159], Judith Highgate[*,159], Anne Cowley[‡,159], Judith Highgate[*,159].

**Cumberland Infirmary, Carlisle, UK** Alison Brown[160], Jane Gregory[‡,160], Susan O'Connell[160], Tim Smith[*,160], Luigi Barberis[160].

**New Cross Hospital, Wolverhampton, UK** Shameer Gopal[*,161], Nichola Harris[‡,161], Victoria Lake[‡,161], Stella Metherell[161], Elizabeth Radford[161].

**The Princess Alexandra Hospital, Harlow, UK** Amelia Daniel[162], Joanne Finn[162], Rajnish Saha[*,162], Nikki White[‡,162], Amy Easthope[162].

**Salisbury District Hospital, Salisbury, UK** Phil Donnison[*,163], Fiona Trim[163], Beena Eapen[163].

**Dorset County Hospital, Dorchester, UK** Jenny Birch[164], Laura Bough[164], Josie Goodsell[164], Rebecca Tutton[164], Patricia Williams[‡,164], Sarah Williams[*,164], Barbara Winter-Goodwin[164].

**University College Dublin, St Vincent's University Hospital, Dublin, Ireland** Ailstair Nichol[*,165], Kathy Brickell[‡,165], Michelle Smyth [165], Lorna Murphy[165].

**Glangwili General Hospital, Camarthen, UK** Samantha Coetzee[‡,166], Alistair Gales[*,166], Igor Otahal[*,166], Meena Raj[166], Craig Sell[*,166].

**Gloucestershire Royal Hospital, Gloucester, UK** Paula Hilltout[*,167], Jayne Evitts[167], Amanda Tyler[167], Joanne Waldron[167].

**Yeovil Hospital, Yeovil, UK** Kate Beesley[168], Sarah Board[168], Agnieszka Kubisz-Pudelko[*,168], Alison Lewis[168], Jess Perry[168], Lucy Pippard[168], Di Wood[‡,168], Clare Buckley[168].

**Leicester Royal Infirmary, Leicester, UK** Peter Barry[*,169], Neil Flint[*,169], Patel Rekha [‡,169], Dawn Hales[‡,169].

**Royal Manchester Children's Hospital, Manchester, UK** Lara Bunni[170], Claire Jennings[‡,170], Monica Latif[170], Rebecca Marshall[‡,170], Gayathri Subramanian[*,170].

**Royal Victoria Hospital, Belfast, NI** Peter J McGuigan[*,171], Christopher Wasson[171], Stephanie Finn[171], Jackie Green[171], Erin Collins[171], Bernadette King[171].

**Wrexham Maelor Hospital, Wrexham, Wales** Andy Campbell[*,172], Sara Smuts[172], Joseph Duffield[172], Oliver Smith[172], Lewis Mallon[172], Watkins Claire[172].

**Walsall Manor Hospital, Walsall, UK** Liam Botfield[173], Joanna Butler[‡,173], Catherine Dexter[173], Jo Fletcher[173], Atul Garg[*,173], Aditya Kuravi[*,173], Poonam Ranga[‡,173], Emma Virgilio[‡,173].

**Darent Valley Hospital, Dartford, UK** Zakaula Belagodu[*,174], Bridget Fuller[174], Anca Gherman[174], Olumide Olufuwa[‡,174], Remi Paramsothy[174], Carmel Stuart[174], Naomi Oakley[174], Charlotte Kamundi[174], David Tyl[174], Katy Collins[174], Pedro Silva[174], June Taylor[174], Laura King[174], Charlotte Coates[174], Maria Crowley[174], Phillipa Wakefield[174], Jane Beadle [174], Laura Johnson[174], Janet Sargeant[174], Madeleine Anderson[174].

**Warrington General Hospital, Warrington, UK** Ailbhe Brady[175], Rebekah Chan[175], Jeff Little[*,175], Shane McIvor[175], Helena Prady[175], Helen Whittle[175], Bijoy Mathew[175].

**Warwick Hospital, Warwick, UK** Ben Attwood[*,176], Penny Parsons[‡,176].

**University Hospitals Coventry & Warwickshire NHS Trust, Coventry, UK** Geraldine Ward[177], Pamela Bremmer[177].

**University Hospital Monklands, Airdrie, UK** West Joe[178], Baird Tracy[178], Ruddy Jim[178].

**Princess of Wales Hospital, Llantrisant, UK** Ellie Davies[179], Lisa Roche[179], Sonia Sathe[*,179].

**Northwick Park Hospital, London, UK** Catherine Dennis[180], Alastair McGregor[*,180], Victoria Parris[*,180], Sinduya Srikaran[180], Anisha Sukha[180].

**Raigmore Hospital, Inverness, UK** Rachael Campbell[‡,181], Noreen Clarke[181], Jonathan Whiteside[*,181], Mairi Mascarenhas[181], Avril Donaldson[181], Joanna Matheson[181], Fiona Barrett[181], Marianne O'Hara[181], Laura Okeefe[181], Clare Bradley[181].

**Royal Free Hospital, London, UK** Christine Eastgate-Jackson[182], Helder Filipe[‡,182], Daniel Martin[*,182], Amitaa Maharajh[182], Sara Mingo Garcia[182], Mark De Neef[182].

**Scunthorpe General Hospital, Scunthorpe, UK** Kathy Dent[‡,183], Elizabeth Horsley[183], Muhmmad Nauman Akhtar[*,183], Sandra Pearson[183], Dorota Potoczna[‡,183], Sue Spencer[183].

**West Cumberland Hospital, Whitehaven, UK** Melanie Clapham[‡,184], Rosemary Harper[184], Una Poultney[184], Polly Rice[184], Tim Smith[184], Rachel Mutch[184], Luigi Barberis[*,184].

**Airedale General Hospital, Keighley, UK** Lisa Armstrong[185], Hayley Bates[‡,185], Emma Dooks[‡,185], Fiona Farquhar[‡,185], Brigid Hairsine[*,185], Chantal McParland[185], Sophie Packham[‡,185].

**Birmingham Children's Hospital, Birmingham, UK** Rehana Bi[‡,186], Barney Scholefield[*,186], Lydia Ashton[‡,186].

**Liverpool Heart and Chest Hospital, Liverpool, UK** Linsha George[‡,187], Sophie Twiss[187], David Wright[*,187].

**Pilgrim Hospital, Lincoln, UK** Manish Chablani[*,188], Amy Kirkby[188], Kimberley Netherton[188].

**Prince Philip Hospital, Lianelli, UK** Kim Davies[189], Linda O'Brien[‡,189], Zohra Omar[189], Igor Otahal[*,189], Emma Perkins[189], Tracy Lewis[189], Isobel Sutherland[189].

**Furness General Hospital, Barrow-in-Furness, UK** Karen Burns[‡,190], Andrew Higham[*,190].

**Scarborough General Hospital, Scarborough, UK** Dr Ben Chandler[191], Kerry Elliott[*,191], Janine Mallinson[*,191], Alison Turnbull[191].

**Southend University Hospital, Westcliff-on-Sea, UK** Prisca Gondo[*,192], Bernard Hadebe[192], Abdul Kayani[192], Bridgett Masunda[‡,192].

**Alder Hey Children's Hospital, Liverpool, UK** Taya Anderson[193], Dan Hawcutt[*,193], Laura O'Malley[‡,193], Laura Rad[193], Naomi Rogers[193], Paula Saunderson[193], Kathryn Sian Allison[193], Deborah Afolabi[193], jennifer whitbread[193], Dawn jones [193], Rachael Dore [193].

**Torbay Hospital, Torquay, UK** Matthew Halkes[*,194], Pauline Mercer[‡,194], Lorraine Thornton[194].

**Borders General Hospital, Melrose, UK** Joy Dawson[‡,195], Sweyn Garrioch[*,195], Melanie Tolson[195], Jonathan Aldridge[195].

**Kent & Canterbury Hospital, Canterbury, UK** Ritoo Kapoor[*,196], David Loader[196], Karen Castle[196].

**West Suffolk Hospital, Bury St Edmunds, UK** Sally humphreys[*,197], Ruth Tampsett[197].

**James Paget University Hospital NHS Trust, Great Yarmouth, UK** Katherine Mackintosh[*,198], Amanda Ayers[198], Wendy Harrison[‡,198], Julie North[198].

**The Christie NHS Foundation Trust, Manchester, UK** Suzanne Allibone[‡,199], Roman Genetu[199], Vidya Kasipandian[199], Amit Patel[*,199], Ainhi Mac[199], Anthony Murphy[199], Parisa Mahjoob[199], Roonak Nazari[199], Lucy Worsley[199], Andrew Fagan[199].

**The Royal Marsden Hospital, London, UK** Thomas Bemand[200], Ethel Black[‡,200], Arnold Dela Rosa[200], Ryan Howle[200], Shaman Jhanji[*,200], Ravishankar Rao Baikady[200], Kate Colette Tatham[*,200], Benjamin Thomas[200].

**University Hospital Hairmyres, East Kilbride, UK** Dina Bell[201], Rosalind Boyle[201], Katie Douglas[201], Lynn Glass[201], Emma Lee[201], Liz Lennon[201], Austin Rattray[*,201].

**Withybush General Hospital, Pembrokeshire, Wales** Abigail Taylor[202], Rachel Anne Hughes[202], Helen Thomas[202], Alun Rees[202], Michaela Duskova[202], Janet Phipps[202], Suzanne Brooks[202], Michelle Edwards[202].

**Ealing Hospital, Southall, UK** Victoria Parris[*,203], Sheena Quaid[‡,203], Ekaterina Watson[203].

**North Devon District Hospital, Barnstaple, UK** Adam Brayne[204], Emma Fisher[204], Jane Hunt[‡,204], Peter Jackson[204], Duncan Kaye[204], Nicholas Love[*,204], Juliet Parkin[204], Victoria Tuckey[204], Lynne Van Koutrik[204], Sasha Carter[204], Benedict Andrew[204], Louise Findlay[204], Katie Adams[204].

**St John's Hospital Livingston, Livingston, UK** Alison Williams[205], Claire Cheyne[205], Anne Saunderson[205], Sam Moultrie[*,205], Miranda Odam[205].

**Northampton General Hospital NHS Trust, Northampton, UK** Kathryn Hall[206], Ishe-unesu Mapfunde [206].

**Harrogate and District NHS Foundation Trust, Harrogate, UK** Chunda Sri-Chandana[207], Joslan Scherewode[207], Lorraine Stephenson [207], Sarah Marsh [207].

**National Hospital for Neurology and Neurosurgery, London, UK** David Brealey[*,208], John Hardy[208], Henry Houlden[208], Eleanor Moncur[208], Eamon Raith[*,208], Ambreen Tariq[‡,208], Arianna Tucci[208].

**Bronglais General Hospital, Aberystwyth, UK** Maria Hobrok[*,209], Ronda Loosley[209], Heather McGuinness[209], Helen Tench[‡,209], Rebecca Wolf-Roberts[209].

**Golden Jubilee National Hospital, Clydebank, UK** Val Irvine[‡,210], Benjamin Shelley[*,210].

**Homerton University Hospital Foundation NHS Trust, London UK** Amy Easthope[211], Claire Gorman[211], Abhinav Gupta[*,211], Elizabeth Timlick[‡,211], Rebecca Brady[211].

**Royal Hospital for Children, Glasgow, UK** Colin Begg[*,3], Barry Milligan[3].

**Sheffield Children's Hospital, Sheffield, UK** Arianna Bellini[‡,212], Jade Bryant[212], Anton Mayer[*,212], Amy Pickard[212], Nicholas Roe[212], Jason Sowter[212], Alex Howlett [212].

‡ - Team Lead

* - PI

[1]Roslin Institute, University of Edinburgh, Easter Bush, Edinburgh, EH25 9RG, UK
[2]Intensive Care Unit, Royal Infirmary of Edinburgh, 54 Little France Drive, Edinburgh, EH16 5SA, UK
[3]Royal Hospital for Children, Glasgow, UK
[4]William Harvey Research Institute, Barts and the London School of Medicine and Dentistry, Queen Mary University of London, London EC1M 6BQ, UK
[5]Centre for Tropical Medicine and Global Health, Nuffield Department of Medicine, University of Oxford, Old Road Campus, Roosevelt Drive, Oxford, OX3 7FZ, UK
[6]Wellcome Centre for Human Genetics, University of Oxford, Oxford, UK
[7]Department of Critical Care Medicine, Queen's University and Kingston Health Sciences Centre, Kingston, ON, Canada
[8]Wellcome-Wolfson Institute for Experimental Medicine, Queen's University Belfast, Belfast, Northern Ireland, UK
[9]Department of Intensive Care Medicine, Royal Victoria Hospital, Belfast, Northern Ireland, UK
[10]UCL Centre for Human Health and Performance, London, W1T 7HA, UK
[11]Clinical Research Centre at St Vincent's University Hospital, University College Dublin, Dublin,

Ireland
[12]National Heart and Lung Institute, Imperial College London, London, UK
[13]Imperial College Healthcare NHS Trust:London,London,UK
[14]MRC Human Genetics Unit, Institute of Genetics and Molecular Medicine, University of Edinburgh, Western General Hospital, Crewe Road, Edinburgh, EH4 2XU, UK
[15]Intensive Care National Audit & Research Centre, London, UK
[16]NIHR Health Protection Research Unit for Emerging and Zoonotic Infections, Institute of Infection, Veterinary and Ecological Sciences University of Liverpool, Liverpool, L69 7BE, UK
[17]Respiratory Medicine, Alder Hey Children's Hospital, Institute in The Park, University of Liverpool, Alder Hey Children's Hospital, Liverpool, UK
[18]Department of Intensive Care Medicine, Guy's and St. Thomas NHS Foundation Trust, London, UK
[19]Department of Medicine, University of Cambridge, Cambridge, UK
[20]Cambridge University Hospitals NHS Foundation Trust, Hills Road, Cambridge, CB2 0QQ, UK
[21]Edinburgh Clinical Research Facility, Western General Hospital, University of Edinburgh, EH4 2XU, UK
[22]Department of Infectious Diseases, Leiden University Medical Center, Leiden, The Netherlands
[23]Guys and St Thomas' Hospital, London, UK
[24]Barts Health NHS Trust, London, UK
[25]James Cook University Hospital, Middlesbrough, UK
[26]Royal Stoke University Hospital, Staffordshire, UK
[27]North Middlesex University Hospital NHS trust, London, UK
[28]north Middlesex University Hospital NHS trust, London, UK
[29]The Royal Liverpool University Hospital, Liverpool, UK
[30]King's College Hospital, London, UK
[31]Charing Cross Hospital, St Mary's Hospital and Hammersmith Hospital, London, UK
[32]Nottingham University Hospital, Nottingham, UK
[33]John Radcliffe Hospital, Oxford, UK
[34]Kingston Hospital, Surrey, UK
[35]Royal Infirmary of Edinburgh, Edinburgh, UK
[36]Queen Alexandra Hospital, Portsmouth, UK
[37]Morriston Hospital, Swansea, UK
[38]Addenbrooke's Hospital, Cambridge, UK
[39]BHRUT (Barking Havering) - Queens Hospital and King George Hospital, Essex, UK
[40]Royal Sussex County Hospital, Brighton, UK
[41]Queen Elizabeth Hospital, Birmingham, UK
[42]St George's Hospital, London, UK
[43]Stepping Hill Hospital, Stockport, UK
[44]Countess of Chester Hospital, Chester, UK
[45]Royal Blackburn Teaching Hospital, Blackburn, UK
[46]The Tunbridge Wells Hospital and Maidstone Hospital, Kent, UK
[47]Royal Gwent Hospital, Newport, UK
[48]Pinderfields General Hospital, Wakefield, UK
[49]Royal Berkshire NHS Foundation Trust, Berkshire, UK
[50]Broomfield Hospital, Chelmsford, UK
[51]Northumbria Healthcare NHS Foundation Trust, North Shields, UK

[52]Whiston Hospital, Prescot, UK
[53]Croydon University Hospital, Croydon, UK
[54]York Hospital, York, UK
[55]Heartlands Hospital, Birmingham, UK
[56]Ashford and St Peter's Hospital, Surrey, UK
[57]Barnet Hospital, London, UK
[58]East Surrey Hospital, Redhill, UK
[59]Ninewells Hospital, Dundee, UK
[60]Worthing Hospital, Worthing, UK and St Richard's Hospital, Chichester, UK
[61]Southampton General Hospital, Southampton, UK
[62]The Alexandra Hospital, Redditch and Worcester Royal Hospital, Worcester, UK
[63]Sandwell General Hospital and City Hospital, Birmingham, UK
[64]Blackpool Victoria Hospital, Blackpool, UK
[65]Royal Glamorgan Hospital, Pontyclun, UK
[66]The Royal Oldham Hospital, Manchester, UK
[67]Glasgow Royal Infirmary, Glasgow, UK
[68]St James's University Hospital and Leeds General Infirmary, Leeds, UK
[69]University Hospital North Durham, Darlington, UK and Darlington Memorial Hospital, Darlington, UK
[70]Fairfield General Hospital, Bury, UK
[71]Wythenshawe Hospital, Manchester, UK
[72]Royal Alexandra Hospital, Paisley, UK
[73]Good Hope Hospital, Birmingham, UK
[74]Tameside General Hospital, Ashton Under Lyne, UK
[75]Royal Derby Hospital, Derby, UK
[76]Medway Maritime Hospital, Gillingham, UK
[77]Royal Victoria Infirmary, Newcastle Upon Tyne, UK
[78]Poole Hospital, Poole, UK
[79]Bedford Hospital, Bedford, UK
[80]Queens Hospital Burton, Burton-On-Trent, UK
[81]North Manchester General Hospital, Manchester, UK
[82]Aberdeen Royal Infirmary, Aberdeen, UK
[83]Derriford Hospital, Plymouth, UK
[84]Manchester Royal Infirmary, Manchester, UK
[85]Salford Royal Hospital, Manchester, UK
[86]William Harvey Hospital, Ashford, UK
[87]Queen Elizabeth University Hospital, Glasgow, UK
[88]Bradford Royal Infirmary, Bradford, UK
[89]Bristol Royal Infirmary, Bristol, UK
[90]Norfolk and Norwich University hospital (NNUH), Norwich, UK
[91]Queen Elizabeth Hospital Gateshead, Gateshead, UK
[92]Sunderland Royal Hospital, Sunderland, UK
[93]Aintree University Hospital, Liverpool, UK
[94]Hull Royal Infirmary, Hull, UK
[95]Hull Royal Infirmary, Hull, Uk
[96]University College Hospital, London, UK

[97]Royal Devon and Exeter Hospital, Exeter, UK
[98]The Royal Papworth Hospital, Cambridge, UK
[99]Ipswich Hospital, Ipswich, UK
[100]Southmead Hospital, Bristol, UK
[101]Milton Keynes University Hospital, Milton Keynes, UK
[102]Royal Hampshire County Hospital, Hampshire, UK
[103]Queen Elizabeth Hospital, Woolwich, London, UK
[104]Great Ormond St Hospital and UCL Great Ormond St Institute of Child Health NIHR Biomedical Research Centre, London, UK
[105]Stoke Mandeville Hospital, Buckinghamshire, UK
[106]University Hospital of Wales, Cardiff, UK
[107]Basingstoke and North Hampshire Hospital, Basingstoke, UK
[108]Arrowe Park Hospital, Wirral, UK
[109]Chesterfield Royal Hospital Foundation Trust, Chesterfield, UK
[110]Musgrove Park Hospital, Taunton, UK
[111]Peterborough City Hospital, Peterborough, UK and Hinchingbrooke Hospital, Huntingdon, UK
[112]Royal Hallamshire Hospital and Northern General Hospital, Sheffield, UK
[113]Dumfries and Galloway Royal Infirmary, Dumfries, UK
[114]Royal Bolton Hospital, Bolton, UK
[115]Lister Hospital, Stevenage, UK
[116]Craigavon Area Hospital, County Armagh, NI
[117]Southport and Formby District General Hospital, Ormskirk, UK
[118]Calderdale Royal Hospital, Halifax, UK and Huddersfield Royal Infirmary, Huddersfield, UK
[119]Prince Charles Hospital, Merthyr Tydfil, UK
[120]Royal Bournemouth Hospital, Bournemouth, UK
[121]Royal Preston Hospital, Preston, UK
[122]Whittington Hospital, London, UK
[123]Princess Royal Hospital, Telford and Royal Shrewsbury Hospital, Shrewsbury, UK
[124]Macclesfield District General Hospital, Macclesfield, UK
[125]Royal Surrey County Hospital, Guildford, UK
[126]Hereford County Hospital, Hereford, UK
[127]University Hospital of North Tees, Stockton on Tees, UK
[128]Lincoln County Hospital, Lincoln, UK
[129]Royal Cornwall Hospital, Truro, UK
[130]Royal United Hospital, Bath, UK
[131]Royal Brompton Hospital, London, UK
[132]University Hospital Crosshouse, Kilmarnock, UK
[133]Basildon Hospital, Basildon, UK
[134]Glan Clwyd Hospital, Bodelwyddan, UK
[135]West Middlesex Hospital, Isleworth, UK
[136]Royal Lancaster Infirmary, Lancaster, UK
[137]Western General Hospital, Edinburgh, UK
[138]Chelsea & Westminster NHS Foundation Trust, London, UK
[139]The Queen Elizabeth Hospital, King's Lynn, UK
[140]King's Mill Hospital, Nottingham, UK
[141]Watford General Hospital, Watford, UK

[142]University Hospital Wishaw, Wishaw, UK
[143]Forth Valley Royal Hospital, Falkirk, UK
[144]George Eliot Hospital NHS Trust, Nuneaton, UK
[145]Barnsley Hospital, Barnsley, UK
[146]The Great Western Hospital, Swindon, UK
[147]Harefield Hospital, London, UK
[148]Rotherham General Hospital, Rotherham, UK
[149]Ysbyty Gwynedd, Bangor, UK
[150]Diana Princess of Wales Hospital, Grimsby, UK
[151]Russell's Hall Hospital, Dudley, UK
[152]Princess Royal Hospital, Haywards Heath, UK
[153]St Mary's Hospital, Newport, UK
[154]University Hospital Lewisham, London, UK
[155]Colchester General Hospital, Colchester, UK
[156]Queen Elizabeth the Queen Mother Hospital, Margate, UK
[157]Royal Albert Edward Infirmary, Wigan, UK
[158]Victoria Hospital, Kirkcaldy, UK
[159]Eastbourne District General Hospital, East Sussex, UK and Conquest Hospital, East Sussex, UK
[160]Cumberland Infirmary, Carlisle, UK
[161]New Cross Hospital, Wolverhampton, UK
[162]The Princess Alexandra Hospital, Harlow, UK
[163]Salisbury District Hospital, Salisbury, UK
[164]Dorset County Hospital, Dorchester, UK
[165]University College Dublin, St Vincent's University Hospital, Dublin, Ireland
[166]Glangwili General Hospital, Camarthen, UK
[167]Gloucestershire Royal Hospital, Gloucester, UK
[168]Yeovil Hospital, Yeovil, UK
[169]Leicester Royal Infirmary, Leicester, UK
[170]Royal Manchester Children's Hospital, Manchester, UK
[171]Royal Victoria Hospital, Belfast, NI
[172]Wrexham Maelor Hospital, Wrexham, Wales
[173]Walsall Manor Hospital, Walsall, UK
[174]Darent Valley Hospital, Dartford, UK
[175]Warrington General Hospital, Warrington, UK
[176]Warwick Hospital, Warwick, UK
[177]University Hospitals Coventry & Warwickshire NHS Trust, Coventry, UK
[178]University Hospital Monklands, Airdrie, UK
[179]Princess of Wales Hospital, Llantrisant, UK
[180]Northwick Park Hospital, London, UK
[181]Raigmore Hospital, Inverness, UK
[182]Royal Free Hospital, London, UK
[183]Scunthorpe General Hospital, Scunthorpe, UK
[184]West Cumberland Hospital, Whitehaven, UK
[185]Airedale General Hospital, Keighley, UK
[186]Birmingham Children's Hospital, Birmingham, UK
[187]Liverpool Heart and Chest Hospital, Liverpool, UK

[188]Pilgrim Hospital, Lincoln, UK
[189]Prince Philip Hospital, Lianelli, UK
[190]Furness General Hospital, Barrow-in-Furness, UK
[191]Scarborough General Hospital, Scarborough, UK
[192]Southend University Hospital, Westcliff-on-Sea, UK
[193]Alder Hey Children's Hospital, Liverpool, UK
[194]Torbay Hospital, Torquay, UK
[195]Borders General Hospital, Melrose, UK
[196]Kent & Canterbury Hospital, Canterbury, UK
[197]West Suffolk Hospital, Bury St Edmunds, UK
[198]James Paget University Hospital NHS Trust, Great Yarmouth, UK
[199]The Christie NHS Foundation Trust, Manchester, UK
[200]The Royal Marsden Hospital, London, UK
[201]University Hospital Hairmyres, East Kilbride, UK
[202]Withybush General Hospital, Pembrokeshire, Wales
[203]Ealing Hospital, Southall, UK
[204]North Devon District Hospital, Barnstaple, UK
[205]St John's Hospital Livingston, Livingston, UK
[206]Northampton General Hospital NHS Trust, Northampton, UK
[207]Harrogate and District NHS Foundation Trust, Harrogate, UK
[208]National Hospital for Neurology and Neurosurgery, London, UK
[209]Bronglais General Hospital, Aberystwyth, UK
[210]Golden Jubilee National Hospital, Clydebank, UK
[211]Homerton University Hospital Foundation NHS Trust, London UK
[212]Sheffield Children's Hospital, Sheffield, UK

### Covid-19 Human Genetics Initiative

A full list of contributors to this project is available at www.covid19hg.org/acknowledgements.

### 23andMe Investigators

Janie F. Shelton*[1], Anjali J. Shastri*[1], Chelsea Ye[1], Catherine H. Weldon[1], Teresa Filshtein-Sonmez[1], Daniella Coker[1], Antony Symons[1], Jorge Esparza-Gordillo[2], The 23andMe COVID-19 Team[1], Stella Aslibekyan[1], Adam Auton[1].

*These authors contributed equally to this work. 1. 23andMe Inc., 223 N Mathilda Ave, Sunnyvale, CA 94086 2. Human genetics - R&D, GSK Medicines Research Centre, Target Sciences-R&D, Stevenage, UK

We thank the 23andMe research participants who made this study possible. We would also like to thank Altovise Ewing, Aaron Petrakovitz, Anne Park, Anne Silk, Aushawna Collins, Becky Macintosh, Carolyn Kao, Courtney Ball, Christine Pai, David Hinds, Devyn Parry, Elo Ratcliff, Emily Bullis, Eric Hall, Farwa Alam, Jacquie Haggarty, Jess Christenson, Jim Lawrence, Jimmy Chau, Josie Shaw, Joe Cackler, Karl Heilbron, Katelyn Kukar, Katie Watson, Marianna Frendo, Olivia Valenti, Ryan Workman, Rachel Lopatin, Robert Bell, Rose Eckert, Sam Rodgers, Sarah Rys,

Shawna Averbeck, Shirin Fuller, Vanessa Lane, and Yunxuan Jiang for contributions and insights.

We also thank the 23andMe Research Team: Barry Hicks, Chao Tian, Devika Dhamija, Elizabeth Babalola, Elizabeth S. Noblin, Ethan M. Jewett, G. David Poznik, Gabriel Cuellar Partida, Jared O'Connell, Jingchunzi Shi, Joanna L. Mountain, Joyce Y. Tung, Katarzyna Bryc, Karen E. Huber, Keng-Han Lin, Kimberly F. McManus, Kipper Fletez-Brant, Marie K. Luff, Matthew H. McIntyre, Maya Lowe, Meghan E. Moreno, Peter Wilton, Pierre Fontanillas, Priyanka Nandakumar, Sahar V. Mozaffari, Sarah L. Elson, Sayantan Das, Steven J. Micheletti, Suyash Shringarpure, Vinh Tran, Wei Wang, Will Freyman, and Xin Wang.

Members of the 23andMe COVID-19 Team are: Adam Auton, Adrian Chubb, Alison Fitch, Alison Kung, Amanda Altman, Andy Kill, Anjali Shastri, Catherine Weldon, Chelsea Ye, Daniella Coker, Janie Shelton, Jason Tan, Jeff Pollard, Jennifer McCreight, Jess Bielenberg, John Matthews, Johnny Lee, Lindsey Tran, Michelle Agee, Monica Royce, Nate Tang, Pooja Gandhi, Raffaello d'Amore, Ruth Tennen, Scott Dvorak, Scott Hadly, Stella Aslibekyan, Sungmin Park, Taylor Morrow, Teresa Filshtein Sonmez, Trung Le, and Yiwen Zheng.
